# Characterising long term Covid-19: a living systematic review

**DOI:** 10.1101/2020.12.08.20246025

**Authors:** Melina Michelen, Vincent Cheng, Lakshmi Manoharan, Natalie Elkheir, Drew Dagens, Claire Hastie, Margaret O’Hara, Jake C. Suett, Dania T. Dahmash, Polina Bugaeva, Ishmeala Rigby, Daniel Munblit, Eli Harriss, Amanda Burls, Carol Foote, Janet T. Scott, Gail Carson, Piero Olliaro, Louise Sigfrid, Charitini Stavropoulou

## Abstract

**Background:** While it is now apparent clinical sequelae (often called Long Covid) may persist after acute Covid-19, their nature, frequency, and aetiology are poorly characterised. This study aims to regularly synthesise evidence on Long Covid characteristics, to inform clinical management, rehabilitation, and interventional studies to improve long term outcomes.

**Methods:** A living systematic review. Medline, CINAHL (EBSCO), Global Health (Ovid), WHO Global Research Database on Covid-19, LitCOVID, and Google Scholar were searched up to 17th March 2021. Published studies including at least 100 people with confirmed or clinically suspected Covid-19 at 12 weeks or more post-onset were included. Results were analysed using descriptive statistics and meta-analyses to estimate prevalence with 95% confidence intervals (CIs).

**Results:** Thirty-nine studies were included: 32 cohort, six cross-sectional, and one case-control. Most showed high or moderate risk of bias. None were set in low-income countries, limited studies included children. Studies reported on 10,951 people (48% female) in 12 countries. Most followed-up post hospital discharge (78%, 8520/10951). The longest mean follow-up was 221.7 (SD: 10.9) days post Covid-19 onset. An extensive range of symptoms with wide prevalence was reported, most commonly weakness (41%; 95% CI 25% to 59%), malaise (33%; 95% CI 15% to 57%), fatigue (31%; 95% CI 24% to 39%), concentration impairment (26%; 95% CI 21% to 32%), and breathlessness (25%; 95% CI 18% to 34%). Other frequent symptoms included musculoskeletal, neurological, and psychological. 37% (95% CI 18% to 60%) of people reported reduced quality of life.

**Conclusion:** Long Covid is a complex condition with heterogeneous symptoms. The nature of the studies precludes a precise case definition or evaluation of risk factors. There is an urgent need for prospective, robust, standardised controlled studies into aetiology, risk factors, and biomarkers to characterise Long Covid in different at-risk populations and settings.

**Systematic review registration:** The protocol was prospectively registered on the PROSPERO database (CRD42020211131).

**Section 1: What is already known?:**

- A significant number of people continue to describe ongoing symptoms long after the acute phase of Covid-19, often referred to as Long Covid.
- Long Covid is a heterogeneous condition with an uncertain prevalence, for which there is currently no precise case definition.

**Section 2: What are the new findings?:**

- This ‘living’ systematic review provides a comprehensive summary of peer-reviewed published evidence on persistent symptoms of Covid-19 and will be regularly updated as new evidence emerges.
- The breadth of reported symptoms suggests a complex, heterogeneous condition affecting both those who were hospitalised and those managed in the community.
- Our review identifies weakness (41%; 95% CI 25% to 59%), general malaise (33%; 95% confidence interval 15% to 57%), fatigue (31%; 95% CI 24% to 39%), concentration impairment (26%; 95% CI 21% to 32%) and breathlessness (25%; 95% CI 18% to 34%) as the most common symptoms.

**Section 3: What do the new findings imply?:**

- The current evidence base of the clinical spectrum of Long Covid is limited, based on heterogenous data, and vulnerable to biases, hence caution should be used when interpreting or generalising the results.
- Our review identifies areas where further Long Covid research is critically needed to help characterise Long Covid in different populations and define its aetiology, risk factors, and biomarkers, as well as the impact on variants of concern and vaccination on long term outcomes.

## INTRODUCTION

The severe acute respiratory syndrome coronavirus 2 (SARS-CoV-2) first emerged in December 2019 causing a widespread pandemic. Most people experience asymptomatic or mild to moderate acute coronavirus disease 2019 (Covid-19) symptoms, whilst around 15% of people are estimated to progress to more severe disease requiring hospitalisation and approximately 5% become critically ill.[1]

While the acute phase of the disease was characterised early, there is still limited data on long term outcomes. Symptoms of long-lasting Covid-19 sequelae and complications, termed Long Covid by patients [2], have been reported worldwide. Yet the underlying aetiology behind prolonged or fluctuating symptomatology is limited and there is no widely-accepted uniformed case definition.[3] Instead, Long Covid has been defined pragmatically as “not recovering [for] several weeks or months following the start of symptoms.”[3] Others have distinguished between post-acute Covid-19, referring to symptoms beyond three weeks and chronic Covid-19 beyond 12 weeks [4], while the National Institute for Health and Care Excellence (NICE) distinguishes between ongoing symptomatic Covid-19 lasting from four to 12 weeks and post-covid-19 syndrome continuing for over 12 weeks.[5]

The number of people living with Long Covid is unknown. Attempts to quantify the prevalence of Long Covid use different methods, including national surveys and patient-led studies, making it difficult to compare across studies. The UK’s Office for National Statistics (ONS) has estimated that on average one in five people have symptoms beyond five weeks, while one in ten have symptoms persisting over 12 weeks.[6] A patient-led survey found that in survival analysis, the chance of full recovery by day 50 was smaller than 20% [7] and a Covid symptom app study found 13.3% had symptoms lasting 28 days or more, 4.5% eight or more weeks and 2.3% had symptoms lasting over 12 weeks.[8]

The symptoms of Long Covid are equally ill-defined, with some people describing it as a fluctuating illness of disparate symptoms. [7,9] Indeed, the National Institute for Health Research (NIHR) has suggested that post-acute Covid-19 may consist of several distinct clinical syndromes including: a post-intensive care syndrome, chronic fatigue syndrome, long-term Covid-19 syndrome and disease from SARS-CoV-2 inflicted organ damage.[10] Additionally, even with an expanding knowledge of risk factors in the acute phase, little is currently known on predictive factors for developing Long Covid.[8] Despite suggested classifications, there is yet no clear consensus.

Our early understanding of Long Covid has been accumulated from case reports and cross-sectional online survey studies as the pandemic global research focus has largely been on studies of hospitalised patients during the acute phase. As the pandemic progresses, emerging studies have followed up people to present the fluctuating multiorgan sequelae of acute Covid-19, yet evidence is still scarce. There continues to be a call to further understand and acknowledge this condition while involving patient knowledge and experiences.[11,12]

Given the enormous number of people worldwide who have suffered from Covid-19, it is essential to establish a precise categorisation of Long Covid. Such categorisation will not only help people better understand their symptoms, but also direct research into prevention, treatment, and support, ultimately allowing us to understand and prepare to respond to the long-term consequences inflicted by the Covid-19 pandemic. Our review seeks to synthesise and continually update the evidence on the character and prevalence of Long Covid.

## METHODS

Systematic reviews conducted early during the Covid-19 pandemic soon became redundant due to the rapidity with which new research was released. In recognition of this, many reviewers have moved towards the concept of a ‘living systematic review’ (LSR), which has in-built mechanisms for regular update and renewal.[13,14] We conducted a ‘living’ systematic review to provide frequently updated evidence on the symptoms and complications of Long Covid. This review was developed in collaboration with infectious disease clinicians, public health professionals, information specialists, review methodologists with experience in clinical epidemic research, and members of the global Long Covid Support Group, that includes people living with Long Covid. This living systematic review will be updated approximately every 6 months as new evidence emerges.

### Protocol registration

This report was structured according to the Preferred Reporting Items for Systematic Reviews and Meta-Analyses (PRISMA) statement guidelines.[15] The protocol was registered with PROSPERO (CRD42020211131) and published in a peer-reviewed journal.[16]

### Search strategy

The following databases were searched: Medline and CINAHL (EBSCO), Global Health (Ovid), WHO Global Research Database on Covid-19, and LitCOVID from 1st January 2020 to 17th March 2021. Additionally, we searched Google Scholar on 17th March 2021, screening the first 500 titles. A ‘backwards’ snowball search was conducted of the references of systematic reviews. Full search terms are included in supplement 1. The search terms and inclusion criteria have, for this first version, been designed to cast a wide net, and will be modified in line with new evidence, research priorities, and clinical and policy needs.

### Eligibility criteria

Peer-reviewed studies were considered eligible if they included at least 100 people with suspected, laboratory confirmed, and/or clinically diagnosed Covid-19. Without a clear or internationally agreed case definition, we included studies that reported symptoms or outcomes assessed at 12 or more weeks post Covid-19 onset.[5] There were no language restrictions. Reviews and opinion pieces were excluded. Studies were excluded if they included fewer than 100 participants or the follow-up was unclear or less than 12 weeks post-onset.

### Screening

Screening was performed independently by two reviewers. Any disagreements were resolved via consensus or a third reviewer. Non-English articles were translated using Google Translate or reviewed by a reviewer with good knowledge of the language. The data were managed using the review software Rayyan.[17]

### Data extraction

Data extraction was performed using Microsoft Excel. A data extraction template informed by a previous review [18] was reviewed, updated, and piloted before being finalised. Data extracted included study design, population characteristics, outcomes, prevalence, duration of symptoms, and risk factors. Data extraction was performed by one reviewer and checked by a second reviewer. Disagreements were resolved through consensus. To avoid duplication of data in future updates and ensure robustness, data extraction was not performed for non-peer-reviewed preprints.

### Risk of bias assessment

The included studies were assessed for risk of bias using a modified version of the tool produced by Hoy *et al*. [19] (Supplement 2). This assessment checklist is a validated tool for assessing risk of bias in prevalence studies. The checklist has ten domains for assessing risk of bias, used to calculate a cumulative overall risk of bias for the whole study.

### Data analysis

We undertook individual descriptive analysis for each study. We presented symptom proportions by different settings, as presented in the individual studies: hospitalised, non-hospitalised, or a mix of both populations if no subset data was available. Symptoms were broadly grouped into physiological clusters through discussion with clinicians. Proportion of symptoms and its 95% confidence intervals (CIs) were estimated using the exact method.[20] If there were two or more studies in each symptom, a meta-analysis was performed using a random intercept logistic regression model with Hartung-Knapp modification due to the heterogeneity and skewed sample sizes.[21,22] Heterogeneity between estimates was assessed using the I^2^ statistic.[23] Additional subgroup analysis was conducted to explore the modification of the following factors on proportion of symptoms: hospitalisation, settings, continents, and follow-up timing. We also conducted meta-regression analysis on the percentage of females and ICU patients where there were more than 10 studies in the symptom. Sensitivity analyses were conducted to examine the impact of high risk of bias studies and statistical methods, Freeman-Tukey Double arcsine transformation using inverse variance meta-analysis, on the estimates. Funnel plots were plotted using proportion of the symptom against the precision and sample sizes [22] where there were more than 10 studies in the symptom to explore risk of publication bias. All analysis and data presentation were performed using *metaprop* and *ggplot2* in R (version 4.0.5) via RStudio (version 1.3.1093). The data is presented using a combination of infographics, prepared by a design company, Design Science, and scientific tables to facilitate interpretation by different stakeholders, including non-specialists.

### Patient and public involvement

The study team includes members who have been affected by long term Covid-19 sequalae (CH, MO, and JCS). CH, MO, and JCS are members of Long Covid Support, a patient support group with global reach, with approximately 40,000 members.

They actively contributed to the development of the study protocol, to inform the research questions and interpretation and presentation of the findings, to communicate the results to different audiences. The results of this LSR will be disseminated to Long Covid patient forums for discussion and feedback to inform research priorities and updates.

## RESULTS

We identified 6,459 studies, of which 39 met the inclusion criteria (Supplement 3). Of these, 32 were included in the meta-analysis. The remaining studies included single symptoms or imaging and diagnostics and are presented narratively.

### Characteristics of included studies

Most studies were set in Europe (62%, 24/39), followed by Asia (23%, 9/39), North America (8%, 3/39), and the Middle East 8% (3/39). There was no study set in a low-middle income (LMIC) country (Figure 1).[24] Most were cohort studies (82%, 32/39), followed by cross sectional studies (15%, 6/39), and a case control study (3%, 1/39).

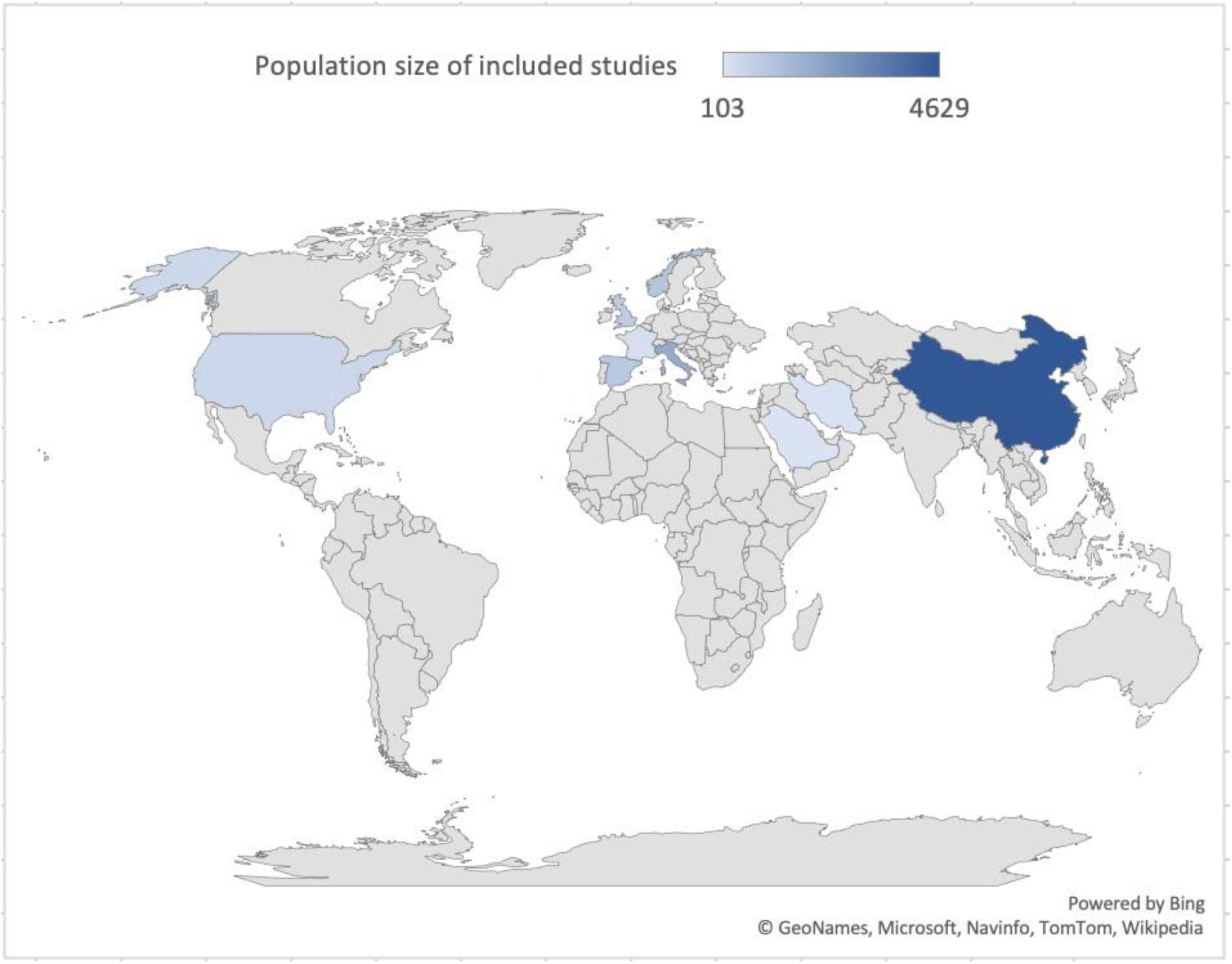

These studies present data on 10,951 (range: 100-1,733) people in 12 countries, aged from 9 months to 93 years old, 48% (5206/10931) were female. Most studies included adults, while 10% (4/39) also included children.[25–28] Only 15% (6/39) of studies reported ethnicity of the participants [29–34], but without stratification. Table 1 presents the included study characteristics.

**Table 1.** Study characteristics

| Study | Design | Country | Population size | Age (years) | Sex (% female) | COVID-19 confirmation method | Follow-up time (days) | Follow-up timepoint | Follow-up mode |
| --- | --- | --- | --- | --- | --- | --- | --- | --- | --- |
| **Non-hospitalised** |  |  |  |  |  |  |  |  |  |
| Hopkins et al. [52] | Cross Sectional | UK | 434 | Median (range): 40 (19-77) | 75 | PCR or serological assays (26.3%) | 6 months | 1st survey | Electronic survey |
| Klein et al. [41] | Cohort (P) | Israel | 103 | Mean (SD): 35 (12) | 38 | PCR (RT-PCR) | 6 months | Onset | Phone interview |
| Petersen et al. [26] | Cohort (P) | Faroe Islands | 180 | Mean (SD; range): 39.9 (19.4; 0-93) | 54 | PCR (RT-PCR) | Mean (SD) 125 (17) | Onset | Phone interview |
| Stavem et al. [62] | Cross Sectional | Norway | 451 | Mean (SD): 49.8 (15.2) | 56 | PCR (RT-PCR) | Median (range): 117 (41-193) | Onset | Outpatient visit and survey |
| **Non-hospitalised & hospitalised** |  |  |  |  |  |  |  |  |  |
| Parente-Arias et al. [49] | Cohort (P) | Spain | 151 | Mean (range): 55.2 (18–88) | 65 | PCR (RT-PCR) | Mean (SD): 100.5 (3.3) | Admission | Phone interview |
| Venturelli et al. [54] | Cohort (P) | Italy | 767 | Mean (SD): 63 (13.6) | 33 | PCR (RT-PCR) (94%); Serology (5%)<br>Clinician diagnosis (1.2%) | Median (IQR): 105 (84–127) | Onset | Outpatient visit |
| Anastasio et al. [35] | Cohort (P) | Italy | 379 | Median (IQR; range): 56 (49-63; 20-80) | 54 | PCR (RT-PCR) | Median (IQR): 135 (102–175) | Onset | Outpatient visit |
| Einvik et al. [61] | Cross Sectional | Norway | 538 | Mean (SD) 57.7 (14.2) (hospital)<br>49.6 (15.3) | 42 (hospital)<br>56 | PCR (RT-PCR) | Mean (SD): 112 (30) (hospital)<br>118 (27) | Onset | Outpatient visit and survey |
| Jacobson et al. [34] | Cohort (P) | USA | 118 | Mean (SD): 43.3 (14.4) | 47 | PCR (RT-PCR) | Mean (SD): 119.3 (33) | Diagnosis | Outpatient visit |
| Logue et al. [29] | Cohort (P) | USA | 177<br>21 (C) | Mean (SD): 48(15.2) | 57 | Lab confirmed | Median (range): 169 (31-300) | Onset | Electronic survey |
| Mazza et al. [64] | Cohort (P) | Italy | 226 | Mean (SD; range): 58 (12.8; 26-87) | 34 | PCR (RT-PCR) | Mean (SD): 90 (13.4) | Discharge | Phone interview |
| Rass et al. [44] | Cohort (P) | Austria | 135 | Median (IQR; range) 56 (48-68; 19-87) | 39 | PCR (RT-PCR) | Median (IQR): 102 (91-110) | Onset | Outpatient visit |
| Sonnweber et al. [42] | Cohort (P) | Austria | 145 | Mean (SD): 57 (14) | 43 | PCR (RT-PCR) | Mean (SD): 103 (21) | Diagnosis | Outpatient visit |
| **Hospitalised** |  |  |  |  |  |  |  |  |  |
| Alharthy et al. [48] | Cohort (P) | Saudi Arabia | 127 | Mean (SD): 47 (11.38) | 21 | PCR (RT-PCR) | 4 months | Discharge | Outpatient visit |
| Arnold et al. [31] | Cohort (P) | UK | 110 | Median (IQR): 60 (46-73) | 38 | PCR or radiological diagnosis | Median (IQR): 90 (80–97) | Onset | Outpatient visit |
| Baricich et al. [57] | Cross Sectional | Italy | 204 | Mean (SD): 57.9 (12.8) | 40 | NR | Mean (SD): 124.7 (17.5) | Discharge | Outpatient visit |
| Bellan et al. [36] | Cohort (P) | Italy | 238 | Median (IQR): 61 (50-71) | 40 | PCR (RT-PCR) (97.5%);<br>Bronchoalveolar lavage (0.4%);<br>Serology/radiological (2.1%) | 3-4 months | Discharge | Outpatient visit |
| Blanco et al. [32] | Cohort (P) | Spain | 100 | Mean (SD)<br>DLCO <80: 54.98 (10.72)<br>DLCO >80: 54.75 (9.83) | 36 | PCR (RT-PCR) | Median (IQR): 104 (89.25-126.75) | Onset | Outpatient visit |
| Doyle et al. [60] | Cohort (P) | UK | 129 | Mean: 62 (Cambridge)<br>56 (London) | 31 (Cambridge)<br>27 (London) | PCR (RT-PCR) | Median (range): 113 (96–138) | Discharge | NR |
| Garrigues et al. [59] | Cohort (P) | France | 120 | Mean (SD): 63.2 (15.7) | 38 | PCR (RT-PCR) | Mean (SD): 110.9 (11.1) | Admission | Phone interview |
| Gherlone et al. [51] | Cohort (P&R) | Italy | 122 | Median (IQR): 62.5 (53.9-74.1) | 25 | PCR (RT-PCR) | Median (IQR): 104 (95-132) | Discharge | Outpatient visit |
| Han et al. [40] | Cohort (P) | China | 114 | Mean (SD; range): 54 (12; 24-82) | 30 | PCR (RT-PCR) | Mean (SD): 175 (20) | Onset | Outpatient visit |
| Huang et al. [50] | Cohort (P&R) | China | 1733 | Median (IQR): 57 (47–65) | 48 | Lab confirmed | Median (IQR): 186 (175–199) | Onset | Outpatient visit |
| Zhang, J et al. [25] | Cohort (R/S) | China | 527 | Median (IQR; range): 42.5 (32–54; 0-91) | 44 | NR | 6 months | Discharge | Outpatient visit |
| Lerum et al. [55] | Cohort (P) | Norway | 103 | Median (25th-75th percentile): 59 (49, 72) | 48 | Nasopharyngeal swab | 3 months | Discharge | Outpatient visit |
| Méndez et al. [43] | Cohort (R/S) | Spain | 215 | Median (IQR): 55 (47-66) | 40 | Lab confirmed | Median (IQR): 87 (62-109) | Discharge | Outpatient visit |
| Nguyen et al. [27] | Cohort (P) | France | 125 | Median (IQR; range): 36 (27-48; 16- 85) | 55 | PCR (RT-PCR) | Mean (SD): 221.7 (10.9) | Onset | Phone interview |
| Nugent et al. [30] | Cohort (R/S) | USA | 182<br>1430 (C) | Median (IQR): 67.4 (58.3-80.1) | 47 | PCR (RT-PCR) | Median (IQR): 92.9 (52.5-127.7) | Discharge | Outpatient visit |
| Qin et al. [47] | Cohort (P) | China | 647 | Mean (SD): 58 (15) years | 56 | PCR (RT-PCR) | 90 | Discharge | Outpatient visit |
| Qu et al. [28] | Cohort (P) | China | 540 | Median (IQR): 47.50 (37-57) | 50 | PCR (RT-PCR) | 3 months | Discharge | Electronic survey |
| Sibila et al. [45] | Cohort (P) | Spain | 172 | Mean (SD): 56.1 (19.8) | 43 | NR | Mean (SD): 101.5 (19.9) | Discharge | Outpatient visit |
| Simani et al. [53] | Cohort (P) | Iran | 120 | Mean (SD): 54.62 (16.94) | 33 | PCR or radiological diagnosis | 6 months | Discharge | Outpatient visit |
| Suárez-Robles et al. [58] | Cross Sectional | Spain | 134 | Mean (SD): 58.53 (18.53) | 54 | PCR (RT-PCR) | 90 | Discharge | Phone survey |
| Sykes et al. [33] | Cohort (P) | UK | 134 | Median (range): 58 (25–89) | 34 | PCR (RT-PCR) | Median (range): 113 (46–167) | Discharge | Outpatient visit |
| Taboada et al. [56] | Cross Sectional | Spain | 183 | Mean (SD): 65.9 (14.1) | 40 | PCR (RT-PCR) | 6 months | Discharge | Unstructured interview |
| Weng et al. [39] | Cohort (P) | China | 117 | 45.3% ≥60years | 44 | Viral nucleic acid test | 90 | Discharge | Phone interview |
| Xiong et. Al [38] | Cohort (P) | China | 538<br>184 (C) | Median (IQR; range): 52 (41-62; 22-79) | 55 | PCR (RT-PCR) | Median (IQR; range): 97.0 (95.0-102.0; 91 - 116) | Discharge | Phone interview |
| Xu et al. [37] | Case control | China | 103<br>27 (C) | Median (IQR)<br>M/M: 56 (45-63)<br>S/C: 61 (55-68) | M/M: 58.8<br>S/C: 53.6 | NR | 3 months | Discharge | Outpatient visit |
| Zhang et al. [46] | Cohort (P) | China | 310 | Median (IQR): 51 (31.8-61) | 50 | PCR (RT-PCR) | Median (IQR): 92.0 (90-100) | Discharge | Outpatient visit |
| C: Control group; IQR; interquartile range; P: Prospective; R: Retrospective; M/M: mild/moderate; NR: Not reported; S/C: severe/critical; SD: Standard deviation; |  |  |  |  |  |  |  |  |  |

Most studies (67%, 26/39) were cohorts of hospitalised patients post-discharge, 10% (4/39) were set in the community (non-hospitalised population) whilst 23% (9/39) included both, with 78% (8520/10951) of individuals hospitalised during acute Covid-19. Twenty-two studies included people requiring ICU admission during the acute phase.[25,27–29,31,32,34–49]

The longest follow-up period in any study was a mean of 221.7 (SD: 10.9) days post-onset. Only 56% (22/39) of studies specified Covid-19 severity [25,27–29,31,32,34–49], 31% (12/39) treatment received during the acute phase [30,34,35,39,40,44,47,50–54], and 62% (24/39) described ventilation support requirements.[30–36,39,40,42–45,47,48,50,51,54–60] Pre-existing comorbidities were reported in the majority of studies (85%, 33/39), with hypertension and diabetes most commonly documented.[27,29–51,53–57,59,61–63]

### Risk of bias

Twelve studies were assessed as high risk of bias, 22 moderate, and five low risk of bias. Most studies had a high risk of bias with regards to the generalisability of their results to the wider population with Covid-19. High risk-of-bias ratings were most common for external validity, with item 1 (representation of target population) and item 3 (random selection) having the most high risk of bias ratings (Supplement 2). Further, the recruitment process and response rates were often not well-described, and several studies applied different data collection methods. Although many studies applied validated measurement methods to assess participants, most were not designed to detect symptoms arising from Covid-19. Only four studies included a comparative control group.[29,30,37,38]

### Symptoms & signs

People suffering from Long Covid report a wide range of new or persistent symptoms, in both the hospitalised and non-hospitalised populations. Symptoms were broadly organised into physiological ‘clusters’ for the purpose of presentation and interpretation of this review (Figure 2).

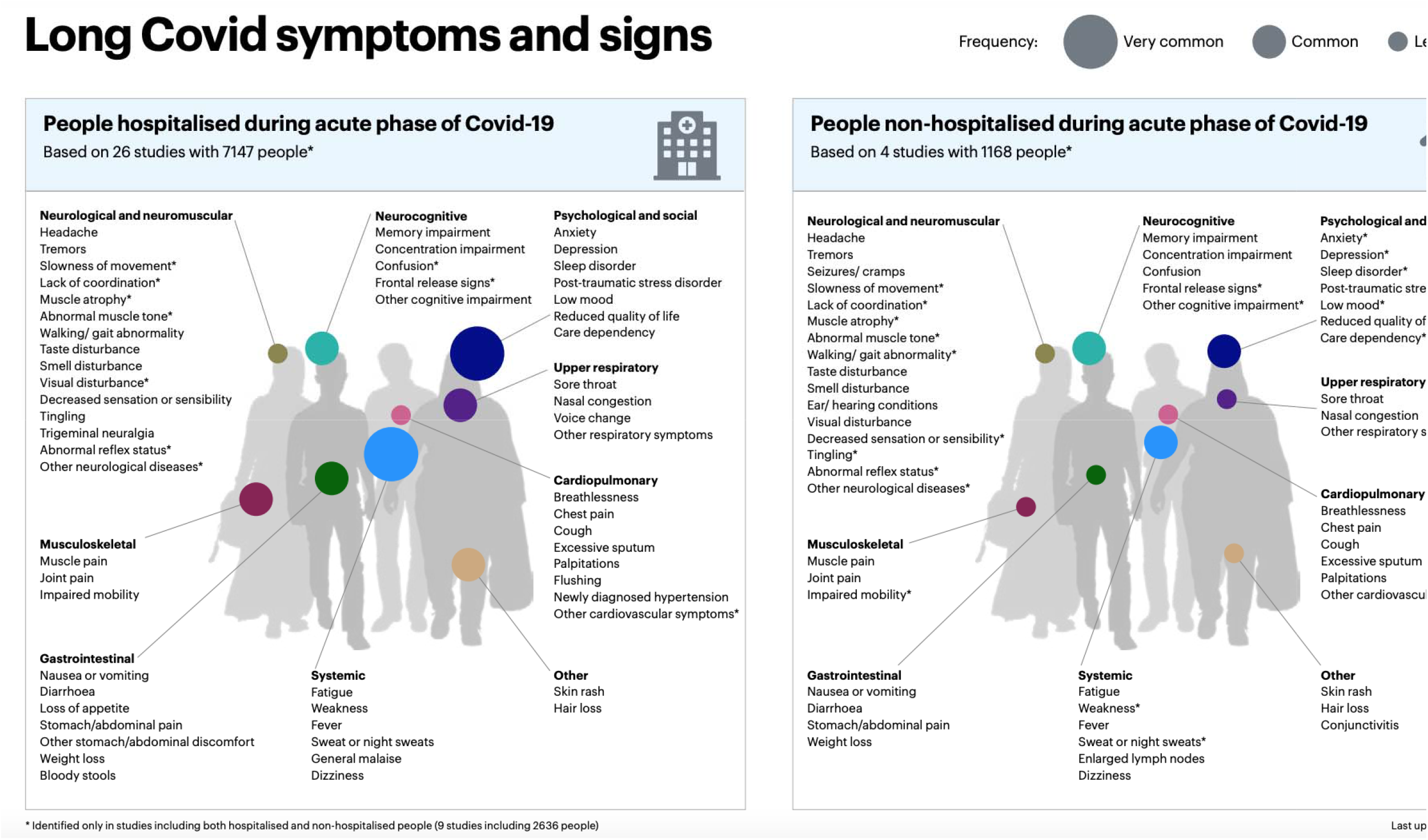

The focus of each study included in our analysis varied. Some authors focused solely on a specialty, such as dentistry, or a specific symptom, such as cognition, making comparative analysis difficult. Even amongst those studies which took a broad approach, the prevalence of symptoms was diverse. Similarly, the prevalence of the more commonly reported symptoms varied markedly.

Within these limitations, we performed a meta-analysis of the most commonly reported symptoms and signs of Long Covid. The most commonly described symptoms (with prevalence of 25% or greater) were weakness (41%, 95% CI 25.43 to 59.01), general malaise (33%, 95% CI 14.91 to 57.36), fatigue (31%, 95% CI 23.91 to 39.03), concentration impairment (26%, 95% CI 20.96 to 31.73) and breathlessness (25%, 95% CI 17.86 to 33.97). Across studies, 37% (95% CI 18.43 to 59.93) of people reported reduced quality of life. Although high I^2^ values (>80%) were observed, they resulted from narrow dispersions in the estimates and well-separated estimates and confidence intervals between studies (Supplement 4). The differences between these symptoms, and the heterogeneity within them is likely to be, to some extent, due to other factors (e.g., study settings, populations, and different measurement tools used).

A diverse array of less prevalent symptoms, including systemic, musculoskeletal, neurological, and psychological symptoms such as sweating, chest pain, sore throat, anxiety, and headaches, amongst others, were also reported. The prevalence of these symptoms was lower, usually less than 20%. Figure 3 presents the range of symptoms.

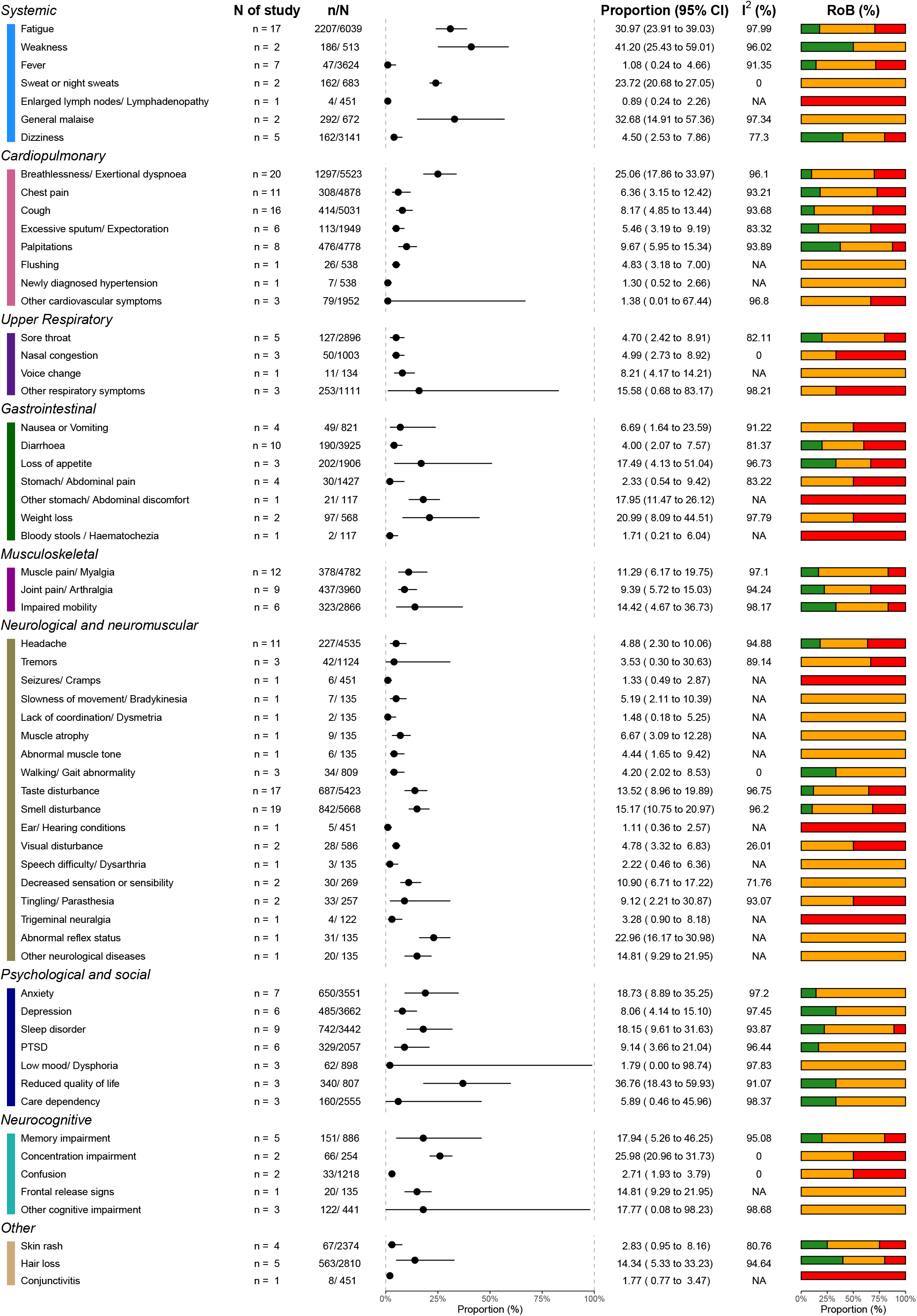

Figure 4 displays this data by population. We also performed subgroup analysis based upon setting (hospitalised vs never hospitalised) and follow-up time. In several symptoms and signs, the heterogeneity of the results was found to be associated with level of hospitalisation, hospital settings, location of the studies, and follow-up timing using subgroup analysis (Supplement 5-8). Using meta-regression, the proportion of females in the studies was positively associated with headache and smell and taste disturbance (Supplement 9), while the proportion of ICU patients in the studies was positively associated with muscle pain (Supplement 10). No major difference was found in the sensitivity analyses (Supplement 11-12). Asymmetries found in the funnel plots suggest reporting biases and limited methodological quality in the included studies (Supplement 13).

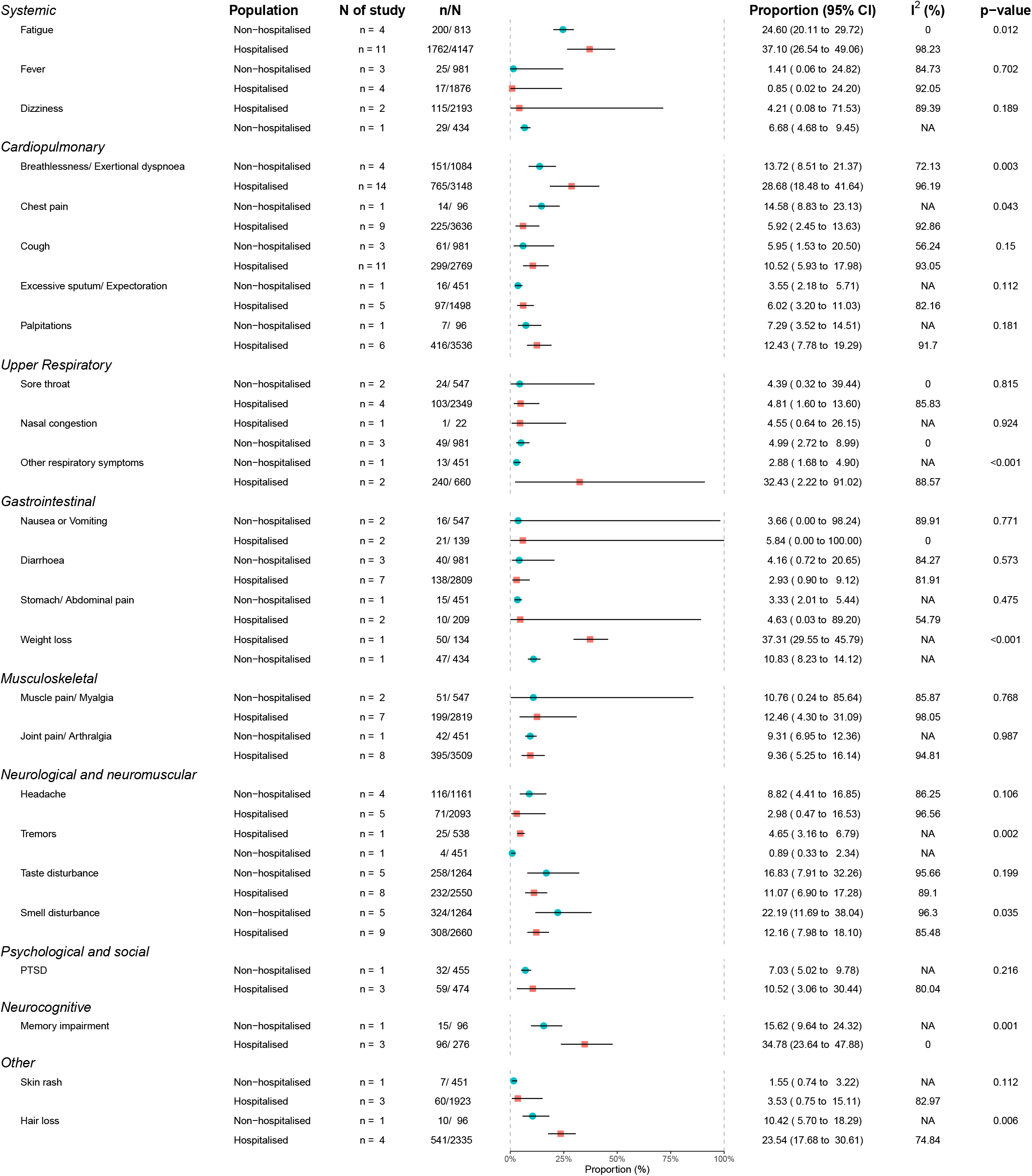

### Imaging & diagnostics

Multiple studies assessed lung sequelae and respiratory performance through outpatient visits follow-up (49%, 19/39).[25,31–37,40,42,43,45–48,50,54,55,60] Imaging results were reported in 33% (13/39) [25,31–33,37,40,42,46–48,50,55,60] of the cohort studies, with one including controls,[37] and one including children.[25] Authors used heterogenous measurement techniques, including artificial intelligence and point-of-care ultrasound.[37,48] Studies found abnormal CT results, including consolidation, reticulation, residual ground glass opacity, interstitial thickening, and fibrotic changes. Some of these studies presented comparisons between initial CT findings and those at follow-up, showing improvements overtime in pulmonary clinical measures and radiologic resolutions at serial visits. [31,33,40,42,48]

Pulmonary function tests were reported in 26% (10/39) of studies [31,32,35–37,42,43,45,47,50,55], including spirometry, diffusion capacity, lung volume, and exercise tests. These studies found evidence of altered pulmonary function, most frequently significant reduction of diffusing lung capacity for carbon monoxide (DLCO). Across these, the study with the longest follow up, at an average of 186 (175–199) days post-hospital discharge, following up adults (median age 57 (IQR 47– 65) years; 52% men) found impaired 6-min walk test in 24–56%, with higher impairment correlating with increased acute phase severity. Using high resolution CT and ultrasonography they detected corresponding lung diffusion impairment in 22% (severity scale 3), 29% (severity scale 4), and 56% (scale 5–6). Median CT scores were 3 (IQR 2–5) for severity scale 3, 4 (3–5) for scale 4, and 5 (4–6) for scale 5-6.[65]

Whereas one study assessing thrombotic complications in Covid-19 with a minimum of 90 days follow-up from critical care admission did not detect any post-discharge venous thromboembolism using CT or doppler, in a cohort of 129 patients.[60]

Another study assessed kidney function in people with Covid-19 –associated acute kidney injury (AKI) compared with people with non-Covid-19 associated AKI, and found that Covid-19–related AKI was associated with decreased kidney recovery during outpatient follow-up (median (IQR): 92.9 (52.5-127.7) days).[30]

### Risk factors

Exploring the literature, we sought to produce a meta-analysis of risk factors for Long Covid. We found a considerable diversity of reported risk factors, including age, sex, comorbidities, ethnicity, and severity of the acute phase.

Several cohorts (64%, 25/39) assessed whether there was an association between the severity of initial Covid-19, including symptom load, level of hospital care, and need for mechanical ventilation, and the risk of persisting sequelae. An association between female gender and risk of prolonged recovery and post-acute SARS-CoV2 sequelae has been identified in longitudinal studies (20.5%, 8/39), as has the association between presence of pre-existing comorbidity,[34,49,51,57,62,66] age,[26,28,44,49,56,57] and minority ethnicity (e.g. Latinx and unspecified)[34,61].

The heterogeneity in study design and evidence base, limited number of studies including non-hospital cohorts, and lack of control groups limits the statistical conclusions or identification of a case definition at this time. We have summarised the reported significant associations to date (Supplement 14) and suggest that these associations be explored in prospective controlled trials to forward the evidence into long term Covid-19 outcomes in different at-risk populations.

## DISCUSSION

Our work represents the most comprehensive review of evidence regarding Long Covid yet produced. Accurate to 17 March 2021, this living systematic review captures the breadth of persistent symptoms reported in 39 studies, including over 10,000 people. These data suggest post-acute Covid-19, or Long Covid, is a multifaceted syndrome affecting several organs, across the acute disease severity spectrum. It is predominantly characterised by marked fatigue, weakness, general malaise, breathlessness, cognitive or concentration impairment, and neuromuscular and psychological sequelae. In a proportion of people, it adversely impacts quality of life. Besides the most commonly reported symptoms, there is a diverse array of additional symptoms.

The findings in this review show symptoms and prevalence aligned to current knowledge on long term Covid-19 outcomes. These findings are in line with the ONS statistical estimates of Long Covid in the UK, reporting 13.7% experienced persistent symptoms for 12 or more weeks, with fatigue most commonly reported. ONS also estimated that prevalence rates of self-reported Long Covid were greatest in females and adults (aged 35 to 69 years old). In addition, prevalence rates were greater for those living in the most deprived areas, working in health or social care, and those with a pre-existing health conditions.[6]

Our data indicates that for some people symptoms of long-term sequelae may be attributed to reduced pulmonary diffusion capacities. This is consistent with recent studies reporting reduced lung perfusion using xenon MRI (XeMRI) three to nine months post Covid-19 hospital discharge [67] and using lung function and high-resolution chest CT 12 months post-onset.

Yet, there is no definite evidence into aetiology or underlying risk factors for post-acute Covid-19 syndromes. Endothelial dysfunction, inflammation, and/or autoimmune events triggering chronic inflammation have been suggested.[68] One recent study identified immunological differences in children with PASC (post-acute Covid-19 syndrome), compared to controls that had recovered fully.

Another study identified a high increase in autoantibody reactivities in Covid-19 patients compared to uninfected controls, with a high prevalence of autoantibodies against immunomodulatory proteins including cytokines, chemokines, complement components, and cell surface proteins.[69] Our findings highlight an urgent need for comprehensive assessments including biochemical, immunological, functional and imaging tests (e.g., MRI and SPECT), in a step-by-step approach guided by initial assessments and symptomatology.

A deeper understanding of Long Covid is currently prevented by the limitations of the published literature. The studies included in our review were highly heterogeneous due to differences in their study designs, settings, populations, follow-up time, and symptom ascertainment methods. In addition, studies used inconsistent terminology describing symptoms and limited details and stratification on pre-existing comorbidities, the severity of Covid-19, and treatment methods. This inconsistency and limited reporting partly explain the high degree of variability observed. The lack of case-control studies prevents a direct attribution of symptoms solely to Covid-19; larger prospective studies with matched control groups are needed. We note that there are large, robust prospective cohort studies of hospitalised patients [70] and non-hospitalised people.[71] Simultaneously, qualitative studies are ongoing to better explore the Long Covid patient experience. [72]

The findings have identified several research gaps and priorities. The majority of Long Covid cohorts were conducted in Western Europe on patients recently discharged from hospital. There is a paucity of evidence on the long-term effects of Covid-19 in low to middle income countries and in people who were not hospitalised. Similarly, there were no studies identified focusing on children, despite evidence showing that children and young people are also affected by Long Covid.[73] Additionally, no study stratified by ethnicity or socioeconomic factors, important risk factors for the acute phase. There is an urgent need to identify appropriate comparative control groups and enhance inclusion of children and people from diverse demographics.

Our review also highlights a need for standardised and validated Covid-19 research tools to harmonise data collection, improve quality and reduce reporting variability. For instance, fatigue is one of the most commonly reported symptoms of Long Covid. However, the symptom alone is not clearly defined and open to different interpretation and requires validated tool such as the Visual Analogue Scale (VAS) graded fatigue scale for robust, objective, and comparative analysis. The International Severe Acute Respiratory and emerging Infection Consortium (ISARIC) has developed open access research tools available to sites globally to facilitate standardisation of data collection, analysis and interpretation for adults and children of an age.[74] We support the broader use of this tool as well as initiatives to standardise outcome measures for Long Covid.

Similarly, our study highlights the need for further research to refine the many circulating interim case definitions and precisely characterise Long Covid, including the potential impacts of variants of concern and vaccination on Long Covid.

As this is a living systematic review, emerging themes from this first version will inform future updates. The LSR will be updated periodically, as new research is published internationally, in order to provide relevant up to date information for clinicians, patients, researchers, policymakers, and health-service commissioners. Version changes will be identified, and previous reports will be archived.

## CONCLUSION

This living systematic review summarises published evidence on the spectrum of long term Covid-19 associated symptoms and sequelae (as of 17 March 2021). It is clear that Long Covid affects different populations, with a wide range of symptomatology. Our findings suggest this multi-organ syndrome is characterised by fatigue, weakness, malaise, breathlessness, and concentration impairment, amongst other less frequent symptoms. Currently the strength of the available evidence is limited and prone to bias. The long-term effects of Covid-19, in both hospitalised and non-hospitalised individuals, including children and at-risk populations, should be a priority for future research using standardised and controlled study designs. Robust research is needed to characterise and define Long Covid, identify risk factors and underlying aetiology, in order to inform prevention, rehabilitation, clinical, and public health management to improve recovery and long-term Covid-19 outcomes. This living systematic review will be updated approximately every six months as new evidence emerges for up to two years.

## Data Availability

All data are available on the paper

## List of Abbreviations

ADL: Activities of daily living
AKI: Acute kidney injury
COVID-19: Coronavirus Disease
DCLO: Diffusion capacity for carbon monoxide
ICU: Intensive care unit
IQR: Interquartile Range
ISARIC: International Severe Acute Respiratory and emerging Infection Consortium
LMIC: Low-middle income country
LSR: Living’ systematic review
NICE: National Institute for Health and Care Excellence
NIHR: National Institute for Health Research
PTSD: post traumatic stress disorder
RoB: Risk of bias
SARS-CoV-2: Severe Acute Respiratory Syndrome Coronavirus-2
SD: Standard deviation
VAS: Visual Analogue Scale

## DECLARATIONS

### Ethics approval and consent to participate

Not applicable

### Consent for publication

Not applicable

### Availability of data and materials

All data generated or analysed during this study are included in this published article and its supplementary information files.

## Authors contributions

MM and CS led on the drafting of the manuscript with contributions from AD, VC, NE, CS, LM, LS, DD CH, MOH, JS, GC, PO, EH. MM, NE, LM, VC, CS critically appraised the studies. VC led on the presentation of the results. CS, MM, VC, LS conceptualised the study, all authors contributed to the final protocol, the interpretation and analysis of the results and reviewed and approved the manuscript.

## Acknowledgments

We would like to thank the members of the Long Covid support group, and the ISARIC Global Support Centre.

