## Supplement 1, Supplement 2, Supplement 3 for "Characterising long term Covid-19: a living systematic review"

#### Supplement 1: Search strategy summary

| Database | Strategy | Results (16/3/2021) |
|---|---|---|
| **Medline (EBSCOhost)** | (COVID-19 OR covid OR SARS-CoV-2. ab)<br>AND<br>(symptom* OR "clinical features" OR signs OR characteristic* OR sequela* OR complication*.ab)<br>AND<br>("long-term Covid" OR long-term OR consequence* OR "long-term impact" OR "long-term effect" OR "post-acute" OR long-tail OR persist* OR "chronic-COVID" OR "long-COVID" OR post-discharge OR postdischarge OR "prolonged symptom" OR "long-haul" .ab)<br>**Limits**: 2020- | 1952 |
| **CINAHL (EBSCOhost)** | (COVID-19 OR covid OR SARS-CoV-2. ab)<br>AND<br>(symptom* OR "clinical features" OR signs OR characteristic* OR sequela* OR complication*.ab)<br>AND<br>("long-term Covid" OR long-term OR consequence* OR "long-term impact" OR "long-term effect" OR "post-acute" OR long-tail OR persist* OR "chronic-COVID" OR "long-COVID" OR post-discharge OR postdischarge OR "prolonged symptom" OR "long-haul" .ab)<br>**Limits**: 2020- | 384 |
| **Global Health** | (COVID-19 or covid or SARS-CoV-2)<br>AND<br>(symptom* or "clinical features" or signs or characteristic* or sequelae or complication*)<br>AND<br>((("long-term Covid" or long-term) adj2 consequence*) or "long-term impact" or "long-term effect" or "post-acute" or long-tail or persist* or "chronic-COVID" or "long-COVID" or post-discharge or postdischarge or "prolonged symptom" or "long-haul")).ab.<br>**Limits**: 2020- | 35 |
| **WHO COVID-19 (WHO COVID, ELSEVIER and Lanzhou University/CNKI)** | tw:((ab:(covid-19 OR covid OR sars-cov-2)) AND (ab:(symptom OR "clinical features" OR signs OR characteristic OR sequela OR complication)) AND (ab:("long-term Covid" OR "long-term consequence" OR "long-term impact" OR "long-term effect" OR "post-acute" OR long-tail OR persist* OR "chronic-COVID" OR "long-COVID" OR post-discharge OR postdischarge OR "prolonged symptom" OR "long-haul"))) AND db:("COVIDWHO" OR "ELSEVIER" OR "CNKI_Lanzhou") | 195 |

| | | |
|---|---|---|
| **Lit Covid** | (“persistent symptoms” OR “after covid-19 infection").ti,ab,kw<br>OR<br>((“outcomes “ OR “characteristics” OR “features” OR “symptoms” OR “inflammation” OR “function” OR “complications” OR “syndrome” OR “manifestation”) ADJ10 (“long-haul” OR “recovery” OR “recovered” OR “recovering” OR “survivors” OR “post-discharge” OR “postdischarge” OR “discharge” OR “persisting” OR “prolonged” OR “long-term” OR “after admission” OR “post-COVID-19” OR “post-COVID”)).ti,ab.<br>OR<br>((“outcomes “ OR “characteristics” OR “features” OR “symptoms” OR “inflammation” OR “function” OR “complications” OR “syndrome” OR “manifestation”) ADJ/10 (“after admission” OR “after hospital” OR “after hospitalisation" OR “after hospitalization" OR “after COVID-19” OR “after SARS-CoV-2")).ti,ab. | 1432 |
| **Google Scholar** | post COVID after discharge persistent symptom | 1000 |
| **Ovid Embase (top-up) [17 Mar 2021]** | See Appendix 1<br>**Limit**: 2020- | 483 |
| **Ovid Medline (top-up) [17 Mar 2021]** | See Appendix 2<br>**Limit**: 2020- | 336 |
| **WHO (top-up)<br>[19 Mar 2021]<br>(excluded: PREPRINT-BIORXVI, PREPRINT-MEDRXVI, PREPRINT-other preprint, ITCRP)** | See Appendix 3 | 340 |

**Appendix 1**

Database(s): **Embase** 1974 to 2021 March 17

| # | Searches | Results |
|---|---|---|
| 1 | (long* adj3 (covid* or ncov* or novel coronavirus or novel betacoronavirus or sars-ncov-2 or sars-cov-2)).mp. | 462 |
| 2 | (persist* adj3 (covid* or ncov* or novel coronavirus or novel betacoronavirus or sars-ncov-2 or sars-cov-2)).mp. | 265 |
| 3 | (chronic adj3 (covid* or ncov* or novel coronavirus or novel betacoronavirus or sars-ncov-2 or sars-cov-2)).mp. | 191 |
| 4 | ((long term or long-term or longterm) adj3 effect* adj3 (covid* or ncov* or novel coronavirus or novel betacoronavirus or sars-ncov-2 or sars-cov-2)).mp. | 59 |
| 5 | (sequela* adj3 (covid* or ncov* or novel coronavirus or novel betacoronavirus or sars-ncov-2 or sars-cov-2)).mp. | 150 |
| 6 | ((post acute or post-acute or postacute) adj3 (covid* or ncov* or novel coronavirus or novel betacoronavirus or sars-ncov-2 or sars-cov-2)).mp. | 38 |
| 7 | ((longhaul* or long haul* or long-haul*) adj3 (covid* or ncov* or novel coronavirus or novel betacoronavirus or sars-ncov-2 or sars-cov-2)).mp. | 18 |
| 8 | ((post-covid* or postcovid*) adj2 (syndrome or condition)).mp. | 32 |
| 9 | 1 or 2 or 3 or 4 or 5 or 6 or 7 or 8 | 1123 |
| 10 | symptom/ | 151132 |
| 11 | symptom assessment/ | 8515 |
| 12 | exp complication/ | 1237024 |
| 13 | (symptom* or "clinical feature*" or signs or characteristic* or sequela* or complication*).mp. | 6906139 |
| 14 | exp physical disease by body function/ | 9306422 |
| 15 | 10 or 11 or 12 or 13 or 14 | 13028765 |
| 16 | 9 and 15 | 748 |
| 17 | limit 16 to yr="2020" | 483 |

**Appendix 2**

Database(s): **Ovid MEDLINE(R) and Epub Ahead of Print, In-Process, In-Data-Review & Other Non-Indexed Citations and Daily** 1946 to March 17, 2021

| # | Searches | Results |
|---|---|---|
| 1 | (long* adj3 (covid* or ncov* or novel coronavirus or novel betacoronavirus or sars-ncov-2 or sars-cov-2)).mp. | 507 |
| 2 | (persist* adj3 (covid* or ncov* or novel coronavirus or novel betacoronavirus or sars-ncov-2 or sars-cov-2)).mp. | 273 |
| 3 | (chronic adj3 (covid* or ncov* or novel coronavirus or novel betacoronavirus or sars-ncov-2 or sars-cov-2)).mp. | 173 |
| 4 | ((long term or long-term or longterm) adj3 effect* adj3 (covid* or ncov* or novel coronavirus or novel betacoronavirus or sars-ncov-2 or sars-cov-2)).mp. | 62 |
| 5 | (sequela* adj3 (covid* or ncov* or novel coronavirus or novel betacoronavirus or sars-ncov-2 or sars-cov-2)).mp. | 159 |
| 6 | ((post acute or post-acute or postacute) adj3 (covid* or ncov* or novel coronavirus or novel betacoronavirus or sars-ncov-2 or sars-cov-2)).mp. | 66 |
| 7 | ((longhaul* or long haul* or long-haul*) adj3 (covid* or ncov* or novel coronavirus or novel betacoronavirus or sars-ncov-2 or sars-cov-2)).mp. | 25 |
| 8 | ((post-covid* or postcovid*) adj2 (syndrome or condition)).mp. | 36 |

| | | |
|---|---|---|
| 9 | 1 or 2 or 3 or 4 or 5 or 6 or 7 or 8 | 1179 |
| 10 | exp "signs and symptoms"/ | 2127623 |
| 11 | (symptom* or "clinical feature*" or signs or characteristic* or sequela* or complication*).tw. | 3644236 |
| 12 | 10 or 11 | 5362470 |
| 13 | 9 and 12 | 591 |
| 14 | limit 13 to yr="2020" | 336 |

**Appendix 3**

There are only three strings of syntax:

1. keywords and phrases associated with "long COVID"
2. keywords and phrases associated with "hospitalisation" and "quarantine"
3. keywords and phrases associated with "symptoms" and "complications"

The combinations of 1 AND 3 and 2 AND 3 were used to search in the combinations of title, abstract and TW (title + abstract + subjects) in the database. These consist of 18 searches as follows:

| # | Syntax | Hits |
|---|---|---|
| 1 | (**ti**:(post-COVID* OR post-nCov* OR "post novel coronavirus" OR "post novel betacoronavirus" OR "post SARS-nCoV-2" OR "post SARS-CoV-2" OR postCOVID* OR "post nCov*" OR "long* COVID*" OR "long* nCov*" OR "long* novel coronavirus" OR "long* novel betacoronavirus" OR "long* SARS-nCoV-2" OR "long* SARS-CoV-2" OR "long-term COVID*" OR "long-term nCov*" OR "long-term novel coronavirus" OR "long-term novel betacoronavirus" OR "long-term SARS-nCoV-2" OR "long-term SARS-CoV-2" OR "long-term COVID*" OR "longterm COVID*" OR "post acute COVID*" OR "postacute COVID*" OR "longhaul* COVID*" OR "long haul* COVID*" OR "long-haul* COVID*" OR "chronic* COVID*" OR "prolonged* COVID*" OR "presist* COVID*" OR "long-term nCov*" OR "longterm nCov*" OR "post acute nCov*" OR "postacute nCov*" OR "longhaul* nCov*" OR "long haul* nCov*" OR "long-haul* nCov*" OR "chronic* nCov*" OR "prolonged* nCov*" OR "presist* nCov*" OR "long-term novel coronavirus" OR "longterm novel coronavirus" OR "post acute novel coronavirus" OR "postacute novel coronavirus" OR "longhaul* novel coronavirus" OR "long haul* novel coronavirus" OR "long-haul* novel coronavirus" OR "chronic* novel coronavirus" OR "prolonged* novel coronavirus" OR "presist* novel coronavirus" OR "long-term novel betacoronavirus" OR "longterm novel betacoronavirus" OR "post acute novel betacoronavirus" OR "postacute novel betacoronavirus" OR "longhaul* novel betacoronavirus" OR "long haul* novel betacoronavirus" OR "long-haul* novel betacoronavirus" OR "chronic* novel betacoronavirus" OR "prolonged* novel betacoronavirus" OR "presist* novel betacoronavirus" OR "long-term SARS-nCoV-2" OR "longterm SARS-nCoV-2" OR "post acute SARS-nCoV-2" OR "postacute SARS-nCoV-2" OR "longhaul* SARS-nCoV-2" OR "long haul* SARS-nCoV-2" OR "long-haul* SARS-nCoV-2" OR "chronic* SARS-nCoV-2" OR "prolonged* SARS-nCoV-2" OR "presist* SARS-nCoV-2" OR "long-term SARS-CoV-2" OR "longterm SARS-CoV-2" OR "post acute SARS-CoV-2" OR "postacute SARS-CoV-2" OR "longhaul* SARS-CoV-2" OR "long haul* SARS-CoV-2" OR "long-haul* SARS-CoV-2" OR "chronic* SARS-CoV-2" OR "prolonged* SARS-CoV-2" OR "presist* SARS-CoV-2") **AND ti**:("clinical feature*" OR abnormal* OR characteristic* OR complication* OR condition* OR convalescence OR disorder* OR dysfunction OR illness* OR impair* OR inflammation OR manifestation OR outcome* OR prevalence OR problem* OR sequela* OR sign* OR symptom* OR syndrome)) | 135 |
| 2 | (**ti**:("after admission" OR "after discharg*" OR "after hospital*" OR "after isolat*" OR "after quarantine" OR "after self isolat*" OR "after self-isolat*" OR "after self-quarantine" OR "post admission" OR "post discharg*" OR "post hospital*" OR "post isolat*" OR "post quarantine" OR "post self isolat*" OR "post self-isolat*" OR "post self-quarantine") **AND ti**:("clinical feature*" OR abnormal* OR characteristic* OR complication* OR condition* OR convalescence OR disorder* OR dysfunction OR illness* OR impair* OR inflammation OR manifestation OR outcome* OR prevalence OR problem* OR sequela* OR sign* OR symptom* OR syndrome)) | 11 |

| | | |
|---|---|---|
| 3 | (**ti**:(post-COVID* OR post-nCov* OR "post novel coronavirus" OR "post novel betacoronavirus" OR "post SARS-nCoV-2" OR "post SARS-CoV-2" OR postCOVID* OR "post nCov*" OR "long* COVID*" OR "long* nCov*" OR "long* novel coronavirus" OR "long* novel betacoronavirus" OR "long* SARS-nCoV-2" OR "long* SARS-CoV-2" OR "long-term COVID*" OR "long-term nCov*" OR "long-term novel coronavirus" OR "long-term novel betacoronavirus" OR "long-term SARS-nCoV-2" OR "long-term SARS-CoV-2" OR "long-term COVID*" OR "longterm COVID*" OR "post acute COVID*" OR "postacute COVID*" OR "longhaul* COVID*" OR "long haul* COVID*" OR "long-haul* COVID*" OR "chronic* COVID*" OR "prolonged* COVID*" OR "presist* COVID*" OR "long-term nCov*" OR "longterm nCov*" OR "post acute nCov*" OR "postacute nCov*" OR "longhaul* nCov*" OR "long haul* nCov*" OR "long-haul* nCov*" OR "chronic* nCov*" OR "prolonged* nCov*" OR "presist* nCov*" OR "long-term novel coronavirus" OR "longterm novel coronavirus" OR "post acute novel coronavirus" OR "postacute novel coronavirus" OR "longhaul* novel coronavirus" OR "long haul* novel coronavirus" OR "long-haul* novel coronavirus" OR "chronic* novel coronavirus" OR "prolonged* novel coronavirus" OR "presist* novel coronavirus" OR "long-term novel betacoronavirus" OR "longterm novel betacoronavirus" OR "post acute novel betacoronavirus" OR "postacute novel betacoronavirus" OR "longhaul* novel betacoronavirus" OR "long haul* novel betacoronavirus" OR "long-haul* novel betacoronavirus" OR "chronic* novel betacoronavirus" OR "prolonged* novel betacoronavirus" OR "presist* novel betacoronavirus" OR "long-term SARS-nCoV-2" OR "longterm SARS-nCoV-2" OR "post acute SARS-nCoV-2" OR "postacute SARS-nCoV-2" OR "longhaul* SARS-nCoV-2" OR "long haul* SARS-nCoV-2" OR "long-haul* SARS-nCoV-2" OR "chronic* SARS-nCoV-2" OR "prolonged* SARS-nCoV-2" OR "presist* SARS-nCoV-2" OR "long-term SARS-CoV-2" OR "longterm SARS-CoV-2" OR "post acute SARS-CoV-2" OR "postacute SARS-CoV-2" OR "longhaul* SARS-CoV-2" OR "long haul* SARS-CoV-2" OR "long-haul* SARS-CoV-2" OR "chronic* SARS-CoV-2" OR "prolonged* SARS-CoV-2" OR "presist* SARS-CoV-2") **AND ab**:("clinical feature*" OR abnormal* OR characteristic* OR complication* OR condition* OR convalescence OR disorder* OR dysfunction OR illness* OR impair* OR inflammation OR manifestation OR outcome* OR prevalence OR problem* OR sequela* OR sign* OR symptom* OR syndrome)) | 352 |
| 4 | (**ti**:("after admission" OR "after discharg*" OR "after hospital*" OR "after isolat*" OR "after quarantine" OR "after self isolat*" OR "after self-isolat*" OR "after self-quarantine" OR "post admission" OR "post discharg*" OR "post hospital*" OR "post isolat*" OR "post quarantine" OR "post self isolat*" OR "post self-isolat*" OR "post self-quarantine") **AND ab**:("clinical feature*" OR abnormal* OR characteristic* OR complication* OR condition* OR convalescence OR disorder* OR dysfunction OR illness* OR impair* OR inflammation OR manifestation OR outcome* OR prevalence OR problem* OR sequela* OR sign* OR symptom* OR syndrome)) | 35 |
| 5 | (**ti**:(post-COVID* OR post-nCov* OR "post novel coronavirus" OR "post novel betacoronavirus" OR "post SARS-nCoV-2" OR "post SARS-CoV-2" OR postCOVID* OR "post nCov*" OR "long* COVID*" OR "long* nCov*" OR "long* novel coronavirus" OR "long* novel betacoronavirus" OR "long* SARS-nCoV-2" OR "long* SARS-CoV-2" OR "long-term COVID*" OR "long-term nCov*" OR "long-term novel coronavirus" OR "long-term novel betacoronavirus" OR "long-term SARS-nCoV-2" OR "long-term SARS-CoV-2" OR "long-term COVID*" OR "longterm COVID*" OR "post acute COVID*" OR "postacute COVID*" OR "longhaul* COVID*" OR "long haul* COVID*" OR "long-haul* COVID*" OR "chronic* COVID*" OR "prolonged* COVID*" OR "presist* COVID*" OR "long-term nCov*" OR "longterm nCov*" OR "post acute nCov*" OR "postacute nCov*" OR "longhaul* nCov*" OR "long haul* nCov*" OR "long-haul* nCov*" OR "chronic* nCov*" OR "prolonged* nCov*" OR "presist* nCov*" OR "long-term novel coronavirus" OR "longterm novel coronavirus" OR "post acute novel coronavirus" OR "postacute novel coronavirus" OR "longhaul* novel coronavirus" OR "long haul* novel coronavirus" OR "long-haul* novel coronavirus" OR "chronic* novel coronavirus" OR "prolonged* novel coronavirus" OR "presist* novel coronavirus" OR "long-term novel betacoronavirus" OR "longterm novel betacoronavirus" OR "post acute novel betacoronavirus" OR "postacute novel betacoronavirus" OR "longhaul* novel betacoronavirus" OR "long haul* novel betacoronavirus" OR "long-haul* novel betacoronavirus" OR "chronic* novel betacoronavirus" OR "prolonged* novel betacoronavirus" OR "presist* novel betacoronavirus" OR "long-term SARS-nCoV-2" OR "longterm SARS-nCoV-2" OR "post acute SARS-nCoV-2" OR "postacute SARS-nCoV-2" OR "longhaul* SARS-nCoV-2" OR "long haul* SARS-nCoV-2" OR "long-haul* SARS-nCoV-2" OR "chronic* SARS-nCoV-2" OR "prolonged* SARS-nCoV-2" OR "presist* SARS-nCoV-2" OR "long-term SARS-CoV-2" OR "longterm SARS-CoV-2" OR "post acute SARS-CoV-2" OR "postacute SARS-CoV-2" OR "longhaul* SARS-CoV-2" OR "long haul* SARS-CoV-2" OR "long-haul* SARS-CoV-2" OR "chronic* SARS-CoV-2" OR "prolonged* SARS-CoV-2" OR "presist* SARS-CoV-2") **AND tw**:("clinical feature*" OR abnormal* OR characteristic* OR complication* OR condition* OR convalescence OR disorder* OR dysfunction OR illness* OR impair* OR inflammation OR manifestation OR outcome* OR prevalence OR problem* OR sequela* OR sign* OR symptom* OR syndrome)) | 508 |
| 6 | (**ti**:("after admission" OR "after discharg*" OR "after hospital*" OR "after isolat*" OR "after quarantine" OR "after self isolat*" OR "after self-isolat*" OR "after self-quarantine" OR "post admission" OR "post discharg*" OR "post hospital*" OR "post isolat*" OR "post quarantine" OR "post self isolat*" OR "post self-isolat*" OR "post self-quarantine") **AND tw**:("clinical feature*" OR abnormal* OR characteristic* OR complication* OR condition* OR convalescence OR disorder* OR dysfunction OR illness* OR impair* OR inflammation OR manifestation OR outcome* OR prevalence OR problem* OR sequela* OR sign* OR symptom* OR syndrome)) | 38 |
| 7 | (**ab**:(post-COVID* OR post-nCov* OR "post novel coronavirus" OR "post novel betacoronavirus" OR "post SARS-nCoV-2" OR "post SARS-CoV-2" OR postCOVID* OR "post nCov*" OR "long* COVID*" OR "long* nCov*" OR "long* novel coronavirus" OR "long* novel betacoronavirus" OR "long* SARS-nCoV-2" OR "long* SARS-CoV-2" OR "long-term COVID*" | 159 |

| | | |
|---|---|---|
| | OR "long-term nCov*" OR "long-term novel coronavirus" OR "long-term novel betacoronavirus" OR "long-term SARS-nCoV-2" OR "long-term SARS-CoV-2" OR "long-term COVID*" OR "longterm COVID*" OR "post acute COVID*" OR "postacute COVID*" OR "longhaul* COVID*" OR "long haul* COVID*" OR "long-haul* COVID*" OR "chronic* COVID*" OR "prolonged* COVID*" OR "presist* COVID*" OR "long-term nCov*" OR "longterm nCov*" OR "post acute nCov*" OR "postacute nCov*" OR "longhaul* nCov*" OR "long haul* nCov*" OR "long-haul* nCov*" OR "chronic* nCov*" OR "prolonged* nCov*" OR "presist* nCov*" OR "long-term novel coronavirus" OR "longterm novel coronavirus" OR "post acute novel coronavirus" OR "postacute novel coronavirus" OR "longhaul* novel coronavirus" OR "long haul* novel coronavirus" OR "long-haul* novel coronavirus" OR "chronic* novel coronavirus" OR "prolonged* novel coronavirus" OR "presist* novel coronavirus" OR "long-term novel betacoronavirus" OR "longterm novel betacoronavirus" OR "post acute novel betacoronavirus" OR "postacute novel betacoronavirus" OR "longhaul* novel betacoronavirus" OR "long haul* novel betacoronavirus" OR "long-haul* novel betacoronavirus" OR "chronic* novel betacoronavirus" OR "prolonged* novel betacoronavirus" OR "presist* novel betacoronavirus" OR "long-term SARS-nCoV-2" OR "longterm SARS-nCoV-2" OR "post acute SARS-nCoV-2" OR "postacute SARS-nCoV-2" OR "longhaul* SARS-nCoV-2" OR "long haul* SARS-nCoV-2" OR "long-haul* SARS-nCoV-2" OR "chronic* SARS-nCoV-2" OR "prolonged* SARS-nCoV-2" OR "presist* SARS-nCoV-2" OR "long-term SARS-CoV-2" OR "longterm SARS-CoV-2" OR "post acute SARS-CoV-2" OR "postacute SARS-CoV-2" OR "longhaul* SARS-CoV-2" OR "long haul* SARS-CoV-2" OR "long-haul* SARS-CoV-2" OR "chronic* SARS-CoV-2" OR "prolonged* SARS-CoV-2" OR "presist* SARS-CoV-2") **AND ti**:("clinical feature*" OR abnormal* OR characteristic* OR complication* OR condition* OR convalescence OR disorder* OR dysfunction OR illness* OR impair* OR inflammation OR manifestation OR outcome* OR prevalence OR problem* OR sequela* OR sign* OR symptom* OR syndrome)) | |
| 8 | (**ab**:("after admission" OR "after discharg*" OR "after hospital*" OR "after isolat*" OR "after quarantine" OR "after self isolat*" OR "after self-isolat*" OR "after self-quarantine" OR "post admission" OR "post discharg*" OR "post hospital*" OR "post isolat*" OR "post quarantine" OR "post self isolat*" OR "post self-isolat*" OR "post self-quarantine") **AND ti**:("clinical feature*" OR abnormal* OR characteristic* OR complication* OR condition* OR convalescence OR disorder* OR dysfunction OR illness* OR impair* OR inflammation OR manifestation OR outcome* OR prevalence OR problem* OR sequela* OR sign* OR symptom* OR syndrome)) | 202 |
| 9 | (**ab**:(post-COVID* OR post-nCov* OR "post novel coronavirus" OR "post novel betacoronavirus" OR "post SARS-nCoV-2" OR "post SARS-CoV-2" OR postCOVID* OR "post nCov*" OR "long* COVID*" OR "long* nCov*" OR "long* novel coronavirus" OR "long* novel betacoronavirus" OR "long* SARS-nCoV-2" OR "long* SARS-CoV-2" OR "long-term COVID*" OR "long-term nCov*" OR "long-term novel coronavirus" OR "long-term novel betacoronavirus" OR "long-term SARS-nCoV-2" OR "long-term SARS-CoV-2" OR "long-term COVID*" OR "longterm COVID*" OR "post acute COVID*" OR "postacute COVID*" OR "longhaul* COVID*" OR "long haul* COVID*" OR "long-haul* COVID*" OR "chronic* COVID*" OR "prolonged* COVID*" OR "presist* COVID*" OR "long-term nCov*" OR "longterm nCov*" OR "post acute nCov*" OR "postacute nCov*" OR "longhaul* nCov*" OR "long haul* nCov*" OR "long-haul* nCov*" OR "chronic* nCov*" OR "prolonged* nCov*" OR "presist* nCov*" OR "long-term novel coronavirus" OR "longterm novel coronavirus" OR "post acute novel coronavirus" OR "postacute novel coronavirus" OR "longhaul* novel coronavirus" OR "long haul* novel coronavirus" OR "long-haul* novel coronavirus" OR "chronic* novel coronavirus" OR "prolonged* novel coronavirus" OR "presist* novel coronavirus" OR "long-term novel betacoronavirus" OR "longterm novel betacoronavirus" OR "post acute novel betacoronavirus" OR "postacute novel betacoronavirus" OR "longhaul* novel betacoronavirus" OR "long haul* novel betacoronavirus" OR "long-haul* novel betacoronavirus" OR "chronic* novel betacoronavirus" OR "prolonged* novel betacoronavirus" OR "presist* novel betacoronavirus" OR "long-term SARS-nCoV-2" OR "longterm SARS-nCoV-2" OR "post acute SARS-nCoV-2" OR "postacute SARS-nCoV-2" OR "longhaul* SARS-nCoV-2" OR "long haul* SARS-nCoV-2" OR "long-haul* SARS-nCoV-2" OR "chronic* SARS-nCoV-2" OR "prolonged* SARS-nCoV-2" OR "presist* SARS-nCoV-2" OR "long-term SARS-CoV-2" OR "longterm SARS-CoV-2" OR "post acute SARS-CoV-2" OR "postacute SARS-CoV-2" OR "longhaul* SARS-CoV-2" OR "long haul* SARS-CoV-2" OR "long-haul* SARS-CoV-2" OR "chronic* SARS-CoV-2" OR "prolonged* SARS-CoV-2" OR "presist* SARS-CoV-2") **AND ab**:("clinical feature*" OR abnormal* OR characteristic* OR complication* OR condition* OR convalescence OR disorder* OR dysfunction OR illness* OR impair* OR inflammation OR manifestation OR outcome* OR prevalence OR problem* OR sequela* OR sign* OR symptom* OR syndrome)) | 800 |
| 10 | (**ab**:("after admission" OR "after discharg*" OR "after hospital*" OR "after isolat*" OR "after quarantine" OR "after self isolat*" OR "after self-isolat*" OR "after self-quarantine" OR "post admission" OR "post discharg*" OR "post hospital*" OR "post isolat*" OR "post quarantine" OR "post self isolat*" OR "post self-isolat*" OR "post self-quarantine") **AND ab**:("clinical feature*" OR abnormal* OR characteristic* OR complication* OR condition* OR convalescence OR disorder* OR dysfunction OR illness* OR impair* OR inflammation OR manifestation OR outcome* OR prevalence OR problem* OR sequela* OR sign* OR symptom* OR syndrome)) | 680 |
| 11 | (**ab**:(post-COVID* OR post-nCov* OR "post novel coronavirus" OR "post novel betacoronavirus" OR "post SARS-nCoV-2" OR "post SARS-CoV-2" OR postCOVID* OR "post nCov*" OR "long* COVID*" OR "long* nCov*" OR "long* novel coronavirus" OR "long* novel betacoronavirus" OR "long* SARS-nCoV-2" OR "long* SARS-CoV-2" OR "long-term COVID*" OR "long-term nCov*" OR "long-term novel coronavirus" OR "long-term novel betacoronavirus" OR "long-term SARS-nCoV-2" OR "long-term SARS-CoV-2" OR "long-term COVID*" OR "longterm COVID*" OR "post acute COVID*" OR "postacute COVID*" OR "longhaul* COVID*" OR "long haul* COVID*" OR "long-haul* COVID*" OR "chronic* | 830 |

| | | |
|---|---|---|
| | COVID*" OR "prolonged* COVID*" OR "presist* COVID*" OR "long-term nCov*" OR "longterm nCov*" OR "post acute nCov*" OR "postacute nCov*" OR "longhaul* nCov*" OR "long haul* nCov*" OR "long-haul* nCov*" OR "chronic* nCov*" OR "prolonged* nCov*" OR "presist* nCov*" OR "long-term novel coronavirus" OR "longterm novel coronavirus" OR "post acute novel coronavirus" OR "postacute novel coronavirus" OR "longhaul* novel coronavirus" OR "long haul* novel coronavirus" OR "long-haul* novel coronavirus" OR "chronic* novel coronavirus" OR "prolonged* novel coronavirus" OR "presist* novel coronavirus" OR "long-term novel betacoronavirus" OR "longterm novel betacoronavirus" OR "post acute novel betacoronavirus" OR "postacute novel betacoronavirus" OR "longhaul* novel betacoronavirus" OR "long haul* novel betacoronavirus" OR "long-haul* novel betacoronavirus" OR "chronic* novel betacoronavirus" OR "prolonged* novel betacoronavirus" OR "presist* novel betacoronavirus" OR "long-term SARS-nCoV-2" OR "longterm SARS-nCoV-2" OR "post acute SARS-nCoV-2" OR "postacute SARS-nCoV-2" OR "longhaul* SARS-nCoV-2" OR "long haul* SARS-nCoV-2" OR "long-haul* SARS-nCoV-2" OR "chronic* SARS-nCoV-2" OR "prolonged* SARS-nCoV-2" OR "presist* SARS-nCoV-2" OR "long-term SARS-CoV-2" OR "longterm SARS-CoV-2" OR "post acute SARS-CoV-2" OR "postacute SARS-CoV-2" OR "longhaul* SARS-CoV-2" OR "long haul* SARS-CoV-2" OR "long-haul* SARS-CoV-2" OR "chronic* SARS-CoV-2" OR "prolonged* SARS-CoV-2" OR "presist* SARS-CoV-2") **AND tw**:("clinical feature*" OR abnormal* OR characteristic* OR complication* OR condition* OR convalescence OR disorder* OR dysfunction OR illness* OR impair* OR inflammation OR manifestation OR outcome* OR prevalence OR problem* OR sequela* OR sign* OR symptom* OR syndrome)) | |
| 12 | (**ab**:("after admission" OR "after discharg*" OR "after hospital*" OR "after isolat*" OR "after quarantine" OR "after self isolat*" OR "after self-isolat*" OR "after self-quarantine" OR "post admission" OR "post discharg*" OR "post hospital*" OR "post isolat*" OR "post quarantine" OR "post self isolat*" OR "post self-isolat*" OR "post self-quarantine") **AND tw**:("clinical feature*" OR abnormal* OR characteristic* OR complication* OR condition* OR convalescence OR disorder* OR dysfunction OR illness* OR impair* OR inflammation OR manifestation OR outcome* OR prevalence OR problem* OR sequela* OR sign* OR symptom* OR syndrome)) | 681 |
| 13 | (**tw**:(post-COVID* OR post-nCov* OR "post novel coronavirus" OR "post novel betacoronavirus" OR "post SARS-nCoV-2" OR "post SARS-CoV-2" OR postCOVID* OR "post nCov*" OR "long* COVID*" OR "long* nCov*" OR "long* novel coronavirus" OR "long* novel betacoronavirus" OR "long* SARS-nCoV-2" OR "long* SARS-CoV-2" OR "long-term COVID*" OR "long-term nCov*" OR "long-term novel coronavirus" OR "long-term novel betacoronavirus" OR "long-term SARS-nCoV-2" OR "long-term SARS-CoV-2" OR "long-term COVID*" OR "longterm COVID*" OR "post acute COVID*" OR "postacute COVID*" OR "longhaul* COVID*" OR "long haul* COVID*" OR "long-haul* COVID*" OR "chronic* COVID*" OR "prolonged* COVID*" OR "presist* COVID*" OR "long-term nCov*" OR "longterm nCov*" OR "post acute nCov*" OR "postacute nCov*" OR "longhaul* nCov*" OR "long haul* nCov*" OR "long-haul* nCov*" OR "chronic* nCov*" OR "prolonged* nCov*" OR "presist* nCov*" OR "long-term novel coronavirus" OR "longterm novel coronavirus" OR "post acute novel coronavirus" OR "postacute novel coronavirus" OR "longhaul* novel coronavirus" OR "long haul* novel coronavirus" OR "long-haul* novel coronavirus" OR "chronic* novel coronavirus" OR "prolonged* novel coronavirus" OR "presist* novel coronavirus" OR "long-term novel betacoronavirus" OR "longterm novel betacoronavirus" OR "post acute novel betacoronavirus" OR "postacute novel betacoronavirus" OR "longhaul* novel betacoronavirus" OR "long haul* novel betacoronavirus" OR "long-haul* novel betacoronavirus" OR "chronic* novel betacoronavirus" OR "prolonged* novel betacoronavirus" OR "presist* novel betacoronavirus" OR "long-term SARS-nCoV-2" OR "longterm SARS-nCoV-2" OR "post acute SARS-nCoV-2" OR "postacute SARS-nCoV-2" OR "longhaul* SARS-nCoV-2" OR "long haul* SARS-nCoV-2" OR "long-haul* SARS-nCoV-2" OR "chronic* SARS-nCoV-2" OR "prolonged* SARS-nCoV-2" OR "presist* SARS-nCoV-2" OR "long-term SARS-CoV-2" OR "longterm SARS-CoV-2" OR "post acute SARS-CoV-2" OR "postacute SARS-CoV-2" OR "longhaul* SARS-CoV-2" OR "long haul* SARS-CoV-2" OR "long-haul* SARS-CoV-2" OR "chronic* SARS-CoV-2" OR "prolonged* SARS-CoV-2" OR "presist* SARS-CoV-2") **AND** ti:("clinical feature*" OR abnormal* OR characteristic* OR complication* OR condition* OR convalescence OR disorder* OR dysfunction OR illness* OR impair* OR inflammation OR manifestation OR outcome* OR prevalence OR problem* OR sequela* OR sign* OR symptom* OR syndrome)) | 167 |
| 14 | (**tw**:("after admission" OR "after discharg*" OR "after hospital*" OR "after isolat*" OR "after quarantine" OR "after self isolat*" OR "after self-isolat*" OR "after self-quarantine" OR "post admission" OR "post discharg*" OR "post hospital*" OR "post isolat*" OR "post quarantine" OR "post self isolat*" OR "post self-isolat*" OR "post self-quarantine") **AND** ti:("clinical feature*" OR abnormal* OR characteristic* OR complication* OR condition* OR convalescence OR disorder* OR dysfunction OR illness* OR impair* OR inflammation OR manifestation OR outcome* OR prevalence OR problem* OR sequela* OR sign* OR symptom* OR syndrome)) | 93 |
| 15 | (**tw**:(post-COVID* OR post-nCov* OR "post novel coronavirus" OR "post novel betacoronavirus" OR "post SARS-nCoV-2" OR "post SARS-CoV-2" OR postCOVID* OR "post nCov*" OR "long* COVID*" OR "long* nCov*" OR "long* novel coronavirus" OR "long* novel betacoronavirus" OR "long* SARS-nCoV-2" OR "long* SARS-CoV-2" OR "long-term COVID*" OR "long-term nCov*" OR "long-term novel coronavirus" OR "long-term novel betacoronavirus" OR "long-term SARS-nCoV-2" OR "long-term SARS-CoV-2" OR "long-term COVID*" OR "longterm COVID*" OR "post acute COVID*" OR "postacute COVID*" OR "longhaul* COVID*" OR "long haul* COVID*" OR "long-haul* COVID*" OR "chronic* COVID*" OR "prolonged* COVID*" OR "presist* COVID*" OR "long-term nCov*" OR "longterm nCov*" OR "post acute nCov*" OR "postacute nCov*" OR "longhaul* nCov*" OR "long haul* nCov*" OR "long-haul* nCov*" OR "chronic* nCov*" OR "prolonged* nCov*" OR "presist* nCov*" OR "long-term novel coronavirus" OR "longterm novel | 752 |

| | | |
|---|---|---|
| | coronavirus" OR "post acute novel coronavirus" OR "postacute novel coronavirus" OR "longhaul* novel coronavirus" OR "long haul* novel coronavirus" OR "long-haul* novel coronavirus" OR "chronic* novel coronavirus" OR "prolonged* novel coronavirus" OR "presist* novel coronavirus" OR "long-term novel betacoronavirus" OR "longterm novel betacoronavirus" OR "post acute novel betacoronavirus" OR "postacute novel betacoronavirus" OR "longhaul* novel betacoronavirus" OR "long haul* novel betacoronavirus" OR "long-haul* novel betacoronavirus" OR "chronic* novel betacoronavirus" OR "prolonged* novel betacoronavirus" OR "presist* novel betacoronavirus" OR "long-term SARS-nCoV-2" OR "longterm SARS-nCoV-2" OR "post acute SARS-nCoV-2" OR "postacute SARS-nCoV-2" OR "longhaul* SARS-nCoV-2" OR "long haul* SARS-nCoV-2" OR "long-haul* SARS-nCoV-2" OR "chronic* SARS-nCoV-2" OR "prolonged* SARS-nCoV-2" OR "presist* SARS-nCoV-2" OR "long-term SARS-CoV-2" OR "longterm SARS-CoV-2" OR "post acute SARS-CoV-2" OR "postacute SARS-CoV-2" OR "longhaul* SARS-CoV-2" OR "long haul* SARS-CoV-2" OR "long-haul* SARS-CoV-2" OR "chronic* SARS-CoV-2" OR "prolonged* SARS-CoV-2" OR "presist* SARS-CoV-2") **AND ab**:("clinical feature*" OR abnormal* OR characteristic* OR complication* OR condition* OR convalescence OR disorder* OR dysfunction OR illness* OR impair* OR inflammation OR manifestation OR outcome* OR prevalence OR problem* OR sequela* OR sign* OR symptom* OR syndrome)) | |
| 16 | (**tw**:("after admission" OR "after discharg*" OR "after hospital*" OR "after isolat*" OR "after quarantine" OR "after self isolat*" OR "after self-isolat*" OR "after self-quarantine" OR "post admission" OR "post discharg*" OR "post hospital*" OR "post isolat*" OR "post quarantine" OR "post self isolat*" OR "post self-isolat*" OR "post self-quarantine") **AND ab**:("clinical feature*" OR abnormal* OR characteristic* OR complication* OR condition* OR convalescence OR disorder* OR dysfunction OR illness* OR impair* OR inflammation OR manifestation OR outcome* OR prevalence OR problem* OR sequela* OR sign* OR symptom* OR syndrome)) | 339 |
| 17 | (**tw**:(post-COVID* OR post-nCov* OR "post novel coronavirus" OR "post novel betacoronavirus" OR "post SARS-nCoV-2" OR "post SARS-CoV-2" OR postCOVID* OR "post nCov*" OR "long* COVID*" OR "long* nCov*" OR "long* novel coronavirus" OR "long* novel betacoronavirus" OR "long* SARS-nCoV-2" OR "long* SARS-CoV-2" OR "long-term COVID*" OR "long-term nCov*" OR "long-term novel coronavirus" OR "long-term novel betacoronavirus" OR "long-term SARS-nCoV-2" OR "long-term SARS-CoV-2" OR "long-term COVID*" OR "longterm COVID*" OR "post acute COVID*" OR "postacute COVID*" OR "longhaul* COVID*" OR "long haul* COVID*" OR "long-haul* COVID*" OR "chronic* COVID*" OR "prolonged* COVID*" OR "presist* COVID*" OR "long-term nCov*" OR "longterm nCov*" OR "post acute nCov*" OR "postacute nCov*" OR "longhaul* nCov*" OR "long haul* nCov*" OR "long-haul* nCov*" OR "chronic* nCov*" OR "prolonged* nCov*" OR "presist* nCov*" OR "long-term novel coronavirus" OR "longterm novel coronavirus" OR "post acute novel coronavirus" OR "postacute novel coronavirus" OR "longhaul* novel coronavirus" OR "long haul* novel coronavirus" OR "long-haul* novel coronavirus" OR "chronic* novel coronavirus" OR "prolonged* novel coronavirus" OR "presist* novel coronavirus" OR "long-term novel betacoronavirus" OR "longterm novel betacoronavirus" OR "post acute novel betacoronavirus" OR "postacute novel betacoronavirus" OR "longhaul* novel betacoronavirus" OR "long haul* novel betacoronavirus" OR "long-haul* novel betacoronavirus" OR "chronic* novel betacoronavirus" OR "prolonged* novel betacoronavirus" OR "presist* novel betacoronavirus" OR "long-term SARS-nCoV-2" OR "longterm SARS-nCoV-2" OR "post acute SARS-nCoV-2" OR "postacute SARS-nCoV-2" OR "longhaul* SARS-nCoV-2" OR "long haul* SARS-nCoV-2" OR "long-haul* SARS-nCoV-2" OR "chronic* SARS-nCoV-2" OR "prolonged* SARS-nCoV-2" OR "presist* SARS-nCoV-2" OR "long-term SARS-CoV-2" OR "longterm SARS-CoV-2" OR "post acute SARS-CoV-2" OR "postacute SARS-CoV-2" OR "longhaul* SARS-CoV-2" OR "long haul* SARS-CoV-2" OR "long-haul* SARS-CoV-2" OR "chronic* SARS-CoV-2" OR "prolonged* SARS-CoV-2" OR "presist* SARS-CoV-2") **AND tw**:("clinical feature*" OR abnormal* OR characteristic* OR complication* OR condition* OR convalescence OR disorder* OR dysfunction OR illness* OR impair* OR inflammation OR manifestation OR outcome* OR prevalence OR problem* OR sequela* OR sign* OR symptom* OR syndrome)) | 906 |
| 18 | (**tw**:("after admission" OR "after discharg*" OR "after hospital*" OR "after isolat*" OR "after quarantine" OR "after self isolat*" OR "after self-isolat*" OR "after self-quarantine" OR "post admission" OR "post discharg*" OR "post hospital*" OR "post isolat*" OR "post quarantine" OR "post self isolat*" OR "post self-isolat*" OR "post self-quarantine") **AND tw**:("clinical feature*" OR abnormal* OR characteristic* OR complication* OR condition* OR convalescence OR disorder* OR dysfunction OR illness* OR impair* OR inflammation OR manifestation OR outcome* OR prevalence OR problem* OR sequela* OR sign* OR symptom* OR syndrome)) | 340 |

#### Supplement 2: Risk of bias assessment

| Study | Representation of national population (e.g. age, sex, occupation) | Sampling frame true or close representation of target population | Random selection used to select sample, OR, census undertaken | Likelihood of non-response bias minimal | Data collected directly from subjects (opposed to proxy) | Acceptable case definition used | Instrument to measure parameter of interest has reliability and validity (if necessary) | Same mode of data collection used for all subjects | Length of shortest prevalence period for parameter of interest appropriate | Numerator(s)/ denominator(s) for parameter of interest appropriate | Overall risk of bias |
|---|---|---|---|---|---|---|---|---|---|---|---|
| Alharthy et al. | High | High | High | Low | Low | Low | Low | Low | Low | Low | Moderate |
| Anastasio et al. | Low | Low | Low | Low | Low | Low | Low | Low | Low | Low | Low |
| Arnold et al. | High | Low | High | Low | Low | Low | Low | Low | Low | Low | Moderate |
| Baricich et al. | High | High | High | Low | Low | Low | Low | Low | Low | High | High |
| Bellan et al. | High | Low | High | Low | Low | Low | Low | Low | Low | Low | Moderate |
| Blanco et al. | High | High | High | Low | Low | Low | Low | Low | Low | Low | Moderate |
| Doyle et al. | High | High | High | Low | Low | Low | Low | Low | Low | Low | Moderate |
| Einvik et al. | High | Low | High | Low | Low | Low | Low | Low | Low | Low | Moderate |
| Garrigues et al. | High | High | High | High | Low | Low | Low | Low | Low | Low | High |
| Gherlone et al. | High | High | High | Low | Low | Low | Low | Low | Low | High | High |
| Han et al. | High | Low | High | Low | Low | Low | Low | Low | Low | Low | Moderate |
| Hopkins et al. | High | High | High | Low | Low | Low | High | Low | Low | Low | High |
| Huang et al. | Low | Low | Low | High | Low | Low | Low | Low | Low | Low | Low |
| Jacobson et al. | High | Low | High | Low | Low | Low | Low | Low | Low | Low | Moderate |
| Klein et al. | High | High | High | Low | Low | Low | High | Low | Low | Low | High |
| Lerum et al. | High | Low | High | Low | Low | Low | Low | Low | High | Low | Moderate |
| Logue et al. | High | Low | High | Low | Low | Low | Low | Low | Low | Low | Moderate |
| Mazza et al. | High | High | High | Low | Low | Low | Low | Low | Low | Low | Moderate |
| Mendez et al. | High | High | High | Low | Low | Low | Low | Low | High | Low | High |
| Nguyen et al. | High | Low | Low | Low | Low | Low | High | Low | Low | Low | Moderate |
| Nugent et al. | High | Low | High | Low | Low | Low | Low | Low | Low | Low | Moderate |
| Parente-Arias et al. | High | High | High | Low | Low | Low | Low | Low | Low | Low | Moderate |
| Petersen et al. | Low | Low | High | High | Low | Low | High | Low | High | Low | High |
| Qin et al. | High | High | High | High | Low | Low | Low | Low | Low | Low | High |
| Qu et al. | Low | Low | High | Low | Low | Low | Low | Low | Low | Low | Low |
| Rass et al. | High | High | High | Low | Low | Low | Low | Low | Low | Low | Moderate |
| Sibila et al. | High | High | High | Low | Low | Low | Low | Low | Low | High | High |
| Simani et al. | Low | Low | Low | High | Low | Low | Low | Low | Low | Low | Low |
| Sonnweber et al. | High | Low | High | Low | Low | Low | Low | Low | Low | Low | Moderate |
| Stavem et al. | Low | High | High | Low | Low | Low | High | Low | High | Low | High |
| Suarez-Robles et al. | High | High | High | Low | Low | Low | Low | Low | Low | Low | Moderate |
| Sykes et al. | High | Low | Low | High | Low | Low | Low | Low | Low | Low | Moderate |
| Taboada et al. | High | High | High | Low | Low | Low | Low | Low | Low | Low | Moderate |
| Venturelli et al. | Low | High | High | High | Low | Low | Low | Low | Low | Low | Moderate |
| Weng et al. | High | High | High | High | Low | Low | Low | Low | Low | High | High |
| Xiong et al. | Low | High | High | Low | Low | Low | Low | Low | Low | Low | Moderate |
| Xu et al. | High | High | Low | Low | Low | Low | Low | Low | Low | Low | Moderate |
| Zhang et al. (a) | High | High | High | High | Low | Low | Low | Low | Low | High | High |
| Zhang et al. (b) | High | Low | Low | Low | Low | Low | Low | Low | Low | Low | Low |

Legend: Low Risk | Moderate risk | High risk

#### Supplement 3: PRISMA diagram

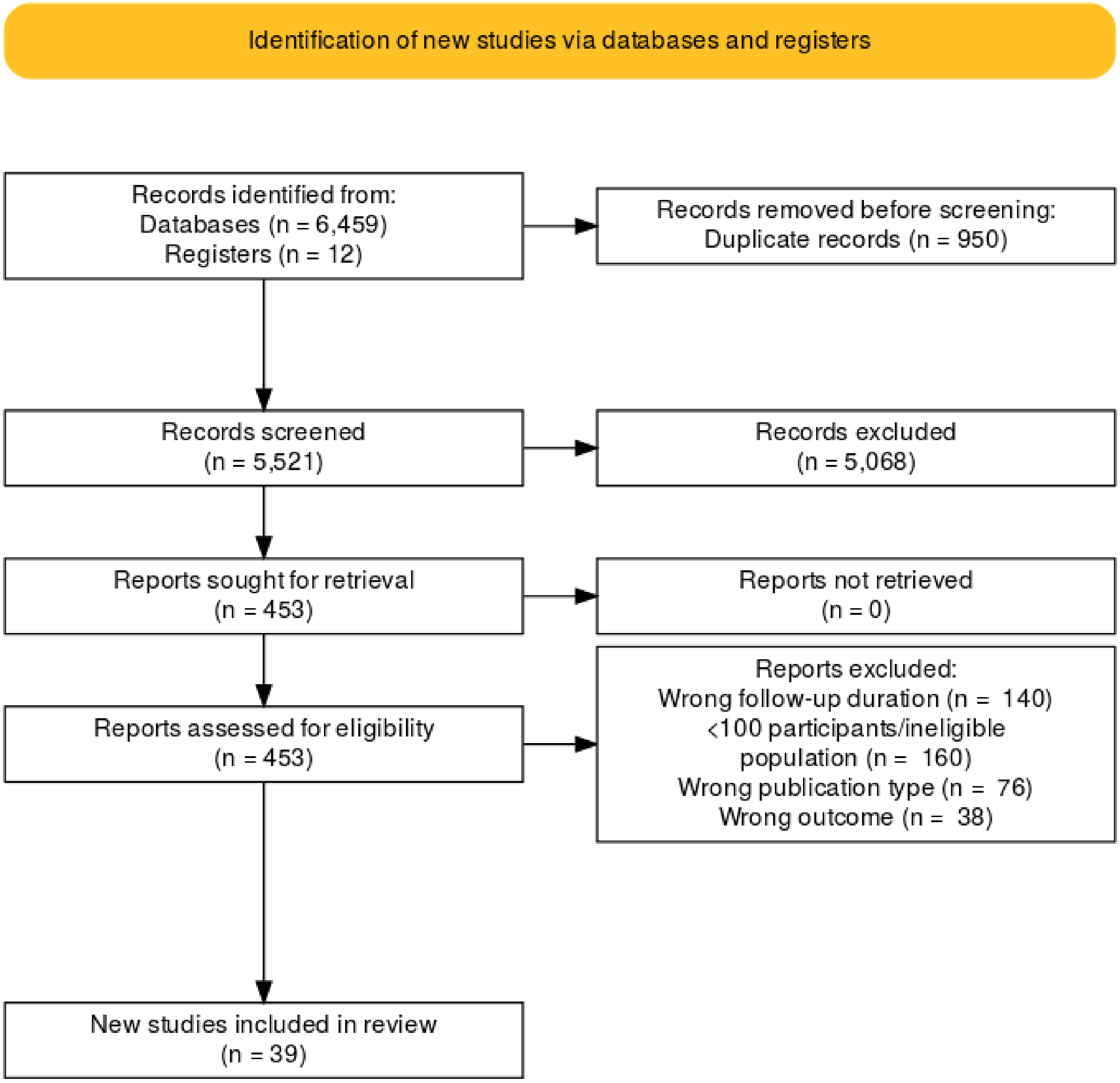

### Supplement 4: Individual forest plots in main results

*Figure 1. Cardiopulmonary*

| Study | n | Total | Proportion (%) | 95% CI |
|---|---|---|---|---|
| **Breathlessness/ Exertional dyspnoea** | | | | |
| Bellan et al. | 13 | 238 | 5.46 | [ 2.94; 9.16] |
| Klein et al. | 8 | 103 | 7.77 | [ 3.41; 14.73] |
| Qin et al. | 56 | 647 | 8.66 | [ 6.60; 11.09] |
| Taboada et al. | 19 | 183 | 10.38 | [ 6.37; 15.74] |
| Hopkins et al. | 50 | 434 | 11.52 | [ 8.67; 14.91] |
| Han et al. | 16 | 114 | 14.04 | [ 8.24; 21.79] |
| Stavem et al. | 73 | 451 | 16.19 | [12.91; 19.92] |
| Xiong et al. | 115 | 538 | 21.38 | [17.98; 25.09] |
| Venturelli et al. | 167 | 767 | 21.77 | [18.90; 24.86] |
| Qu et al. | 141 | 540 | 26.11 | [22.45; 30.03] |
| Jacobson et al. | 31 | 118 | 26.27 | [18.60; 35.17] |
| Sonnweber et al. | 52 | 145 | 35.86 | [28.07; 44.24] |
| Arnold et al. | 43 | 110 | 39.09 | [29.93; 48.86] |
| Sibila et al. | 68 | 172 | 39.53 | [32.18; 47.26] |
| Suárez-Robles et al. | 54 | 134 | 40.30 | [31.92; 49.11] |
| Garrigues et al. | 50 | 120 | 41.67 | [32.74; 51.02] |
| Anastasio et al. | 162 | 379 | 42.74 | [37.71; 47.90] |
| Alharthy et al. | 62 | 127 | 48.82 | [39.85; 57.84] |
| Lerum et al. | 37 | 69 | 53.62 | [41.20; 65.72] |
| Sykes et al. | 80 | 134 | 59.70 | [50.89; 68.08] |
| **Random effects** | **1297** | **5523** | **25.06** | **[18.27; 33.36]** |
| **Hartung-Knapp correction** | | | **25.06** | **[17.86; 33.97]** |
| **Prediction interval** | . | . | | **[ 4.59; 69.93]** |
| Heterogeneity: $I^2 = 96\%$, $\tau^2 = 0.81$, $p < 0.01$ | | | | |
| **Chest pain** | | | | |
| Bellan et al. | 1 | 238 | 0.42 | [ 0.01; 2.32] |
| Qin et al. | 6 | 647 | 0.93 | [ 0.34; 2.01] |
| Venturelli et al. | 24 | 767 | 3.13 | [ 2.01; 4.62] |
| Huang et al. | 75 | 1655 | 4.53 | [ 3.58; 5.65] |
| Sibila et al. | 14 | 172 | 8.14 | [ 4.52; 13.28] |
| Garrigues et al. | 13 | 120 | 10.83 | [ 5.90; 17.81] |
| Anastasio et al. | 45 | 379 | 11.87 | [ 8.79; 15.56] |
| Arnold et al. | 14 | 110 | 12.73 | [ 7.14; 20.43] |
| Jacobson et al. | 16 | 118 | 13.56 | [ 7.95; 21.08] |
| Xiong et al. | 76 | 538 | 14.13 | [11.29; 17.36] |
| Sykes et al. | 24 | 134 | 17.91 | [11.83; 25.47] |
| **Random effects** | **308** | **4878** | **6.36** | **[ 3.43; 11.49]** |
| **Hartung-Knapp correction** | | | **6.36** | **[ 3.15; 12.42]** |
| **Prediction interval** | . | . | | **[ 0.56; 45.11]** |
| Heterogeneity: $I^2 = 93\%$, $\tau^2 = 1.11$, $p < 0.01$ | | | | |
| **Cough** | | | | |
| Jacobson et al. | 1 | 118 | 0.85 | [ 0.02; 4.63] |
| Venturelli et al. | 8 | 767 | 1.04 | [ 0.45; 2.04] |
| Bellan et al. | 6 | 238 | 2.52 | [ 0.93; 5.41] |
| Qin et al. | 38 | 647 | 5.87 | [ 4.19; 7.97] |
| Stavem et al. | 27 | 451 | 5.99 | [ 3.98; 8.59] |
| Anastasio et al. | 23 | 379 | 6.07 | [ 3.89; 8.97] |
| Han et al. | 7 | 114 | 6.14 | [ 2.50; 12.24] |
| Xiong et al. | 38 | 538 | 7.06 | [ 5.05; 9.57] |
| Hopkins et al. | 33 | 434 | 7.60 | [ 5.29; 10.51] |
| Qu et al. | 63 | 540 | 11.67 | [ 9.08; 14.68] |
| Arnold et al. | 13 | 110 | 11.82 | [ 6.45; 19.36] |
| Garrigues et al. | 20 | 120 | 16.67 | [10.49; 24.56] |
| Sonnweber et al. | 23 | 135 | 17.04 | [11.12; 24.46] |
| Sibila et al. | 31 | 172 | 18.02 | [12.59; 24.60] |
| Suárez-Robles et al. | 36 | 134 | 26.87 | [19.58; 35.20] |
| Sykes et al. | 47 | 134 | 35.07 | [27.04; 43.79] |
| **Random effects** | **414** | **5031** | **8.17** | **[ 5.06; 12.93]** |
| **Hartung-Knapp correction** | | | **8.17** | **[ 4.85; 13.44]** |
| **Prediction interval** | . | . | | **[ 0.95; 45.20]** |
| Heterogeneity: $I^2 = 94\%$, $\tau^2 = 1.01$, $p < 0.01$ | | | | |

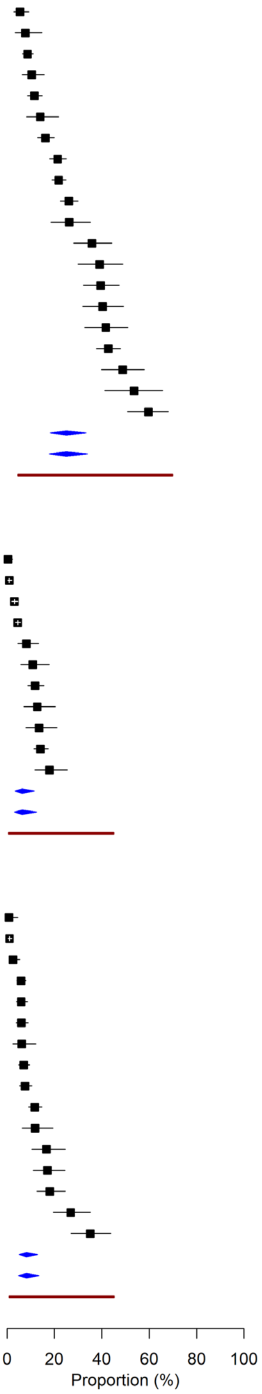

*Figure 2.Cardiopulmonary (page 2)*

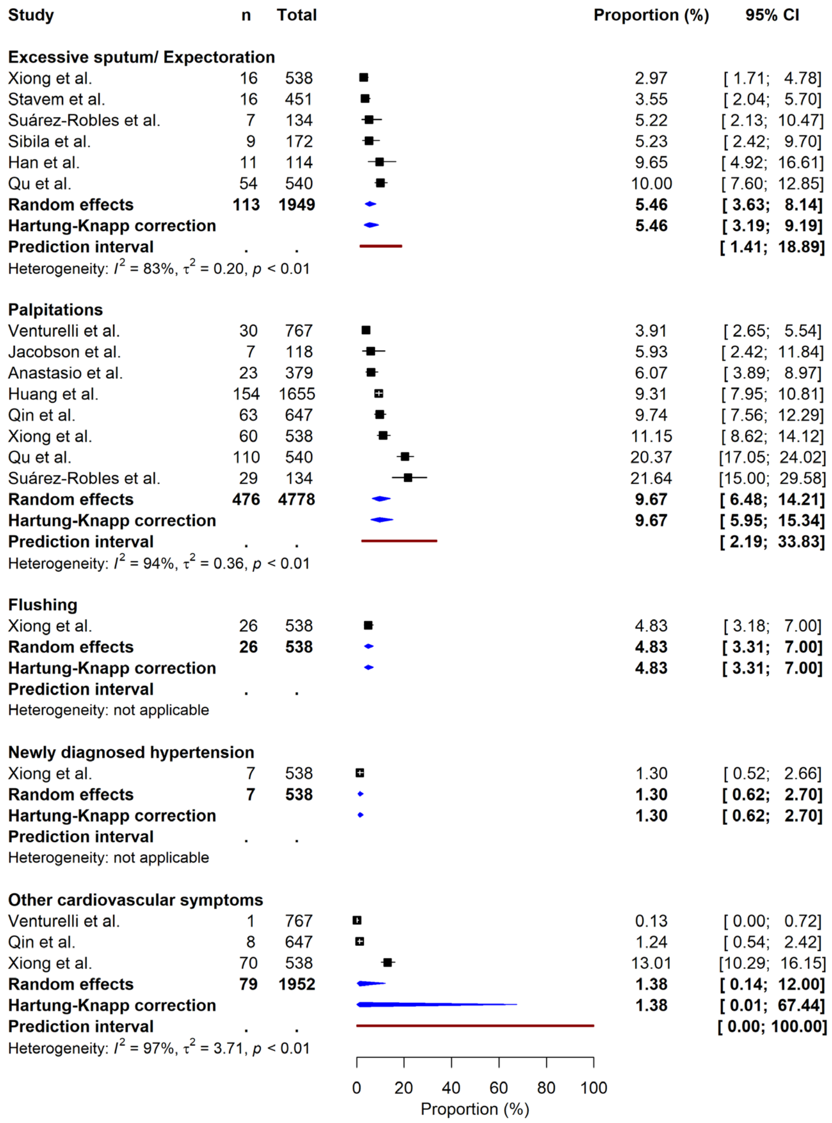

*Figure 3. Gastrointestinal*

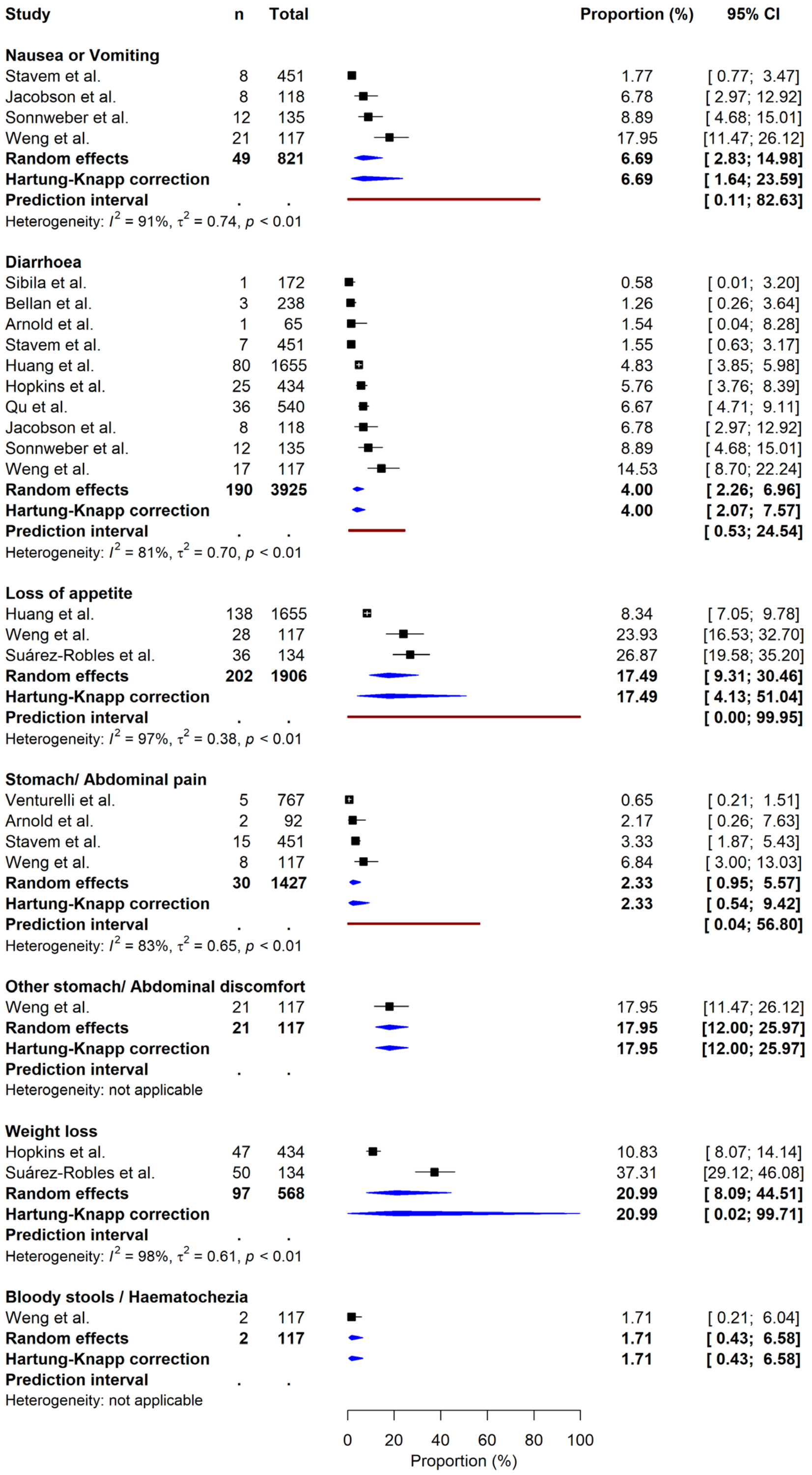

*Figure 4. Musculoskeletal*

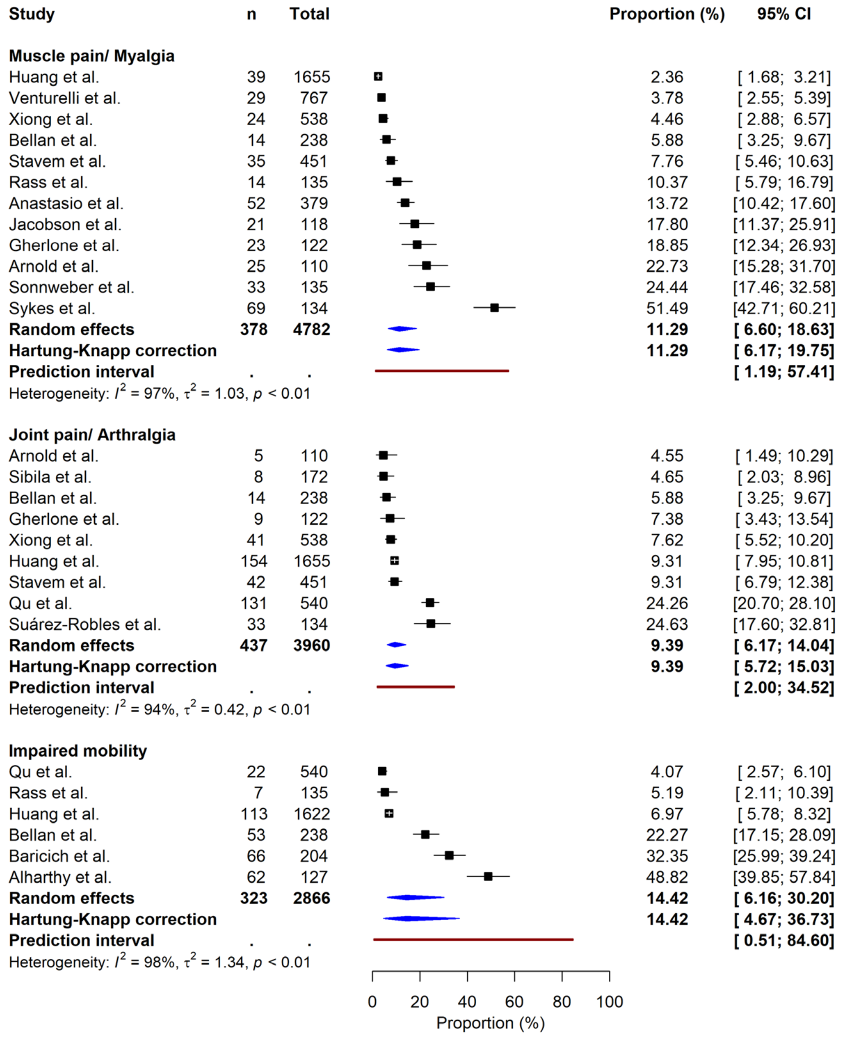

*Figure 5. Neurocognitive*

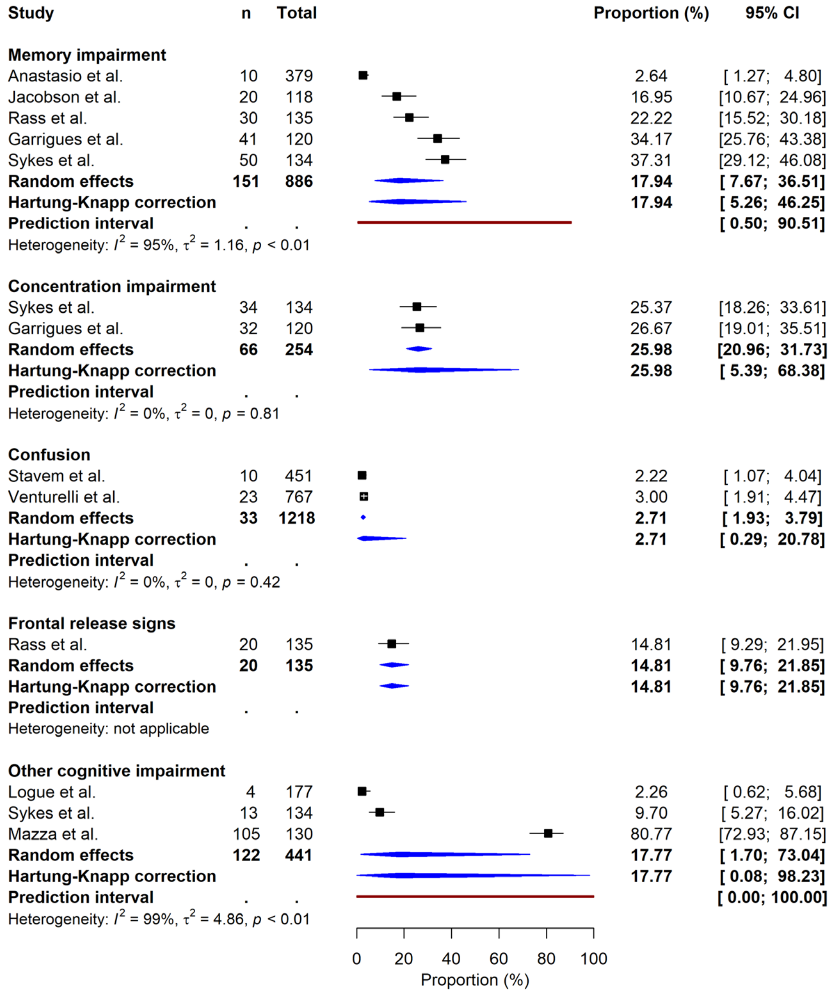

*Figure 6. Neurological and neuromuscular*

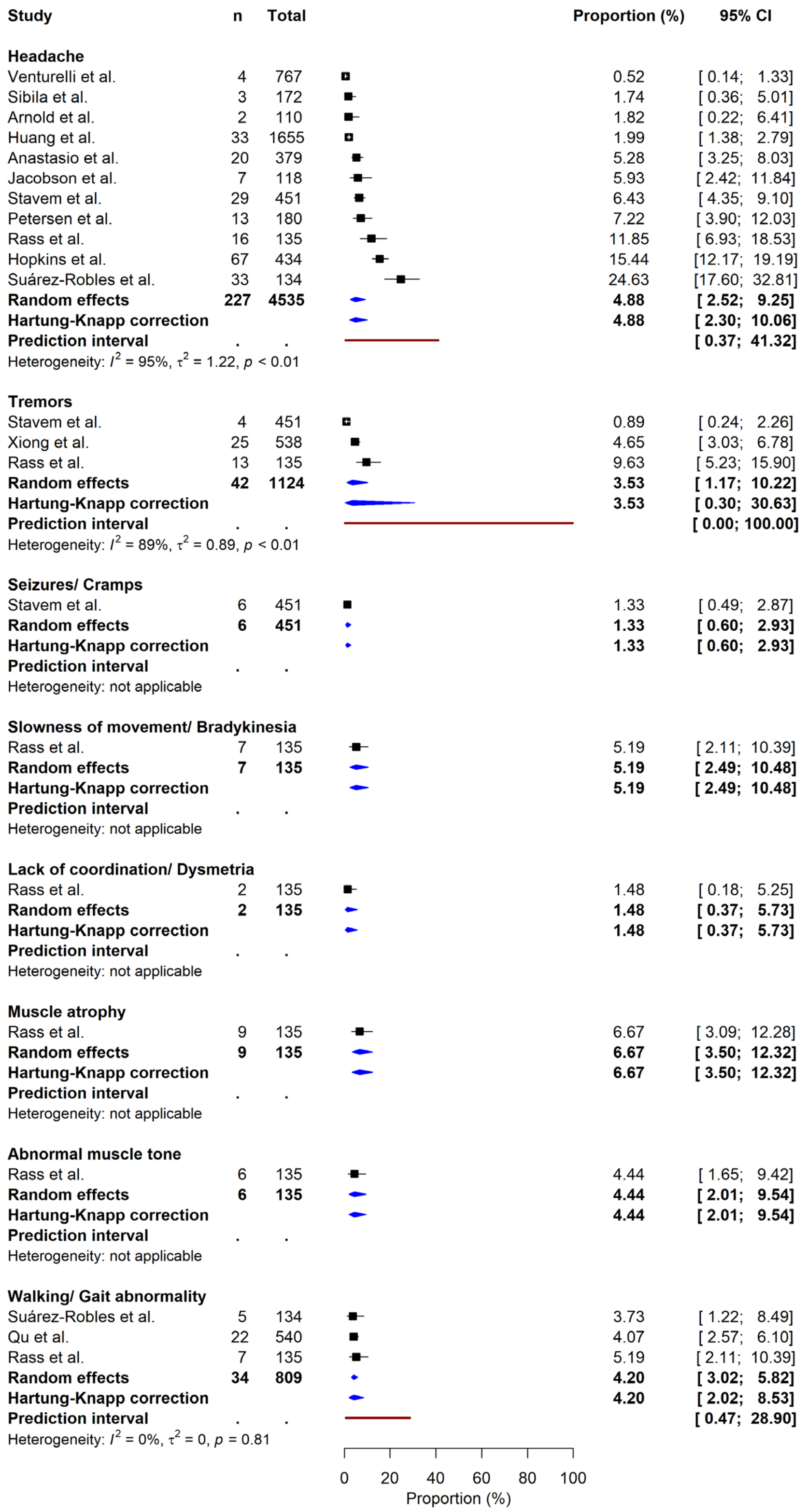

*Figure 7.Neurological and neuromuscular (page 2)*

| Study | n | Total | Proportion (%) | 95% CI |
|---|---|---|---|---|
| **Taste disturbance** | | | | |
| Venturelli et al. | 23 | 767 | 3.00 | [ 1.91; 4.47] |
| Bellan et al. | 12 | 238 | 5.04 | [ 2.63; 8.64] |
| Huang et al. | 120 | 1655 | 7.25 | [ 6.05; 8.61] |
| Klein et al. | 8 | 103 | 7.77 | [ 3.41; 14.73] |
| Sykes et al. | 12 | 134 | 8.96 | [ 4.71; 15.12] |
| Stavem et al. | 45 | 451 | 9.98 | [ 7.37; 13.12] |
| Anastasio et al. | 39 | 379 | 10.29 | [ 7.42; 13.80] |
| Garrigues et al. | 13 | 120 | 10.83 | [ 5.90; 17.81] |
| Gherlone et al. | 14 | 122 | 11.48 | [ 6.42; 18.50] |
| Logue et al. | 24 | 177 | 13.56 | [ 8.88; 19.50] |
| Rass et al. | 20 | 135 | 14.81 | [ 9.29; 21.95] |
| Petersen et al. | 29 | 180 | 16.11 | [11.06; 22.31] |
| Jacobson et al. | 25 | 118 | 21.19 | [14.20; 29.67] |
| Suárez-Robles et al. | 29 | 134 | 21.64 | [15.00; 29.58] |
| Nguyen et al. | 30 | 125 | 24.00 | [16.82; 32.46] |
| Hopkins et al. | 153 | 434 | 35.25 | [30.76; 39.95] |
| Parente-Arias et al. | 91 | 151 | 60.26 | [51.99; 68.13] |
| **Random effects** | **687** | **5423** | **13.52** | **[ 9.25; 19.34]** |
| **Hartung-Knapp correction** | | | **13.52** | **[ 8.96; 19.89]** |
| **Prediction interval** | . | . | | **[ 2.26; 51.39]** |
| Heterogeneity: $I^2$ = 97%, $\tau^2$ = 0.76, $p$ < 0.01 | | | | |
| **Smell disturbance** | | | | |
| Venturelli et al. | 23 | 767 | 3.00 | [ 1.91; 4.47] |
| Bellan et al. | 11 | 238 | 4.62 | [ 2.33; 8.12] |
| Sykes et al. | 13 | 134 | 9.70 | [ 5.27; 16.02] |
| Gherlone et al. | 12 | 122 | 9.84 | [ 5.19; 16.55] |
| Anastasio et al. | 39 | 379 | 10.29 | [ 7.42; 13.80] |
| Huang et al. | 176 | 1655 | 10.63 | [ 9.19; 12.22] |
| Arnold et al. | 13 | 110 | 11.82 | [ 6.45; 19.36] |
| Stavem et al. | 56 | 451 | 12.42 | [ 9.52; 15.82] |
| Garrigues et al. | 16 | 120 | 13.33 | [ 7.82; 20.75] |
| Logue et al. | 24 | 177 | 13.56 | [ 8.88; 19.50] |
| Klein et al. | 15 | 103 | 14.56 | [ 8.39; 22.88] |
| Rass et al. | 23 | 135 | 17.04 | [11.12; 24.46] |
| Sonnweber et al. | 26 | 135 | 19.26 | [12.98; 26.93] |
| Jacobson et al. | 25 | 118 | 21.19 | [14.20; 29.67] |
| Petersen et al. | 43 | 180 | 23.89 | [17.86; 30.80] |
| Nguyen et al. | 30 | 125 | 24.00 | [16.82; 32.46] |
| Suárez-Robles et al. | 35 | 134 | 26.12 | [18.92; 34.41] |
| Hopkins et al. | 187 | 434 | 43.09 | [38.37; 47.90] |
| Parente-Arias et al. | 75 | 151 | 49.67 | [41.44; 57.91] |
| **Random effects** | **842** | **5668** | **15.17** | **[11.01; 20.54]** |
| **Hartung-Knapp correction** | | | **15.17** | **[10.75; 20.97]** |
| **Prediction interval** | . | . | | **[ 3.13; 49.73]** |
| Heterogeneity: $I^2$ = 96%, $\tau^2$ = 0.62, $p$ < 0.01 | | | | |

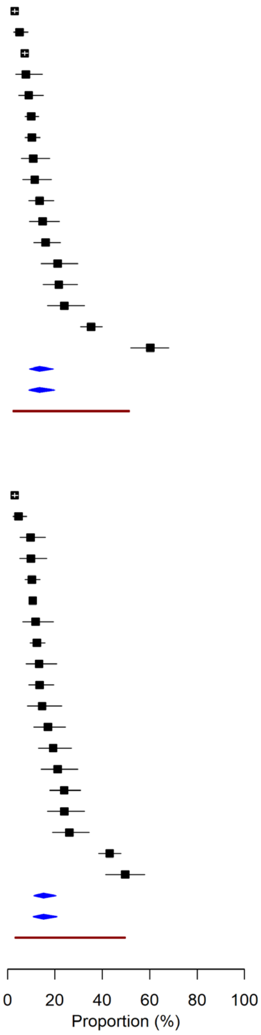

*Figure 8. Neurological and neuromuscular (page 3)*

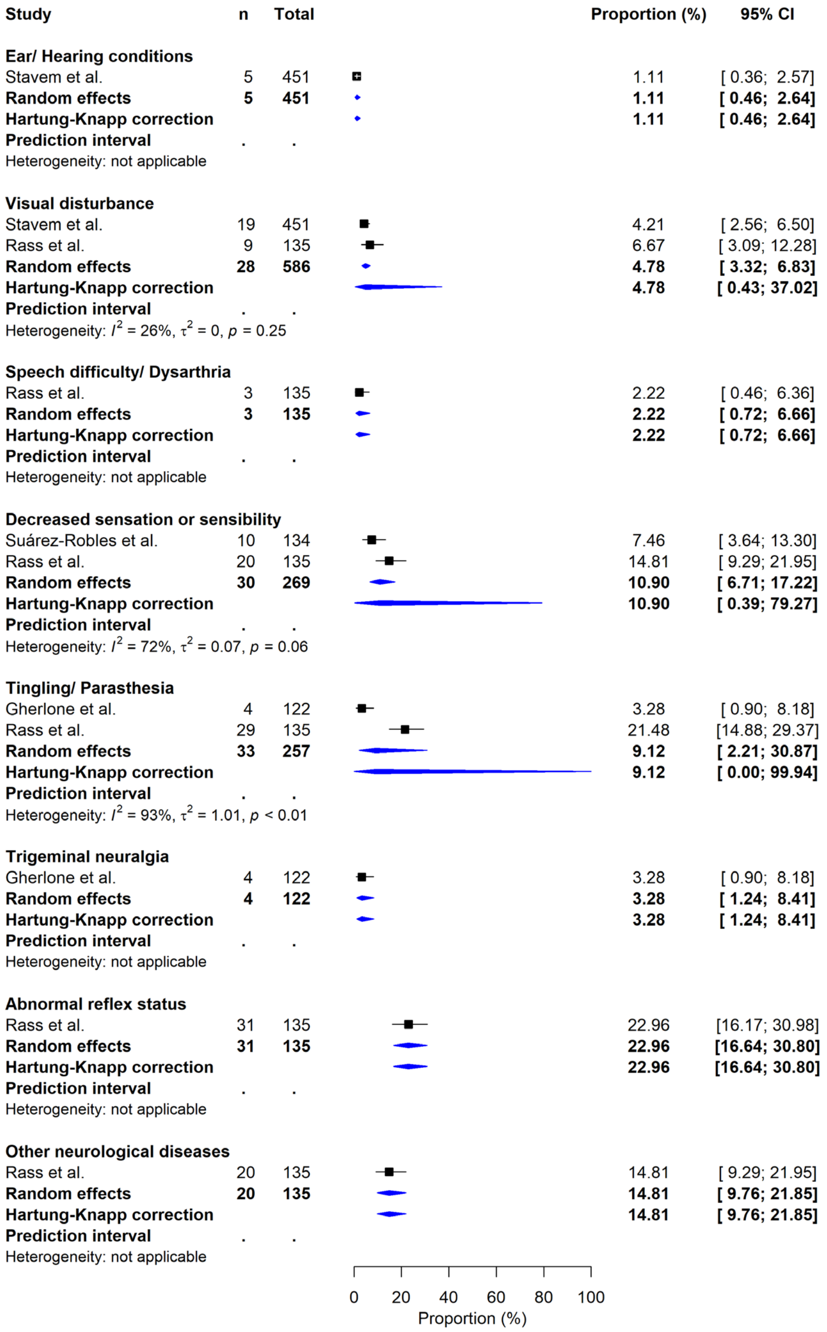

*Figure 9. Other*

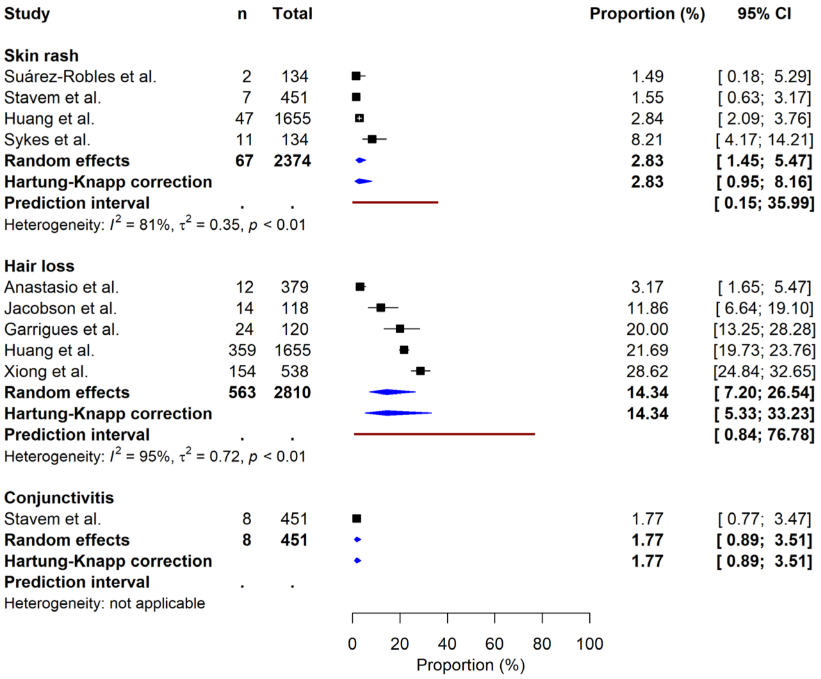

*Figure 10. Psychological and social*

| Study | n | Total | Proportion (%) | 95% CI |
|---|---|---|---|---|
| **Anxiety** | | | | |
| Xiong et al. | 35 | 538 | 6.51 | [ 4.57; 8.93] |
| Mazza et al. | 20 | 226 | 8.85 | [ 5.49; 13.34] |
| Venturelli et al. | 82 | 767 | 10.69 | [ 8.59; 13.10] |
| Rass et al. | 24 | 135 | 17.78 | [11.74; 25.29] |
| Huang et al. | 367 | 1617 | 22.70 | [20.67; 24.82] |
| Suárez-Robles et al. | 58 | 134 | 43.28 | [34.76; 52.11] |
| Sykes et al. | 64 | 134 | 47.76 | [39.07; 56.56] |
| **Random effects** | **650** | **3551** | **18.73** | **[10.37; 31.45]** |
| **Hartung-Knapp correction** | | | **18.73** | **[ 8.89; 35.25]** |
| **Prediction interval** | . | . | | **[ 1.83; 74.04]** |
| Heterogeneity: $I^2$ = 97%, $\tau^2$ = 0.83, $p$ < 0.01 | | | | |
| **Depression** | | | | |
| Xiong et al. | 23 | 538 | 4.28 | [ 2.73; 6.35] |
| Venturelli et al. | 33 | 767 | 4.30 | [ 2.98; 5.99] |
| Rass et al. | 11 | 135 | 8.15 | [ 4.14; 14.11] |
| Anastasio et al. | 31 | 379 | 8.18 | [ 5.63; 11.41] |
| Mazza et al. | 20 | 226 | 8.85 | [ 5.49; 13.34] |
| Huang et al. | 367 | 1617 | 22.70 | [20.67; 24.82] |
| **Random effects** | **485** | **3662** | **8.06** | **[ 4.86; 13.07]** |
| **Hartung-Knapp correction** | | | **8.06** | **[ 4.14; 15.10]** |
| **Prediction interval** | . | . | | **[ 1.25; 37.81]** |
| Heterogeneity: $I^2$ = 97%, $\tau^2$ = 0.41, $p$ < 0.01 | | | | |
| **Sleep disorder** | | | | |
| Mazza et al. | 7 | 226 | 3.10 | [ 1.25; 6.28] |
| Anastasio et al. | 16 | 379 | 4.22 | [ 2.43; 6.77] |
| Xiong et al. | 95 | 538 | 17.66 | [14.53; 21.15] |
| Sonnweber et al. | 32 | 145 | 22.07 | [15.61; 29.70] |
| Arnold et al. | 26 | 110 | 23.64 | [16.06; 32.68] |
| Huang et al. | 437 | 1655 | 26.40 | [24.29; 28.60] |
| Garrigues et al. | 37 | 120 | 30.83 | [22.73; 39.91] |
| Rass et al. | 45 | 135 | 33.33 | [25.46; 41.96] |
| Sykes et al. | 47 | 134 | 35.07 | [27.04; 43.79] |
| **Random effects** | **742** | **3442** | **18.15** | **[10.61; 29.30]** |
| **Hartung-Knapp correction** | | | **18.15** | **[ 9.61; 31.63]** |
| **Prediction interval** | . | . | | **[ 2.11; 69.54]** |
| Heterogeneity: $I^2$ = 94%, $\tau^2$ = 0.87, $p$ < 0.01 | | | | |
| **PTSD** | | | | |
| Mazza et al. | 5 | 226 | 2.21 | [ 0.72; 5.09] |
| Simani et al. | 7 | 120 | 5.83 | [ 2.38; 11.65] |
| Einvik et al. | 43 | 571 | 7.53 | [ 5.50; 10.01] |
| Rass et al. | 11 | 135 | 8.15 | [ 4.14; 14.11] |
| Bellan et al. | 41 | 238 | 17.23 | [12.65; 22.64] |
| Venturelli et al. | 222 | 767 | 28.94 | [25.76; 32.30] |
| **Random effects** | **329** | **2057** | **9.14** | **[ 4.57; 17.45]** |
| **Hartung-Knapp correction** | | | **9.14** | **[ 3.66; 21.04]** |
| **Prediction interval** | . | . | | **[ 0.69; 59.18]** |
| Heterogeneity: $I^2$ = 96%, $\tau^2$ = 0.78, $p$ < 0.01 | | | | |
| **Low mood/ Dysphoria** | | | | |
| Mazza et al. | 0 | 226 | 0.00 | [ 0.00; 1.62] |
| Xiong et al. | 9 | 538 | 1.67 | [ 0.77; 3.15] |
| Sykes et al. | 53 | 134 | 39.55 | [31.22; 48.36] |
| **Random effects** | **62** | **898** | **1.79** | **[ 0.04; 45.16]** |
| **Hartung-Knapp correction** | | | **1.79** | **[ 0.00; 98.74]** |
| **Prediction interval** | . | . | | **[ 0.00; 100.00]** |
| Heterogeneity: $I^2$ = 98%, $\tau^2$ = 9.19, $p$ < 0.01 | | | | |
| **Reduced quality of life** | | | | |
| Logue et al. | 53 | 177 | 29.94 | [23.30; 37.27] |
| Rass et al. | 28 | 90 | 31.11 | [21.77; 41.74] |
| Qu et al. | 259 | 540 | 47.96 | [43.68; 52.27] |
| **Random effects** | **340** | **807** | **36.76** | **[27.43; 47.21]** |
| **Hartung-Knapp correction** | | | **36.76** | **[18.43; 59.93]** |
| **Prediction interval** | . | . | | **[ 0.34; 99.01]** |
| Heterogeneity: $I^2$ = 91%, $\tau^2$ = 0.12, $p$ < 0.01 | | | | |
| **Care dependency** | | | | |
| Huang et al. | 25 | 1611 | 1.55 | [ 1.01; 2.28] |
| Logue et al. | 14 | 177 | 7.91 | [ 4.39; 12.91] |
| Venturelli et al. | 121 | 767 | 15.78 | [13.27; 18.55] |
| **Random effects** | **160** | **2555** | **5.89** | **[ 1.87; 17.05]** |
| **Hartung-Knapp correction** | | | **5.89** | **[ 0.46; 45.96]** |
| **Prediction interval** | . | . | | **[ 0.00; 100.00]** |
| Heterogeneity: $I^2$ = 98%, $\tau^2$ = 1.06, $p$ < 0.01 | | | | |

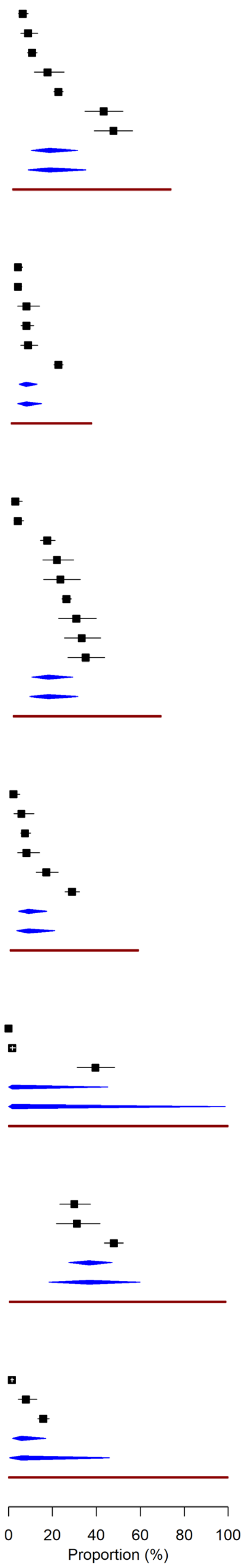

*Figure 11. Systemic*

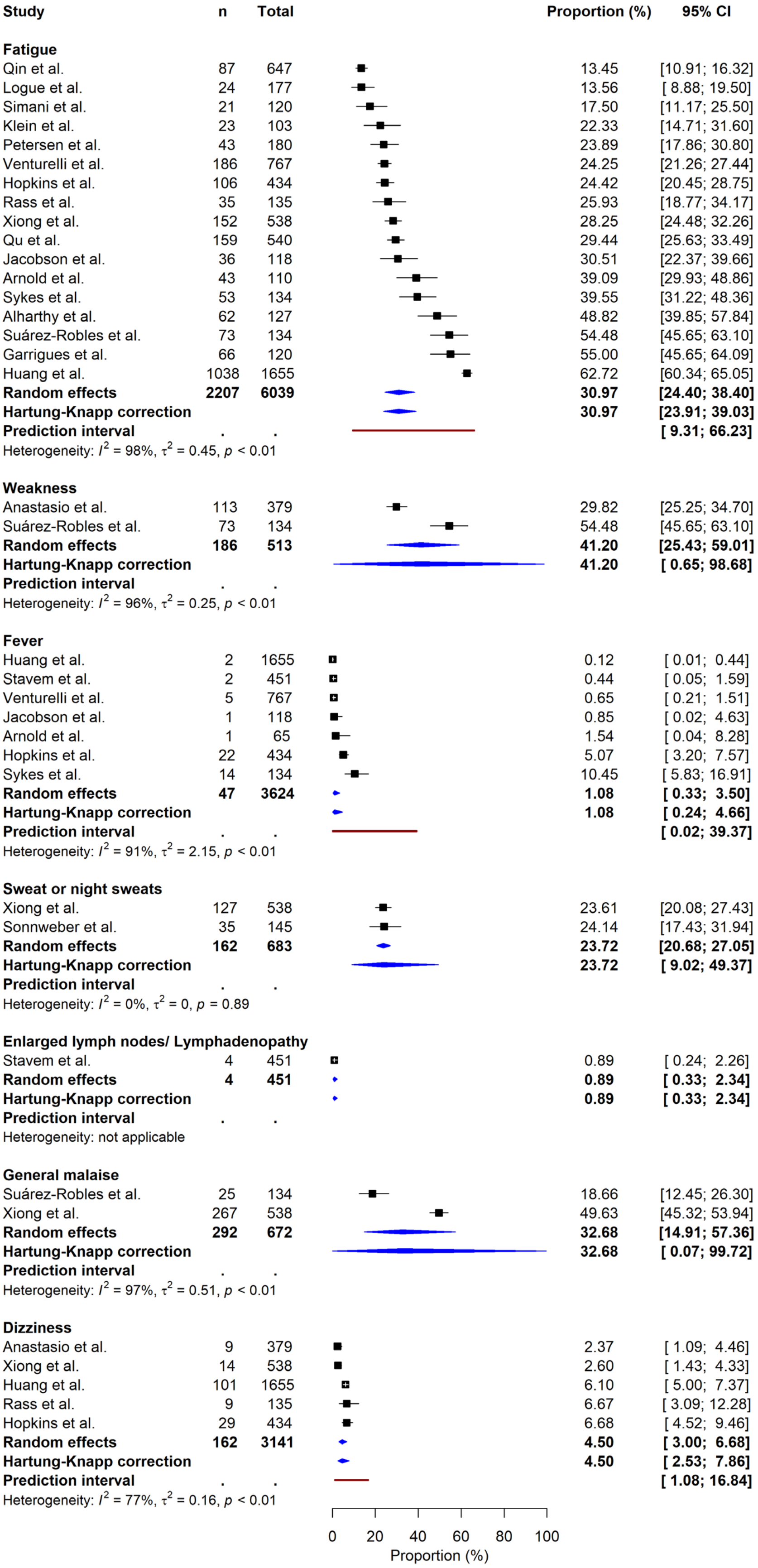

*Figure 12. Upper respiratory*

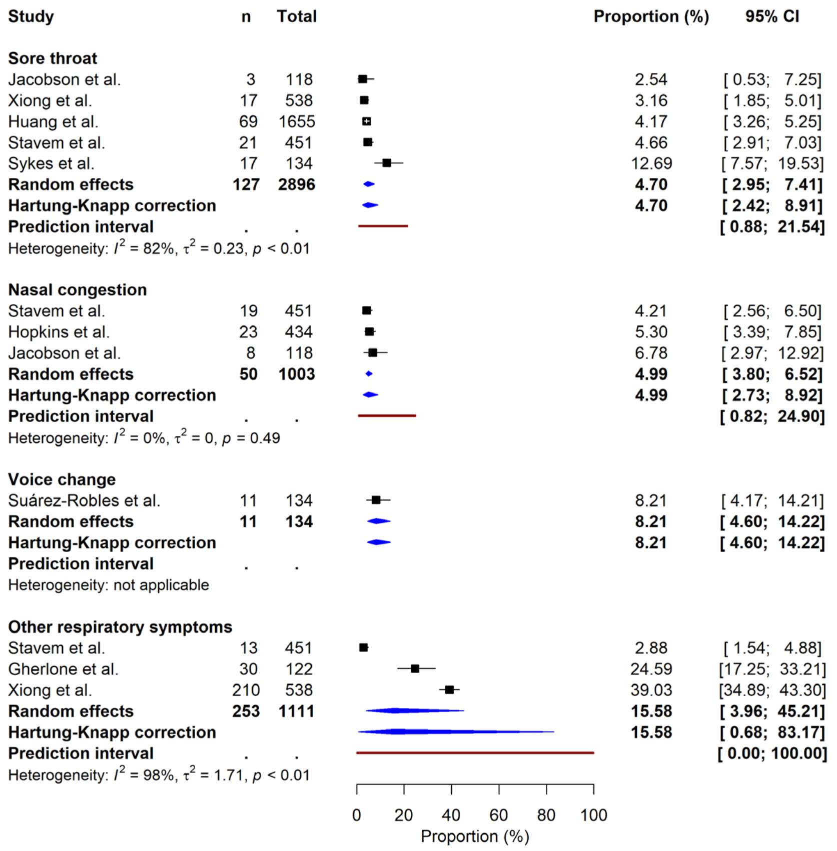

#### Supplement 5: Subgroup analysis: hospitalisation

| Classification | Symptom | Subgroup | N Studies | Event/total | Proportion% (95% CIs) | Heterogeneity (%) | P value |
|---|---|---|---|---|---|---|---|
| Cardiopulmonary | Breathlessness/ Exertional dyspnoea | Hospitalised | n = 14 | 765/3148 | 28.68 (18.48 to 41.64) | 96.19 | <0.001 |
| | | Mixed | n = 3 | 381/1291 | 32.57 (14.26 to 58.38) | 96.38 | |
| | | Non-hospitalised | n = 4 | 151/1084 | 13.72 (8.51 to 21.37) | 72.13 | |
| | Chest pain | Mixed | n = 2 | 69/1146 | 6.18 (0.01 to 97.66) | 96.65 | 0.071 |
| | | Hospitalised | n = 9 | 225/3636 | 5.92 (2.45 to 13.63) | 92.86 | |
| | | Non-hospitalised | n = 1 | 14/ 96 | 14.58 (8.83 to 23.13) | NA | |
| | Cough | Mixed | n = 3 | 54/1281 | 4.91 (0.25 to 51.82) | 96.03 | 0.265 |
| | | Hospitalised | n = 11 | 299/2769 | 10.52 (5.93 to 17.98) | 93.05 | |
| | | Non-hospitalised | n = 3 | 61/981 | 5.95 (1.53 to 20.50) | 56.24 | |
| | Excessive sputum/ Expectoration | Hospitalised | n = 5 | 97/1498 | 6.02 (3.20 to 11.03) | 82.16 | 0.112 |
| | | Non-hospitalised | n = 1 | 16/451 | 3.55 (2.18 to 5.71) | NA | |
| | Other cardiovascular symptoms | Hospitalised | n = 2 | 78/1185 | 4.20 (0.00 to 99.97) | 97.68 | 0.009 |
| | | Mixed | n = 1 | 1/767 | 0.13 (0.02 to 0.92) | NA | |
| | Palpitations | Mixed | n = 2 | 53/1146 | 4.67 (0.60 to 28.47) | 62.05 | <0.001 |
| | | Hospitalised | n = 6 | 416/3536 | 12.43 (7.78 to 19.29) | 91.7 | |
| | | Non-hospitalised | n = 1 | 7/ 96 | 7.29 (3.52 to 14.51) | NA | |
| Gastrointestinal | Diarrhoea | Hospitalised | n = 7 | 138/2809 | 2.93 (0.90 to 9.12) | 81.91 | 0.077 |
| | | Non-hospitalised | n = 3 | 40/981 | 4.16 (0.72 to 20.65) | 84.27 | |
| | | Mixed | n = 1 | 12/135 | 8.89 (5.12 to 15.00) | NA | |
| | Nausea or Vomiting | Hospitalised | n = 2 | 21/139 | 5.84 (0.00 to 100.00) | 0 | 0.343 |
| | | Non-hospitalised | n = 2 | 16/547 | 3.66 (0.00 to 98.24) | 89.91 | |
| | | Mixed | n = 1 | 12/135 | 8.89 (5.12 to 15.00) | NA | |
| | Stomach/ Abdominal pain | Hospitalised | n = 2 | 10/209 | 4.63 (0.03 to 89.20) | 54.79 | 0.002 |
| | | Non-hospitalised | n = 1 | 15/451 | 3.33 (2.01 to 5.44) | NA | |
| | | Mixed | n = 1 | 5/767 | 0.65 (0.27 to 1.56) | NA | |
| | Weight loss | Non-hospitalised | n = 1 | 47/434 | 10.83 (8.23 to 14.12) | NA | <0.001 |
| | | Hospitalised | n = 1 | 50/134 | 37.31 (29.55 to 45.79) | NA | |
| Musculoskeletal | Impaired mobility | Hospitalised | n = 5 | 316/2731 | 17.33 (4.75 to 46.83) | 98.49 | 0.038 |
| | | Mixed | n = 1 | 7/135 | 5.19 (2.49 to 10.48) | NA | |

| | | | | | | | |
|---|---|---|---|---|---|---|---|
| | Joint pain/ Arthralgia | Hospitalised | n = 8 | 395/3509 | 9.36 (5.25 to 16.14) | 94.81 | 0.987 |
| | | Non-hospitalised | n = 1 | 42/451 | 9.31 (6.95 to 12.36) | NA | |
| | Muscle pain/ Myalgia | Mixed | n = 4 | 128/1416 | 10.86 (3.45 to 29.36) | 95.21 | 0.954 |
| | | Hospitalised | n = 7 | 199/2819 | 12.46 (4.30 to 31.09) | 98.05 | |
| | | Non-hospitalised | n = 2 | 51/547 | 10.76 (0.24 to 85.64) | 85.87 | |
| Neurocognitive | Confusion | Non-hospitalised | n = 1 | 10/451 | 2.22 (1.20 to 4.07) | NA | 0.419 |
| | | Mixed | n = 1 | 23/767 | 3.00 (2.00 to 4.47) | NA | |
| | Memory impairment | Mixed | n = 2 | 40/514 | 8.06 (0.00 to 99.97) | 97.38 | <0.001 |
| | | Hospitalised | n = 3 | 96/276 | 34.78 (23.64 to 47.88) | 0 | |
| | | Non-hospitalised | n = 1 | 15/ 96 | 15.62 (9.64 to 24.32) | NA | |
| | Other cognitive impairment | Mixed | n = 2 | 109/307 | 23.55 (0.00 to 100.00) | 98.87 | 0.581 |
| | | Hospitalised | n = 1 | 13/134 | 9.70 (5.72 to 15.99) | NA | |
| Neurological and neuromuscular | Decreased sensation or sensibility | Mixed | n = 1 | 20/135 | 14.81 (9.76 to 21.85) | NA | 0.060 |
| | | Hospitalised | n = 1 | 10/134 | 7.46 (4.06 to 13.31) | NA | |
| | Headache | Mixed | n = 3 | 40/1281 | 3.30 (0.12 to 50.20) | 93.93 | 0.145 |
| | | Hospitalised | n = 5 | 71/2093 | 2.98 (0.47 to 16.53) | 96.56 | |
| | | Non-hospitalised | n = 4 | 116/1161 | 8.82 (4.41 to 16.85) | 86.25 | |
| | Smell disturbance | Mixed | n = 6 | 210/1744 | 14.63 (5.46 to 33.72) | 97.32 | 0.108 |
| | | Hospitalised | n = 9 | 308/2660 | 12.16 (7.98 to 18.10) | 85.48 | |
| | | Non-hospitalised | n = 5 | 324/1264 | 22.19 (11.69 to 38.04) | 96.3 | |
| | Taste disturbance | Mixed | n = 5 | 197/1609 | 14.50 (3.40 to 44.98) | 98.32 | 0.425 |
| | | Hospitalised | n = 8 | 232/2550 | 11.07 (6.90 to 17.28) | 89.1 | |
| | | Non-hospitalised | n = 5 | 258/1264 | 16.83 (7.91 to 32.26) | 95.66 | |
| | Tingling/ Paraesthesia | Hospitalised | n = 1 | 4/122 | 3.28 (1.24 to 8.41) | NA | <0.001 |
| | | Mixed | n = 1 | 29/135 | 21.48 (15.36 to 29.21) | NA | |
| | Tremors | Mixed | n = 1 | 13/135 | 9.63 (5.67 to 15.88) | NA | <0.001 |
| | | Non-hospitalised | n = 1 | 4/451 | 0.89 (0.33 to 2.34) | NA | |
| | | Hospitalised | n = 1 | 25/538 | 4.65 (3.16 to 6.79) | NA | |
| | Visual disturbance | Mixed | n = 1 | 9/135 | 6.67 (3.50 to 12.32) | NA | 0.245 |
| | | Non-hospitalised | n = 1 | 19/451 | 4.21 (2.70 to 6.51) | NA | |
| | Walking/ Gait abnormality | Hospitalised | n = 2 | 27/674 | 4.01 (0.34 to 33.61) | 0 | 0.534 |
| | | Mixed | n = 1 | 7/135 | 5.19 (2.49 to 10.48) | NA | |

| | | | | | | | |
|---|---|---|---|---|---|---|---|
| Other | Hair loss | Mixed | n = 1 | 12/379 | 3.17 (1.81 to 5.49) | NA | <0.001 |
| | | Hospitalised | n = 4 | 541/2335 | 23.54 (17.68 to 30.61) | 74.84 | |
| | | Non-hospitalised | n = 1 | 10/ 96 | 10.42 (5.70 to 18.29) | NA | |
| | Skin rash | Hospitalised | n = 3 | 60/1923 | 3.53 (0.75 to 15.11) | 82.97 | 0.112 |
| | | Non-hospitalised | n = 1 | 7/451 | 1.55 (0.74 to 3.22) | NA | |
| Psychological and social | Anxiety | Hospitalised | n = 4 | 524/2423 | 25.58 (6.36 to 63.49) | 97.85 | 0.072 |
| | | Mixed | n = 3 | 126/1128 | 11.60 (6.03 to 21.15) | 72 | |
| | Care dependency | Hospitalised | n = 1 | 25/1611 | 1.55 (1.05 to 2.29) | NA | <0.001 |
| | | Mixed | n = 2 | 135/944 | 12.00 (0.39 to 82.45) | 85.63 | |
| | Depression | Mixed | n = 4 | 95/1507 | 6.80 (3.99 to 11.37) | 71 | 0.506 |
| | | Hospitalised | n = 2 | 390/2155 | 10.38 (0.00 to 99.83) | 98.62 | |
| | Low mood/ Dysphoria | Mixed | n = 1 | 0/226 | 0.00 (0.00 to 100.00) | NA | 1.000 |
| | | Hospitalised | n = 2 | 62/672 | 9.49 (0.00 to 100.00) | 98.92 | |
| | PTSD | Hospitalised | n = 3 | 59/474 | 10.52 (3.06 to 30.44) | 80.04 | 0.458 |
| | | Non-hospitalised | n = 1 | 32/455 | 7.03 (5.02 to 9.78) | NA | |
| | | Mixed | n = 3 | 238/1128 | 8.73 (0.46 to 66.23) | 96.63 | |
| | Reduced quality of life | Mixed | n = 2 | 81/267 | 30.34 (7.43 to 70.27) | 0 | <0.001 |
| | | Hospitalised | n = 1 | 259/540 | 47.96 (43.77 to 52.18) | NA | |
| | Sleep disorder | Mixed | n = 4 | 100/885 | 10.66 (1.76 to 44.22) | 96.51 | 0.081 |
| | | Hospitalised | n = 5 | 642/2557 | 25.81 (18.85 to 34.26) | 84.7 | |
| Systemic | Dizziness | Mixed | n = 2 | 18/514 | 3.78 (0.03 to 83.74) | 79.93 | 0.224 |
| | | Non-hospitalised | n = 1 | 29/434 | 6.68 (4.68 to 9.45) | NA | |
| | | Hospitalised | n = 2 | 115/2193 | 4.21 (0.08 to 71.53) | 89.39 | |
| | Fatigue | Hospitalised | n = 11 | 1762/4147 | 37.10 (26.54 to 49.06) | 98.23 | 0.017 |
| | | Non-hospitalised | n = 4 | 200/813 | 24.60 (20.11 to 29.72) | 0 | |
| | | Mixed | n = 3 | 245/1079 | 21.04 (10.48 to 37.75) | 79.86 | |
| | Fever | Hospitalised | n = 4 | 17/1876 | 0.85 (0.02 to 24.20) | 92.05 | 0.661 |
| | | Non-hospitalised | n = 3 | 25/981 | 1.41 (0.06 to 24.82) | 84.73 | |
| | | Mixed | n = 1 | 5/767 | 0.65 (0.27 to 1.56) | NA | |
| | Sweat or night sweats | Mixed | n = 1 | 35/145 | 24.14 (17.87 to 31.76) | NA | 0.894 |
| | | Hospitalised | n = 1 | 127/538 | 23.61 (20.21 to 27.38) | NA | |
| | Weakness | Mixed | n = 1 | 113/379 | 29.82 (25.42 to 34.61) | NA | <0.001 |

| | | | | | | | |
|---|---|---|---|---|---|---|---|
| | | Hospitalised | n = 1 | 73/134 | 54.48 (46.00 to 62.70) | NA | |
| Upper respiratory | Nasal congestion | Non-hospitalised | n = 3 | 49/981 | 4.99 (2.72 to 8.99) | 0 | 0.924 |
| | | Hospitalised | n = 1 | 1/ 22 | 4.55 (0.64 to 26.15) | NA | |
| | Other respiratory symptoms | Hospitalised | n = 2 | 240/660 | 32.43 (2.22 to 91.02) | 88.57 | <0.001 |
| | | Non-hospitalised | n = 1 | 13/451 | 2.88 (1.68 to 4.90) | NA | |
| | Sore throat | Hospitalised | n = 4 | 103/2349 | 4.81 (1.60 to 13.60) | 85.83 | 0.815 |
| | | Non-hospitalised | n = 2 | 24/547 | 4.39 (0.32 to 39.44) | 0 | |

#### Supplement 6: Subgroup analysis: Setting

| Classification | Symptom | Subgroup | N Studies | Event/total | Proportion% (95% CIs) | Heterogeneity (%) | P value |
|---|---|---|---|---|---|---|---|
| Cardiopulmonary | Cough | Multicentre | n = 5 | 121/1358 | 6.90 (2.46 to 17.92) | 84.05 | 0.842 |
| | | Single-centre | n = 10 | 260/3239 | 9.07 (4.23 to 18.41) | 95.62 | |
| | | Online survey | n = 1 | 33/ 434 | 7.60 (5.46 to 10.50) | NA | |
| | Chest pain | Single-centre | n = 10 | 292/4760 | 5.87 (2.70 to 12.26) | 93.7 | 0.039 |
| | | Multicentre | n = 1 | 16/ 118 | 13.56 (8.48 to 20.99) | NA | |
| | Breathlessness/ Exertional dyspnoea | Multicentre | n = 6 | 350/1437 | 26.79 (15.81 to 41.63) | 91.82 | <0.001 |
| | | Single-centre | n = 12 | 889/3549 | 27.78 (17.16 to 41.67) | 96.93 | |
| | | Online survey | n = 2 | 58/ 537 | 10.80 (2.03 to 41.47) | 16.75 | |
| | Palpitations | Single-centre | n = 6 | 359/4120 | 9.02 (5.17 to 15.27) | 90.72 | 0.571 |
| | | Multicentre | n = 2 | 117/ 658 | 11.96 (0.02 to 98.84) | 91.67 | |
| | Excessive sputum/ Expectoration | Multicentre | n = 3 | 81/1105 | 6.97 (2.02 to 21.38) | 86.45 | 0.066 |
| | | Single-centre | n = 3 | 32/ 844 | 3.79 (1.78 to 7.88) | 24.53 | |
| Gastrointestinal | Weight loss | Online survey | n = 1 | 47/ 434 | 10.83 (8.23 to 14.12) | NA | <0.001 |
| | | Single-centre | n = 1 | 50/ 134 | 37.31 (29.55 to 45.79) | NA | |
| | Stomach/ Abdominal pain | Single-centre | n = 2 | 7/ 859 | 0.81 (0.01 to 50.51) | 52.12 | <0.001 |
| | | Multicentre | n = 2 | 23/ 568 | 4.05 (0.28 to 38.69) | 64.62 | |
| | Loss of appetite | Single-centre | n = 2 | 174/1789 | 15.09 (0.03 to 98.93) | 97.64 | 0.288 |
| | | Multicentre | n = 1 | 28/ 117 | 23.93 (17.06 to 32.48) | NA | |
| | Diarrhoea | Single-centre | n = 4 | 85/2130 | 1.81 (0.36 to 8.72) | 72.7 | 0.081 |
| | | Multicentre | n = 5 | 80/1361 | 6.23 (2.49 to 14.76) | 85.33 | |
| | | Online survey | n = 1 | 25/ 434 | 5.76 (3.92 to 8.39) | NA | |
| Musculoskeletal | Impaired mobility | Single-centre | n = 4 | 294/2191 | 23.71 (6.53 to 58.01) | 98.62 | <0.001 |
| | | Multicentre | n = 2 | 29/ 675 | 4.30 (0.40 to 33.37) | 0 | |
| | Joint pain/ Arthralgia | Single-centre | n = 7 | 264/2969 | 8.03 (4.64 to 13.55) | 86.94 | 0.116 |
| | | Multicentre | n = 2 | 173/ 991 | 15.45 (0.11 to 96.89) | 97.19 | |
| | Muscle pain/ Myalgia | Multicentre | n = 4 | 103/ 839 | 13.72 (6.26 to 27.48) | 89.49 | 0.508 |
| | | Single-centre | n = 8 | 275/3943 | 10.23 (4.03 to 23.63) | 97.97 | |
| Neurocognitive | Other cognitive impairment | Single-centre | n = 2 | 118/ 264 | 40.19 (0.00 to 100.00) | 99 | 0.017 |
| | | Multicentre | n = 1 | 4/ 177 | 2.26 (0.85 to 5.86) | NA | |

| | | | | | | | |
|---|---|---|---|---|---|---|---|
| | Confusion | Multicentre | n = 1 | 10/ 451 | 2.22 (1.20 to 4.07) | NA | 0.419 |
| | | Single-centre | n = 1 | 23/ 767 | 3.00 (2.00 to 4.47) | NA | |
| | Memory impairment | Multicentre | n = 2 | 50/ 253 | 19.76 (3.21 to 64.68) | 8.97 | 0.821 |
| | | Single-centre | n = 3 | 101/ 633 | 16.93 (0.58 to 87.62) | 97.39 | |
| Neurological and neuromuscular | Walking/ Gait abnormality | Multicentre | n = 2 | 29/ 675 | 4.30 (0.40 to 33.37) | 0 | 0.766 |
| | | Single-centre | n = 1 | 5/ 134 | 3.73 (1.56 to 8.65) | NA | |
| | Tremors | Multicentre | n = 2 | 17/ 586 | 2.98 (0.00 to 99.96) | 94.5 | 0.615 |
| | | Single-centre | n = 1 | 25/ 538 | 4.65 (3.16 to 6.79) | NA | |
| | Headache | Single-centre | n = 6 | 95/3217 | 2.82 (0.69 to 10.88) | 96.4 | <0.001 |
| | | Multicentre | n = 4 | 65/ 884 | 7.35 (5.00 to 10.68) | 37.26 | |
| | | Online survey | n = 1 | 67/ 434 | 15.44 (12.34 to 19.15) | NA | |
| | Smell disturbance | Multicentre | n = 6 | 197/1196 | 17.21 (13.03 to 22.38) | 68.78 | 0.235 |
| | | Single-centre | n = 11 | 443/3935 | 12.49 (7.13 to 20.97) | 95.97 | |
| | | Online survey | n = 2 | 202/ 537 | 27.06 (0.04 to 99.70) | 96.07 | |
| | Taste disturbance | Single-centre | n = 10 | 383/3825 | 12.21 (6.16 to 22.76) | 97.3 | 0.793 |
| | | Multicentre | n = 5 | 143/1061 | 14.27 (10.13 to 19.73) | 65.8 | |
| | | Online survey | n = 2 | 161/ 537 | 18.21 (0.00 to 99.91) | 95.82 | |
| | Tingling/ Paraesthesia | Single-centre | n = 1 | 4/ 122 | 3.28 (1.24 to 8.41) | NA | <0.001 |
| | | Multicentre | n = 1 | 29/ 135 | 21.48 (15.36 to 29.21) | NA | |
| | Decreased sensation or sensibility | Multicentre | n = 1 | 20/ 135 | 14.81 (9.76 to 21.85) | NA | 0.060 |
| | | Single-centre | n = 1 | 10/ 134 | 7.46 (4.06 to 13.31) | NA | |
| Other | Hair loss | Multicentre | n = 1 | 14/ 118 | 11.86 (7.15 to 19.04) | NA | 0.630 |
| | | Single-centre | n = 4 | 549/2692 | 14.99 (3.66 to 45.01) | 95.58 | |
| | Skin rash | Single-centre | n = 3 | 60/1923 | 3.53 (0.75 to 15.11) | 82.97 | 0.112 |
| | | Multicentre | n = 1 | 7/ 451 | 1.55 (0.74 to 3.22) | NA | |
| Psychological and social | Care dependency | Single-centre | n = 2 | 146/2378 | 5.16 (0.00 to 99.97) | 99.18 | 0.621 |
| | | Multicentre | n = 1 | 14/ 177 | 7.91 (4.74 to 12.91) | NA | |
| | PTSD | Multicentre | n = 2 | 54/ 706 | 7.65 (1.35 to 33.36) | 0 | 0.653 |
| | | Single-centre | n = 4 | 275/1351 | 9.73 (1.74 to 39.56) | 95.56 | |
| | Sleep disorder | Single-centre | n = 7 | 665/3162 | 15.96 (6.78 to 33.15) | 95.15 | 0.119 |
| | | Multicentre | n = 2 | 77/ 280 | 27.41 (2.84 to 82.98) | 77.28 | |
| | Depression | Single-centre | n = 5 | 474/3527 | 8.06 (3.47 to 17.62) | 97.89 | 0.979 |

| | | | | | | | |
|---|---|---|---|---|---|---|---|
| | | Multicentre | n = 1 | 11/ 135 | 8.15 (4.57 to 14.11) | NA | |
| | Anxiety | Single-centre | n = 6 | 626/3416 | 18.92 (7.55 to 40.00) | 97.66 | 0.870 |
| | | Multicentre | n = 1 | 24/ 135 | 17.78 (12.21 to 25.16) | NA | |
| Systemic | Dizziness | Single-centre | n = 3 | 124/2572 | 3.55 (1.05 to 11.30) | 87.17 | 0.138 |
| | | Online survey | n = 1 | 29/ 434 | 6.68 (4.68 to 9.45) | NA | |
| | | Multicentre | n = 1 | 9/ 135 | 6.67 (3.50 to 12.32) | NA | |
| | Sweat or night sweats | Multicentre | n = 1 | 35/ 145 | 24.14 (17.87 to 31.76) | NA | 0.894 |
| | | Single-centre | n = 1 | 127/ 538 | 23.61 (20.21 to 27.38) | NA | |
| | Fever | Single-centre | n = 4 | 22/2621 | 0.98 (0.06 to 14.94) | 94.58 | <0.001 |
| | | Online survey | n = 1 | 22/ 434 | 5.07 (3.36 to 7.58) | NA | |
| | | Multicentre | n = 2 | 3/ 569 | 0.53 (0.00 to 89.24) | 0 | |
| | Fatigue | Single-centre | n = 10 | 1781/4352 | 36.55 (25.00 to 49.88) | 98.57 | 0.067 |
| | | Multicentre | n = 5 | 297/1150 | 24.28 (17.14 to 33.19) | 78.24 | |
| | | Online survey | n = 2 | 129/ 537 | 24.02 (8.05 to 53.30) | 0 | |
| Upper respiratory | Other respiratory symptoms | Single-centre | n = 2 | 240/ 660 | 32.43 (2.22 to 91.02) | 88.57 | <0.001 |
| | | Multicentre | n = 1 | 13/ 451 | 2.88 (1.68 to 4.90) | NA | |
| | Nasal congestion | Online survey | n = 1 | 23/ 434 | 5.30 (3.55 to 7.85) | NA | 0.690 |
| | | Multicentre | n = 2 | 27/ 569 | 4.75 (0.41 to 37.90) | 25.27 | |
| | Sore throat | Single-centre | n = 3 | 103/2327 | 5.38 (1.20 to 20.94) | 90.55 | 0.538 |
| | | Multicentre | n = 2 | 24/ 569 | 4.22 (0.31 to 38.40) | 0.31 | |

#### Supplement 7: Subgroup analysis: Continents

| Classification | Symptom | Subgroup | N Studies | Event/total | Proportion% (95% CIs) | Heterogeneity (%) | P value |
|---|---|---|---|---|---|---|---|
| Cardiopulmonary | Cough | Europe | n = 11 | 267/3074 | 9.71 (4.88 to 18.39) | 94.92 | 0.057 |
| | | Asia | n = 4 | 146/1839 | 7.64 (4.60 to 12.44) | 79.54 | |
| | | North America | n = 1 | 1/ 118 | 0.85 (0.12 to 5.77) | NA | |
| | Chest pain | Asia | n = 3 | 157/2840 | 4.11 (0.23 to 44.11) | 97.52 | 0.097 |
| | | Europe | n = 7 | 135/1920 | 6.93 (2.77 to 16.31) | 89.56 | |
| | | North America | n = 1 | 16/ 118 | 13.56 (8.48 to 20.99) | NA | |
| | Breathlessness/ Exertional dyspnoea | Europe | n = 13 | 868/3336 | 28.59 (18.52 to 41.35) | 96.32 | 0.227 |
| | | Asia | n = 4 | 328/1839 | 16.53 (7.91 to 31.34) | 95.25 | |
| | | Middle East | n = 2 | 70/ 230 | 22.35 (0.00 to 99.99) | 97.16 | |
| | | North America | n = 1 | 31/ 118 | 26.27 (19.13 to 34.93) | NA | |
| | Other cardiovascular symptoms | Europe | n = 1 | 1/ 767 | 0.13 (0.02 to 0.92) | NA | 0.009 |
| | | Asia | n = 2 | 78/1185 | 4.20 (0.00 to 99.97) | 97.68 | |
| | Palpitations | Asia | n = 4 | 387/3380 | 12.04 (7.03 to 19.85) | 93.93 | 0.168 |
| | | Europe | n = 3 | 82/1280 | 8.10 (1.14 to 40.21) | 95.9 | |
| | | North America | n = 1 | 7/ 118 | 5.93 (2.85 to 11.92) | NA | |
| | Excessive sputum/ Expectoration | Asia | n = 3 | 81/1192 | 6.56 (1.54 to 23.96) | 90.08 | 0.238 |
| | | Europe | n = 3 | 32/ 757 | 4.23 (1.99 to 8.76) | 0 | |
| Gastrointestinal | Stomach/ Abdominal pain | Europe | n = 3 | 22/1310 | 1.61 (0.22 to 10.84) | 80.31 | 0.011 |
| | | Asia | n = 1 | 8/ 117 | 6.84 (3.46 to 13.08) | NA | |
| | Loss of appetite | Asia | n = 2 | 166/1772 | 13.98 (0.06 to 97.66) | 96.44 | 0.088 |
| | | Europe | n = 1 | 36/ 134 | 26.87 (20.05 to 34.99) | NA | |
| | Diarrhoea | Europe | n = 6 | 49/1495 | 2.37 (0.80 to 6.77) | 80.93 | 0.055 |
| | | Asia | n = 3 | 133/2312 | 7.38 (2.34 to 20.91) | 89.09 | |
| | | North America | n = 1 | 8/ 118 | 6.78 (3.43 to 12.97) | NA | |
| | Nausea or Vomiting | Europe | n = 2 | 20/ 586 | 3.92 (0.00 to 98.84) | 92.31 | 0.004 |
| | | Asia | n = 1 | 21/ 117 | 17.95 (12.00 to 25.97) | NA | |
| | | North America | n = 1 | 8/ 118 | 6.78 (3.43 to 12.97) | NA | |
| Musculoskeletal | Impaired mobility | Asia | n = 2 | 135/2162 | 5.63 (0.46 to 43.57) | 82.31 | <0.001 |
| | | Europe | n = 3 | 126/ 577 | 16.90 (2.10 to 65.88) | 92.96 | |

| | | | | | | | |
|---|---|---|---|---|---|---|---|
| | | Middle East | n = 1 | 62/ 127 | 48.82 (40.25 to 57.46) | NA | |
| | Joint pain/ Arthralgia | Asia | n = 3 | 326/2733 | 12.25 (3.07 to 38.13) | 97.8 | 0.295 |
| | | Europe | n = 6 | 111/1227 | 8.03 (4.01 to 15.42) | 88.36 | |
| | Muscle pain/ Myalgia | Europe | n = 9 | 294/2471 | 14.02 (7.27 to 25.34) | 96.38 | <0.001 |
| | | Asia | n = 2 | 63/2193 | 3.11 (0.16 to 38.54) | 83.96 | |
| | | North America | n = 1 | 21/ 118 | 17.80 (11.90 to 25.76) | NA | |
| Neurocognitive | Other cognitive impairment | Europe | n = 2 | 118/ 264 | 40.19 (0.00 to 100.00) | 99 | 0.017 |
| | | North America | n = 1 | 4/ 177 | 2.26 (0.85 to 5.86) | NA | |
| | Memory impairment | North America | n = 1 | 20/ 118 | 16.95 (11.20 to 24.82) | NA | 0.898 |
| | | Europe | n = 4 | 131/ 768 | 18.19 (3.02 to 61.38) | 96.14 | |
| Neurological and neuromuscular | Walking/ Gait abnormality | Europe | n = 2 | 12/ 269 | 4.46 (0.11 to 66.56) | 0 | 0.796 |
| | | Asia | n = 1 | 22/ 540 | 4.07 (2.70 to 6.11) | NA | |
| | Tremors | Europe | n = 2 | 17/ 586 | 2.98 (0.00 to 99.96) | 94.5 | 0.615 |
| | | Asia | n = 1 | 25/ 538 | 4.65 (3.16 to 6.79) | NA | |
| | Headache | Asia | n = 1 | 33/1655 | 1.99 (1.42 to 2.79) | NA | 0.005 |
| | | Europe | n = 9 | 187/2762 | 5.30 (2.12 to 12.66) | 92.94 | |
| | | North America | n = 1 | 7/ 118 | 5.93 (2.85 to 11.92) | NA | |
| | Smell disturbance | Europe | n = 15 | 602/3615 | 15.35 (9.88 to 23.06) | 96.43 | 0.027 |
| | | Asia | n = 1 | 176/1655 | 10.63 (9.24 to 12.21) | NA | |
| | | North America | n = 2 | 49/ 295 | 16.74 (1.75 to 69.42) | 65.9 | |
| | | Middle East | n = 1 | 15/ 103 | 14.56 (8.97 to 22.76) | NA | |
| | Taste disturbance | Asia | n = 1 | 120/1655 | 7.25 (6.10 to 8.60) | NA | <0.001 |
| | | Europe | n = 13 | 510/3370 | 14.25 (8.44 to 23.06) | 96.81 | |
| | | North America | n = 2 | 49/ 295 | 16.74 (1.75 to 69.42) | 65.9 | |
| | | Middle East | n = 1 | 8/ 103 | 7.77 (3.93 to 14.77) | NA | |
| Other | Hair loss | North America | n = 1 | 14/ 118 | 11.86 (7.15 to 19.04) | NA | 0.005 |
| | | Asia | n = 2 | 513/2193 | 24.69 (5.86 to 63.32) | 90.76 | |
| | | Europe | n = 2 | 36/ 499 | 8.21 (0.00 to 99.89) | 96.66 | |
| | Skin rash | Asia | n = 1 | 47/1655 | 2.84 (2.14 to 3.76) | NA | 0.952 |
| | | Europe | n = 3 | 20/ 719 | 2.75 (0.30 to 20.89) | 86.08 | |
| Psychosological and social | Care dependency | Asia | n = 1 | 25/1611 | 1.55 (1.05 to 2.29) | NA | <0.001 |
| | | North America | n = 1 | 14/ 177 | 7.91 (4.74 to 12.91) | NA | |

| | | | | | | | |
|---|---|---|---|---|---|---|---|
| | | Europe | n = 1 | 121/ 767 | 15.78 (13.36 to 18.53) | NA | |
| | Reduced quality of life | Europe | n = 1 | 28/90 | 31.11 (22.42 to 41.37) | NA | <0.001 |
| | | North America | n = 1 | 53/ 177 | 29.94 (23.66 to 37.09) | NA | |
| | | Asia | n = 1 | 259/ 540 | 47.96 (43.77 to 52.18) | NA | |
| | Low mood/ Dysphoria | Europe | n = 2 | 53/ 360 | 0.86 (0.00 to 100.00) | 0 | 0.868 |
| | | Asia | n = 1 | 9/ 538 | 1.67 (0.87 to 3.18) | NA | |
| | PTSD | Europe | n = 5 | 322/1937 | 9.93 (3.21 to 26.84) | 96.87 | 0.322 |
| | | Middle East | n = 1 | 7/ 120 | 5.83 (2.81 to 11.73) | NA | |
| | Sleep disorder | Asia | n = 2 | 532/2193 | 22.00 (2.69 to 74.18) | 94 | 0.486 |
| | | Europe | n = 7 | 210/1249 | 17.09 (7.01 to 36.03) | 94.68 | |
| | Depression | Asia | n = 2 | 390/2155 | 10.38 (0.00 to 99.83) | 98.62 | 0.506 |
| | | Europe | n = 4 | 95/1507 | 6.80 (3.99 to 11.37) | 71 | |
| | Anxiety | Asia | n = 2 | 402/2155 | 12.63 (0.02 to 98.99) | 98.36 | 0.320 |
| | | Europe | n = 5 | 248/1396 | 21.85 (8.03 to 47.22) | 97.39 | |
| Systemic | Dizziness | Asia | n = 2 | 115/2193 | 4.21 (0.08 to 71.53) | 89.39 | 0.764 |
| | | Europe | n = 3 | 47/ 948 | 4.76 (1.43 to 14.71) | 75.61 | |
| | General malaise | Europe | n = 1 | 25/ 134 | 18.66 (12.93 to 26.16) | NA | <0.001 |
| | | Asia | n = 1 | 267/ 538 | 49.63 (45.42 to 53.85) | NA | |
| | Sweat or night sweats | Europe | n = 1 | 35/ 145 | 24.14 (17.87 to 31.76) | NA | 0.894 |
| | | Asia | n = 1 | 127/ 538 | 23.61 (20.21 to 27.38) | NA | |
| | Fever | Europe | n = 5 | 44/1851 | 1.91 (0.36 to 9.62) | 90.51 | 0.011 |
| | | Asia | n = 1 | 2/1655 | 0.12 (0.03 to 0.48) | NA | |
| | | North America | n = 1 | 1/ 118 | 0.85 (0.12 to 5.77) | NA | |
| | Fatigue | Europe | n = 8 | 605/2014 | 34.68 (25.12 to 45.66) | 93 | 0.382 |
| | | North America | n = 2 | 60/ 295 | 20.71 (0.25 to 96.48) | 91.68 | |
| | | Asia | n = 4 | 1436/3380 | 31.33 (10.49 to 63.98) | 99.42 | |
| | | Middle East | n = 3 | 106/ 350 | 28.07 (6.94 to 67.15) | 93.59 | |
| Upper respiratory | Other respiratory symptoms | Europe | n = 2 | 43/ 573 | 8.88 (0.00 to 99.98) | 97.85 | 0.029 |
| | | Asia | n = 1 | 210/ 538 | 39.03 (35.00 to 43.22) | NA | |
| | Nasal congestion | Europe | n = 2 | 42/ 885 | 4.75 (0.66 to 27.08) | 0 | 0.343 |
| | | North America | n = 1 | 8/ 118 | 6.78 (3.43 to 12.97) | NA | |
| | Sore throat | Asia | n = 2 | 86/2193 | 3.92 (1.00 to 14.17) | 8.33 | 0.165 |

| | | | | | |
|---|---|---|---|---|---|
| | Europe | n = 2 | 38/ 585 | 7.48 (0.06 to 91.61) | 90.13 |
| | North America | n = 1 | 3/ 118 | 2.54 (0.82 to 7.59) | NA |

#### Supplement 8: Subgroup analysis: Follow-up timing

| Classification | Symptom | Subgroup | N Studies | Event/total | Proportion% (95% CIs) | Heterogeneity (%) | P value |
|---|---|---|---|---|---|---|---|
| Cardiopulmonary | Cough | < 4 months | n = 14 | 374/4483 | 8.35 (4.56 to 14.81) | 94.34 | 0.669 |
| | | > 4 months | n = 2 | 40/548 | 7.30 (0.97 to 38.82) | 0 | |
| | Chest pain | > 4 months | n = 1 | 75/1655 | 4.53 (3.63 to 5.65) | NA | 0.311 |
| | | < 4 months | n = 10 | 233/3223 | 6.55 (2.97 to 13.84) | 91.52 | |
| | Breathlessness/ Exertional dyspnoea | < 4 months | n = 15 | 1142/4562 | 28.92 (20.29 to 39.41) | 96.15 | 0.075 |
| | | > 4 months | n = 5 | 155/961 | 15.41 (5.74 to 35.30) | 95.81 | |
| | Palpitations | > 4 months | n = 1 | 154/1655 | 9.31 (8.00 to 10.80) | NA | 0.863 |
| | | < 4 months | n = 7 | 322/3123 | 9.71 (5.42 to 16.78) | 94.42 | |
| | Excessive sputum/ Expectoration | > 4 months | n = 1 | 11/114 | 9.65 (5.42 to 16.59) | NA | 0.069 |
| | | < 4 months | n = 5 | 102/1835 | 4.95 (2.64 to 9.09) | 85.74 | |
| Gastrointestinal | Weight loss | > 4 months | n = 1 | 47/434 | 10.83 (8.23 to 14.12) | NA | <0.001 |
| | | < 4 months | n = 1 | 50/134 | 37.31 (29.55 to 45.79) | NA | |
| | Loss of appetite | > 4 months | n = 1 | 138/1655 | 8.34 (7.10 to 9.77) | NA | <0.001 |
| | | < 4 months | n = 2 | 64/251 | 25.50 (5.15 to 68.31) | 0 | |
| | Diarrhoea | < 4 months | n = 8 | 85/1836 | 3.53 (1.41 to 8.59) | 84.4 | 0.371 |
| | | > 4 months | n = 2 | 105/2089 | 5.03 (1.46 to 15.89) | 0 | |
| Musculoskeletal | Impaired mobility | > 4 months | n = 3 | 241/1953 | 24.29 (2.15 to 82.41) | 99.05 | 0.108 |
| | | < 4 months | n = 3 | 82/913 | 8.07 (0.95 to 44.49) | 96.49 | |
| | Joint pain/ Arthralgia | > 4 months | n = 1 | 154/1655 | 9.31 (8.00 to 10.80) | NA | 0.986 |
| | | < 4 months | n = 8 | 283/2305 | 9.35 (5.22 to 16.17) | 94.04 | |
| | Muscle pain/ Myalgia | < 4 months | n = 11 | 339/3127 | 12.95 (7.31 to 21.91) | 96.15 | <0.001 |
| | | > 4 months | n = 1 | 39/1655 | 2.36 (1.73 to 3.21) | NA | |
| Neurocognitive | Other cognitive impairment | < 4 months | n = 2 | 118/264 | 40.19 (0.00 to 100.00) | 99 | 0.017 |
| | | > 4 months | n = 1 | 4/177 | 2.26 (0.85 to 5.86) | NA | |
| Neurological and neuromuscular | Headache | > 4 months | n = 3 | 113/2269 | 6.11 (0.65 to 39.33) | 97.98 | 0.620 |
| | | < 4 months | n = 8 | 114/2266 | 4.42 (1.60 to 11.59) | 92.68 | |
| | Smell disturbance | < 4 months | n = 13 | 367/2994 | 13.26 (8.37 to 20.37) | 94.8 | 0.166 |
| | | > 4 months | n = 6 | 475/2674 | 19.96 (11.27 to 32.87) | 97.79 | |
| | Taste disturbance | > 4 months | n = 6 | 364/2674 | 15.36 (7.94 to 27.63) | 97.61 | 0.580 |

| | | | | | | | |
|---|---|---|---|---|---|---|---|
| | | < 4 months | n = 11 | 323/2749 | 12.61 (6.95 to 21.81) | 96.44 | |
| Other | Hair loss | < 4 months | n = 4 | 204/1155 | 12.72 (3.10 to 39.89) | 95.98 | 0.181 |
| | | > 4 months | n = 1 | 359/1655 | 21.69 (19.77 to 23.74) | NA | |
| | Skin rash | > 4 months | n = 1 | 47/1655 | 2.84 (2.14 to 3.76) | NA | 0.952 |
| | | < 4 months | n = 3 | 20/719 | 2.75 (0.30 to 20.89) | 86.08 | |
| Psychological and social | Care dependency | > 4 months | n = 2 | 39/1788 | 3.38 (0.00 to 98.56) | 95.89 | 0.006 |
| | | < 4 months | n = 1 | 121/767 | 15.78 (13.36 to 18.53) | NA | |
| | Reduced quality of life | < 4 months | n = 2 | 287/630 | 40.64 (2.65 to 94.52) | 88.36 | 0.119 |
| | | > 4 months | n = 1 | 53/177 | 29.94 (23.66 to 37.09) | NA | |
| | PTSD | < 4 months | n = 5 | 322/1937 | 9.93 (3.21 to 26.84) | 96.87 | 0.322 |
| | | > 4 months | n = 1 | 7/120 | 5.83 (2.81 to 11.73) | NA | |
| | Sleep disorder | > 4 months | n = 1 | 437/1655 | 26.40 (24.34 to 28.58) | NA | 0.130 |
| | | < 4 months | n = 8 | 305/1787 | 17.20 (8.21 to 32.54) | 94.08 | |
| | Depression | > 4 months | n = 1 | 367/1617 | 22.70 (20.72 to 24.80) | NA | <0.001 |
| | | < 4 months | n = 5 | 118/2045 | 6.16 (3.99 to 9.40) | 71.69 | |
| | Anxiety | > 4 months | n = 1 | 367/1617 | 22.70 (20.72 to 24.80) | NA | 0.492 |
| | | < 4 months | n = 6 | 283/1934 | 18.10 (7.16 to 38.77) | 97.51 | |
| Systemic | Dizziness | > 4 months | n = 2 | 130/2089 | 6.22 (2.06 to 17.34) | 0 | 0.011 |
| | | < 4 months | n = 3 | 32/1052 | 3.20 (1.08 to 9.12) | 68.78 | |
| | Fever | < 4 months | n = 5 | 23/1535 | 1.26 (0.22 to 7.01) | 90.73 | 0.762 |
| | | > 4 months | n = 2 | 24/2089 | 0.79 (0.00 to 100.00) | 96.18 | |
| | Fatigue | < 4 months | n = 10 | 890/3243 | 32.50 (23.93 to 42.42) | 94.57 | 0.608 |
| | | > 4 months | n = 7 | 1317/2796 | 28.61 (16.00 to 45.73) | 98.47 | |
| Upper respiratory | Nasal congestion | > 4 months | n = 1 | 23/434 | 5.30 (3.55 to 7.85) | NA | 0.690 |
| | | < 4 months | n = 2 | 27/569 | 4.75 (0.41 to 37.90) | 25.27 | |
| | Sore throat | > 4 months | n = 1 | 69/1655 | 4.17 (3.31 to 5.25) | NA | 0.655 |
| | | < 4 months | n = 4 | 58/1241 | 4.84 (1.76 to 12.65) | 85.28 | |

#### Supplement 9: Meta-regression: % Female

| Classification | Symptom | N Studies | Constant (SE) | Beta (SE) | $R^2$ | P value |
|---|---|---|---|---|---|---|
| Cardiopulmonary | Breathlessness/ Exertional dyspnoea | 20 | -0.23 (0.79) | -1.93 (1.71) | 0.07 | 0.258 |
| | Chest pain | 11 | -2.13 (1.85) | -1.27 (4.11) | 0.01 | 0.758 |
| | Cough | 16 | -2.43 (1.12) | 0.02 (2.32) | 0.00 | 0.994 |
| Gastrointestinal | Diarrhoea | 10 | -3.9 (1.46) | 1.46 (2.87) | 0.00 | 0.612 |
| Systemic | Fatigue | 17 | -0.08 (0.61) | -1.58 (1.29) | 0.09 | 0.222 |
| Musculoskeletal | Muscle pain/ Myalgia | 12 | -0.37 (1.33) | -3.97 (3.05) | 0.13 | 0.194 |
| Neurological and neuromuscular | Headache | 11 | -6.29 (1.27) | 6.7 (2.46) | 0.43 | 0.007 |
| | Smell disturbance | 19 | -4.07 (0.53) | 4.95 (1.08) | 0.56 | <0.001 |
| | Taste disturbance | 17 | -4.29 (0.63) | 5.04 (1.27) | 0.51 | <0.001 |

*Figure 13. Metaregression on percentage of female. Neurological and neuromuscular (Headache)*

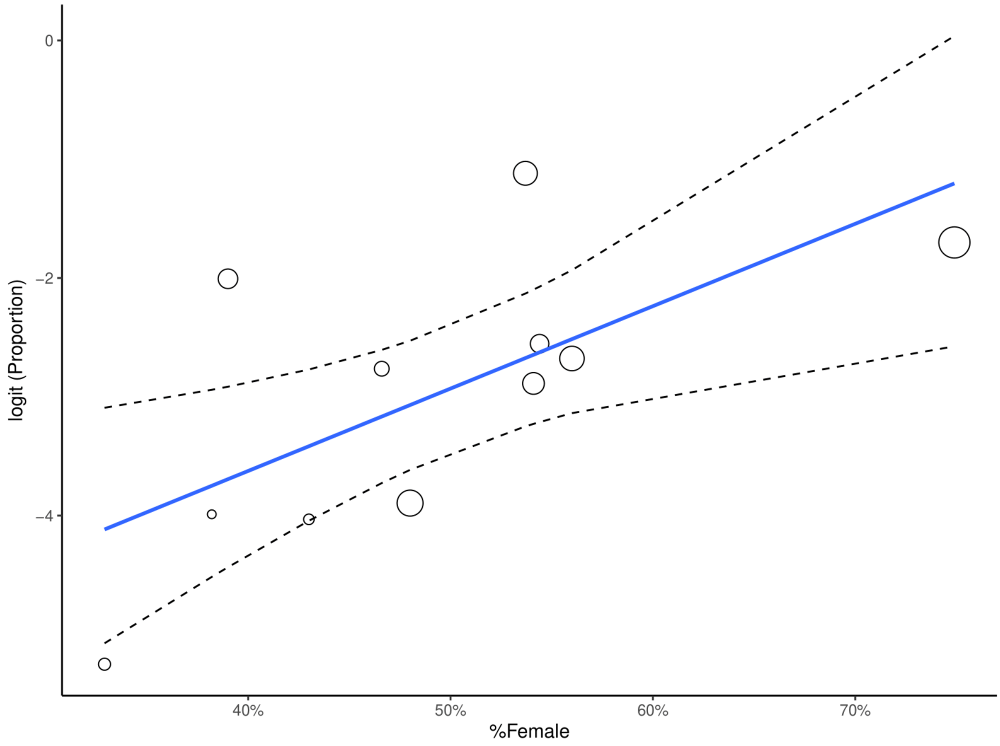

*Figure 14. Metaregression on percentage of female. Neurological and neuromuscular (Small disturbance).*

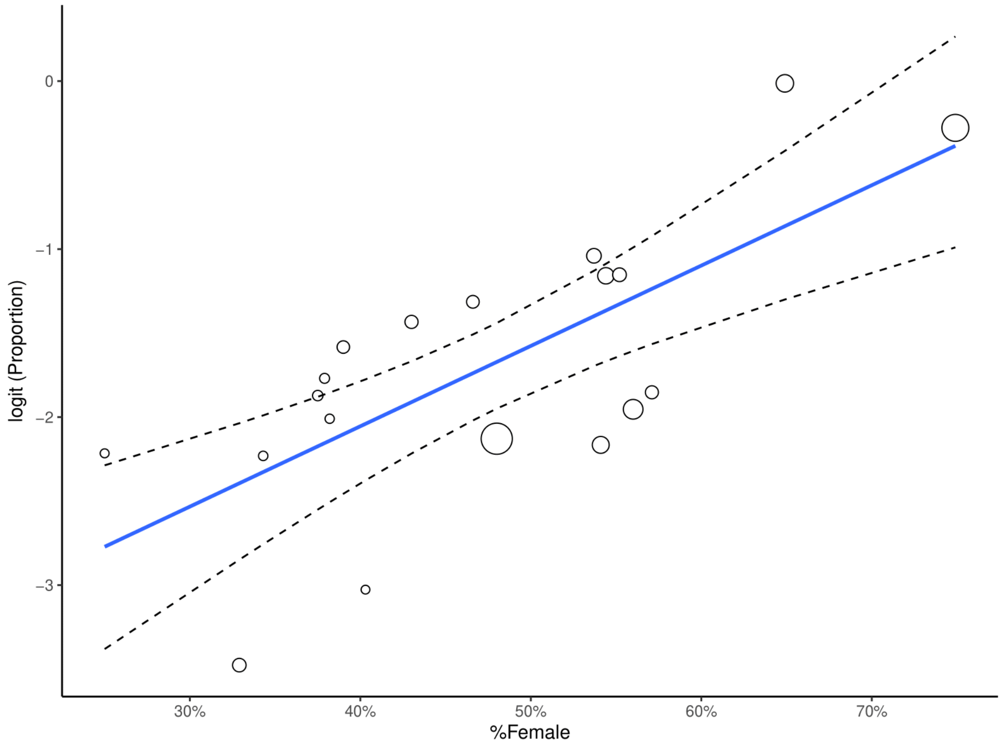

*Figure 15. Metaregression on percentage of female. Neurological and neuromuscular (Taste disturbance).*

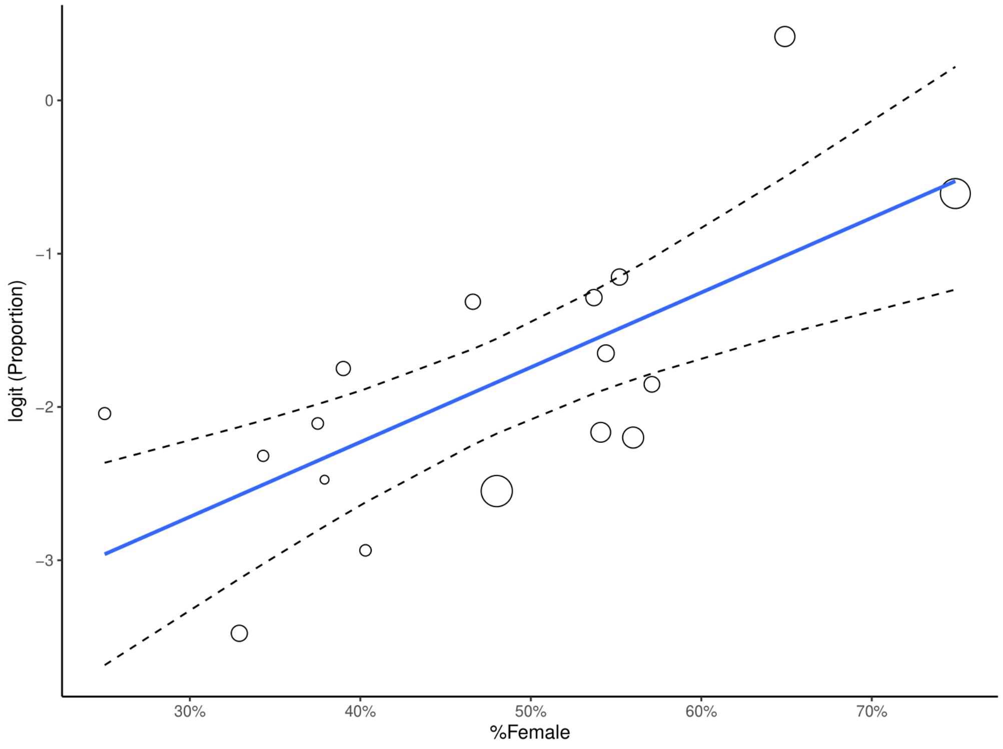

#### Supplement 10: Meta-regression: % ICU patients

| Classification | Symptom | N Studies | Constant (SE) | Beta (SE) | $R^2$ | P value |
|---|---|---|---|---|---|---|
| Cardiopulmonary | Breathlessness/ Exertional dyspnoea | 14 | -1.03 (0.31) | 1.02 (0.89) | 0.09 | 0.254 |
| | Cough | 11 | -2.25 (0.63) | -0.75 (2.73) | -0.01* | 0.783 |
| Systemic | Fatigue | 11 | -0.67 (0.29) | 0.4 (0.81) | 0.02 | 0.620 |
| Musculoskeletal | Muscle pain/ Myalgia | 11 | -2.71 (0.46) | 4.19 (2.12) | 0.27 | 0.048 |
| Neurological and neuromuscular | Smell disturbance | 14 | -2.24 (0.26) | 1.32 (1.3) | 0.08 | 0.311 |
| | Taste disturbance | 12 | -2.45 (0.25) | 1.72 (1.23) | 0.16 | 0.161 |

*poor fitting

*Figure 16. Metaregression on percentage of ICU patients. Musculoskeletal (Muscle pain/ Myalgia).*

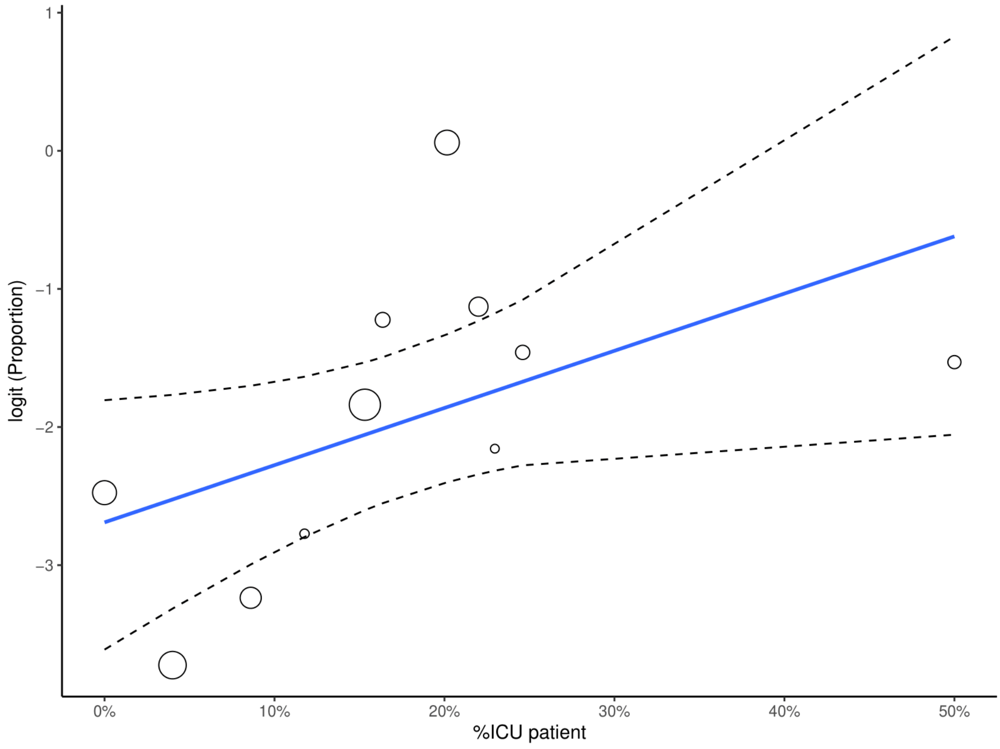

#### Supplement 11: Sensitivity analysis: versus removing high risk of bias studies

| Classification | Symptom | Main results | | | Main results after removing high risk of bias studies | | |
|---|---|---|---|---|---|---|---|
| | | N Studies | n/Total | Prop (95% CIs) | N Studies | n/Total | Prop (95% CIs) |
| Cardiopulmonary | Other cardiovascular symptoms | n = 3 | 79/1952 | 1.38 (0.01 to 67.44) | n = 2 | 71/1305 | 1.37 (0.04 to 32.38) |
| | Palpitations | n = 8 | 476/4778 | 9.67 (5.95 to 15.34) | n = 7 | 413/4131 | 9.65 (5.39 to 16.66) |
| | Excessive sputum/ Expectoration | n = 6 | 113/1949 | 5.46 (3.19 to 9.19) | n = 4 | 88/1326 | 6.22 (2.62 to 14.05) |
| | Cough | n = 16 | 414/5031 | 8.17 (4.85 to 13.44) | n = 11 | 265/3207 | 7.42 (3.38 to 15.55) |
| | Chest pain | n = 11 | 308/4878 | 6.36 (3.15 to 12.42) | n = 8 | 275/3939 | 7.33 (3.25 to 15.68) |
| | Breathlessness/ Exertional dyspnoea | n = 20 | 1297/5523 | 25.06 (17.86 to 33.97) | n = 14 | 992/3596 | 28.85 (19.67 to 40.16) |
| Gastrointestinal | Weight loss | n = 2 | 97/568 | 20.99 (8.09 to 44.51) | n = 1 | 50/134 | 37.31 (29.12 to 46.08) |
| | Stomach/ Abdominal pain | n = 4 | 30/1427 | 2.33 (0.54 to 9.42) | n = 2 | 7/859 | 0.81 (0.39 to 1.70) |
| | Loss of appetite | n = 3 | 202/1906 | 17.49 (4.13 to 51.04) | n = 2 | 174/1789 | 15.09 (6.35 to 31.80) |
| | Diarrhoea | n = 10 | 190/3925 | 4.00 (2.07 to 7.57) | n = 6 | 140/2751 | 4.68 (2.49 to 8.66) |
| | Nausea or Vomiting | n = 4 | 49/821 | 6.69 (1.64 to 23.59) | n = 2 | 20/253 | 7.91 (5.16 to 11.93) |
| Musculoskeletal | Impaired mobility | n = 6 | 323/2866 | 14.42 (4.67 to 36.73) | n = 5 | 257/2662 | 12.00 (3.02 to 37.39) |
| | Joint pain/ Arthralgia | n = 9 | 437/3960 | 9.39 (5.72 to 15.03) | n = 6 | 378/3215 | 10.79 (5.21 to 21.00) |
| | Muscle pain/ Myalgia | n = 12 | 378/4782 | 11.29 (6.17 to 19.75) | n = 10 | 320/4209 | 11.14 (5.35 to 21.75) |
| Neurocognitive | Other cognitive impairment | n = 3 | 122/441 | 17.77 (0.08 to 98.23) | n = 3 | 122/441 | 17.77 (0.08 to 98.23) |
| | Confusion | n = 2 | 33/1218 | 2.71 (1.93 to 3.79) | n = 1 | 23/767 | 3.00 (1.91 to 4.47) |
| | Concentration impairment | n = 2 | 66/254 | 25.98 (20.96 to 31.73) | n = 1 | 34/134 | 25.37 (18.26 to 33.61) |
| | Memory impairment | n = 5 | 151/886 | 17.94 (5.26 to 46.25) | n = 4 | 110/766 | 14.93 (2.77 to 51.97) |
| Neurological and neuromuscular | Tingling/ Paraesthesia | n = 2 | 33/257 | 9.12 (2.21 to 30.87) | n = 1 | 29/135 | 21.48 (14.88 to 29.37) |
| | Visual disturbance | n = 2 | 28/586 | 4.78 (3.32 to 6.83) | n = 1 | 9/135 | 6.67 (3.09 to 12.28) |
| | Smell disturbance | n = 19 | 842/5668 | 15.17 (10.75 to 20.97) | n = 13 | 513/4258 | 14.08 (8.87 to 21.62) |
| | Taste disturbance | n = 17 | 687/5423 | 13.52 (8.96 to 19.89) | n = 11 | 425/4013 | 13.44 (7.31 to 23.42) |
| | Tremors | n = 3 | 42/1124 | 3.53 (0.30 to 30.63) | n = 2 | 38/673 | 6.20 (3.68 to 10.26) |
| | Headache | n = 11 | 227/4535 | 4.88 (2.30 to 10.06) | n = 7 | 115/3298 | 4.19 (1.30 to 12.71) |
| Other | Hair loss | n = 5 | 563/2810 | 14.34 (5.33 to 33.23) | n = 4 | 539/2690 | 13.14 (3.17 to 41.12) |

| | | | | | | | |
|---|---|---|---|---|---|---|---|
| | Skin rash | n = 4 | 67/2374 | 2.83 (0.95 to 8.16) | n = 3 | 60/1923 | 3.53 (0.75 to 15.11) |
| Psychological and social | Sleep disorder | n = 9 | 742/3442 | 18.15 (9.61 to 31.63) | n = 8 | 705/3322 | 16.88 (8.18 to 31.65) |
| Systemic | Dizziness | n = 5 | 162/3141 | 4.50 (2.53 to 7.86) | n = 4 | 133/2707 | 4.02 (1.87 to 8.42) |
| | Fever | n = 7 | 47/3624 | 1.08 (0.24 to 4.66) | n = 5 | 23/2739 | 0.91 (0.11 to 7.18) |
| | Fatigue | n = 17 | 2207/6039 | 30.97 (23.91 to 39.03) | n = 12 | 1882/4555 | 33.24 (24.57 to 43.22) |
| Upper respiratory | Other respiratory symptoms | n = 3 | 253/1111 | 15.58 (0.68 to 83.17) | n = 1 | 210/538 | 39.03 (34.89 to 43.30) |
| | Nasal congestion | n = 3 | 50/1003 | 4.99 (2.73 to 8.92) | n = 1 | 8/118 | 6.78 (2.97 to 12.92) |
| | Sore throat | n = 5 | 127/2896 | 4.70 (2.42 to 8.91) | n = 4 | 106/2445 | 4.70 (1.73 to 12.10) |

#### Supplement 12: Sensitivity analysis: versus statistical methods

| Classification | Symptom | N Studies | n/Total | Main results | FTDAT/IV* |
|---|---|---|---|---|---|
| | | | | Prop (95% CIs) | Prop (95% CIs) |
| Cardiopulmonary | Breathlessness/ Exertional dyspnoea | n = 20 | 1297/5523 | 25.06 (17.86 to 33.97) | 26.68 (20.36 to 33.51) |
| | Palpitations | n = 8 | 476/4778 | 9.67 (5.95 to 15.34) | 10.21 (6.76 to 14.26) |
| | Cough | n = 16 | 414/5031 | 8.17 (4.85 to 13.44) | 9.52 (6.16 to 13.50) |
| | Chest pain | n = 11 | 308/4878 | 6.36 (3.15 to 12.42) | 7.52 (4.29 to 11.52) |
| | Excessive sputum/ Expectoration | n = 6 | 113/1949 | 5.46 (3.19 to 9.19) | 5.69 (3.23 to 8.75) |
| | Flushing | n = 1 | 26/538 | 4.83 (3.18 to 7.00) | 4.83 (3.17 to 6.82) |
| | Newly diagnosed hypertension | n = 1 | 7/538 | 1.30 (0.52 to 2.66) | 1.30 (0.49 to 2.46) |
| | Other cardiovascular symptoms | n = 3 | 79/1952 | 1.38 (0.01 to 67.44) | 2.99 (0.00 to 12.59) |
| Gastrointestinal | Weight loss | n = 2 | 97/568 | 20.99 (8.09 to 44.51) | 22.47 (3.00 to 52.50) |
| | Other stomach/ Abdominal discomfort | n = 1 | 21/117 | 17.95 (11.47 to 26.12) | 17.95 (11.47 to 25.47) |
| | Loss of appetite | n = 3 | 202/1906 | 17.49 (4.13 to 51.04) | 18.57 (6.35 to 35.21) |
| | Nausea or Vomiting | n = 4 | 49/821 | 6.69 (1.64 to 23.59) | 7.69 (1.89 to 16.67) |
| | Diarrhoea | n = 10 | 190/3925 | 4.00 (2.07 to 7.57) | 4.41 (2.65 to 6.57) |
| | Bloody stools / Haematochezia | n = 1 | 2/117 | 1.71 (0.21 to 6.04) | 1.71 (0.03 to 5.08) |
| | Stomach/ Abdominal pain | n = 4 | 30/1427 | 2.33 (0.54 to 9.42) | 2.63 (0.56 to 5.93) |
| Musculoskeletal | Impaired mobility | n = 6 | 323/2866 | 14.42 (4.67 to 36.73) | 17.09 (7.35 to 29.77) |
| | Muscle pain/ Myalgia | n = 12 | 378/4782 | 11.29 (6.17 to 19.75) | 13.09 (7.71 to 19.59) |
| | Joint pain/ Arthralgia | n = 9 | 437/3960 | 9.39 (5.72 to 15.03) | 10.04 (6.33 to 14.46) |
| Neurocognitive | Concentration impairment | n = 2 | 66/254 | 25.98 (20.96 to 31.73) | 25.98 (20.74 to 31.59) |
| | Memory impairment | n = 5 | 151/886 | 17.94 (5.26 to 46.25) | 20.55 (6.54 to 39.62) |
| | Other cognitive impairment | n = 3 | 122/441 | 17.77 (0.08 to 98.23) | 25.51 (0.00 to 79.72) |
| | Frontal release signs | n = 1 | 20/135 | 14.81 (9.29 to 21.95) | 14.81 (9.27 to 21.35) |
| | Confusion | n = 2 | 33/1218 | 2.71 (1.93 to 3.79) | 2.69 (1.84 to 3.69) |
| Neurological and neuromuscular | Abnormal reflex status | n = 1 | 31/135 | 22.96 (16.17 to 30.98) | 22.96 (16.22 to 30.47) |
| | Other neurological diseases | n = 1 | 20/135 | 14.81 (9.29 to 21.95) | 14.81 (9.27 to 21.35) |
| | Smell disturbance | n = 19 | 842/5668 | 15.17 (10.75 to 20.97) | 16.48 (11.36 to 22.31) |
| | Taste disturbance | n = 17 | 687/5423 | 13.52 (8.96 to 19.89) | 14.99 (9.76 to 21.09) |
| | Decreased sensation or sensibility | n = 2 | 30/269 | 10.90 (6.71 to 17.22) | 10.88 (4.73 to 19.05) |
| | Tingling/ Paraesthesia | n = 2 | 33/257 | 9.12 (2.21 to 30.87) | 10.74 (0.02 to 34.12) |
| | Muscle atrophy | n = 1 | 9/135 | 6.67 (3.09 to 12.28) | 6.67 (2.98 to 11.58) |
| | Headache | n = 11 | 227/4535 | 4.88 (2.30 to 10.06) | 6.12 (2.97 to 10.25) |
| | Slowness of movement/ Bradykinesia | n = 1 | 7/135 | 5.19 (2.11 to 10.39) | 5.19 (1.98 to 9.67) |
| | Visual disturbance | n = 2 | 28/586 | 4.78 (3.32 to 6.83) | 4.86 (2.79 to 7.43) |
| | Abnormal muscle tone | n = 1 | 6/135 | 4.44 (1.65 to 9.42) | 4.44 (1.50 to 8.68) |
| | Tremors | n = 3 | 42/1124 | 3.53 (0.30 to 30.63) | 4.12 (0.76 to 9.75) |
| | Walking/ Gait abnormality | n = 3 | 34/809 | 4.20 (2.02 to 8.53) | 4.11 (2.80 to 5.63) |

| | | | | | |
|---|---|---|---|---|---|
| | Trigeminal neuralgia | n = 1 | 4/122 | 3.28 (0.90 to 8.18) | 3.28 (0.71 to 7.33) |
| | Speech difficulty/ Dysarthria | n = 1 | 3/135 | 2.22 (0.46 to 6.36) | 222 (0.28 to 5.56) |
| | Ear/ Hearing conditions | n = 1 | 5/451 | 1.11 (0.36 to 2.57) | 1.11 (0.31 to 2.33) |
| | Lack of coordination/ Dysmetria | n = 1 | 2/135 | 1.48 (0.18 to 5.25) | 1.48 (0.02 to 4.41) |
| | Seizures/ Cramps | n = 1 | 6/451 | 1.33 (0.49 to 2.87) | 1.33 (0.44 to 2.63) |
| Other | Hair loss | n = 5 | 563/2810 | 14.34 (5.33 to 33.23) | 15.86 (7.42 to 26.68) |
| | Skin rash | n = 4 | 67/2374 | 2.83 (0.95 to 8.16) | 2.86 (1.29 to 4.96) |
| | Conjunctivitis | n = 1 | 8/451 | 1.77 (0.77 to 3.47) | 1.77 (0.73 to 3.23) |
| Psychological and social | Reduced quality of life | n = 3 | 340/807 | 36.76 (18.43 to 59.93) | 36.60 (23.89 to 50.32) |
| | Anxiety | n = 7 | 650/3551 | 18.73 (8.89 to 35.25) | 20.39 (11.95 to 30.38) |
| | Sleep disorder | n = 9 | 742/3442 | 18.15 (9.61 to 31.63) | 20.01 (12.32 to 28.99) |
| | PTSD | n = 6 | 329/2057 | 9.14 (3.66 to 21.04) | 10.41 (3.36 to 20.59) |
| | Depression | n = 6 | 485/3662 | 8.06 (4.14 to 15.10) | 8.72 (3.02 to 16.93) |
| | Care dependency | n = 3 | 160/2555 | 5.89 (0.46 to 45.96) | 7.24 (0.36 to 21.24) |
| | Low mood/ Dysphoria | n = 3 | 62/898 | 1.79 (0.00 to 98.74) | 7.49 (0.00 to 31.80) |
| Systemic | Weakness | n = 2 | 186/513 | 41.20 (25.43 to 59.01) | 41.62 (19.16 to 66.08) |
| | General malaise | n = 2 | 292/672 | 32.68 (14.91 to 57.36) | 33.47 (8.11 to 65.59) |
| | Fatigue | n = 17 | 2207/6039 | 30.97 (23.91 to 39.03) | 31.75 (22.81 to 41.41) |
| | Sweat or night sweats | n = 2 | 162/683 | 23.72 (20.68 to 27.05) | 23.68 (20.55 to 26.96) |
| | Dizziness | n = 5 | 162/3141 | 4.50 (2.53 to 7.86) | 4.58 (2.80 to 6.76) |
| | Enlarged lymph nodes/ Lymphadenopathy | n = 1 | 4/451 | 0.89 (0.24 to 2.26) | 0.89 (0.19 to 2.01) |
| | Fever | n = 7 | 47/3624 | 1.08 (0.24 to 4.66) | 1.74 (0.28 to 4.14) |
| Upper respiratory | Other respiratory symptoms | n = 3 | 253/1111 | 15.58 (0.68 to 83.17) | 19.30 (0.95 to 52.06) |
| | Voice change | n = 1 | 11/134 | 8.21 (4.17 to 14.21) | 8.21 (4.08 to 13.53) |
| | Nasal congestion | n = 3 | 50/1003 | 4.99 (2.73 to 8.92) | 4.89 (3.61 to 6.34) |
| | Sore throat | n = 5 | 127/2896 | 4.70 (2.42 to 8.91) | 4.66 (2.94 to 6.74) |

* FTDAT/IV: Freeman-Tukey Double arcsine transformation within inverse Variance Method

#### Supplement 13: Funnel plots

*Figure 17. Funnel plot. Cardiopulmonary (Breathlessness or Exertional dyspnoea) by Egger's method*

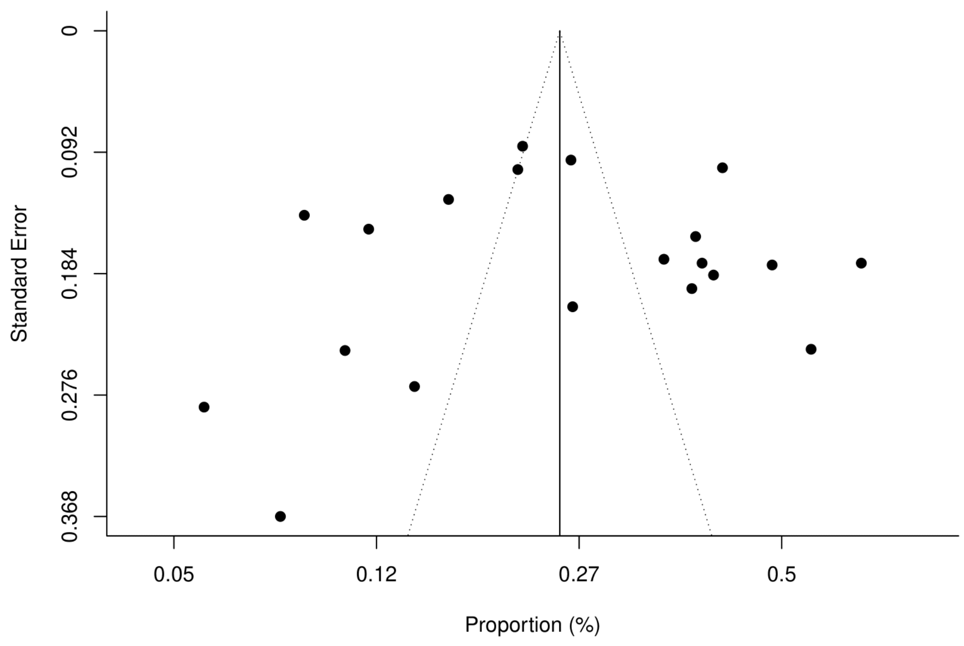

*Figure 18. Funnel plot. Cardiopulmonary (Breathlessness or Exertional dyspnoea) by Peter's method*

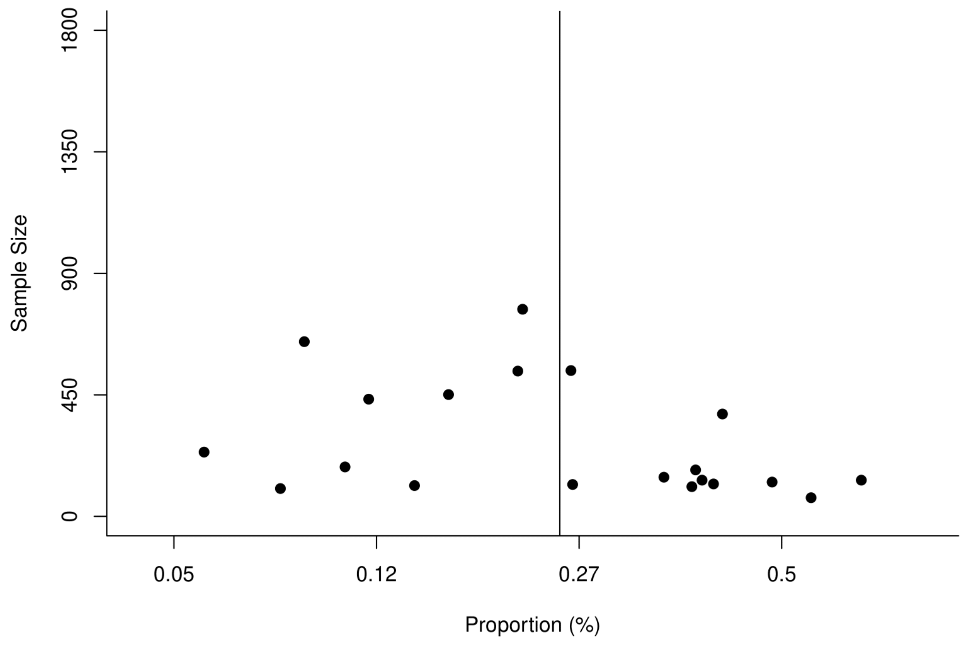

*Figure 19. Funnel plot. Cardiopulmonary (Chest pain) by Egger's method*

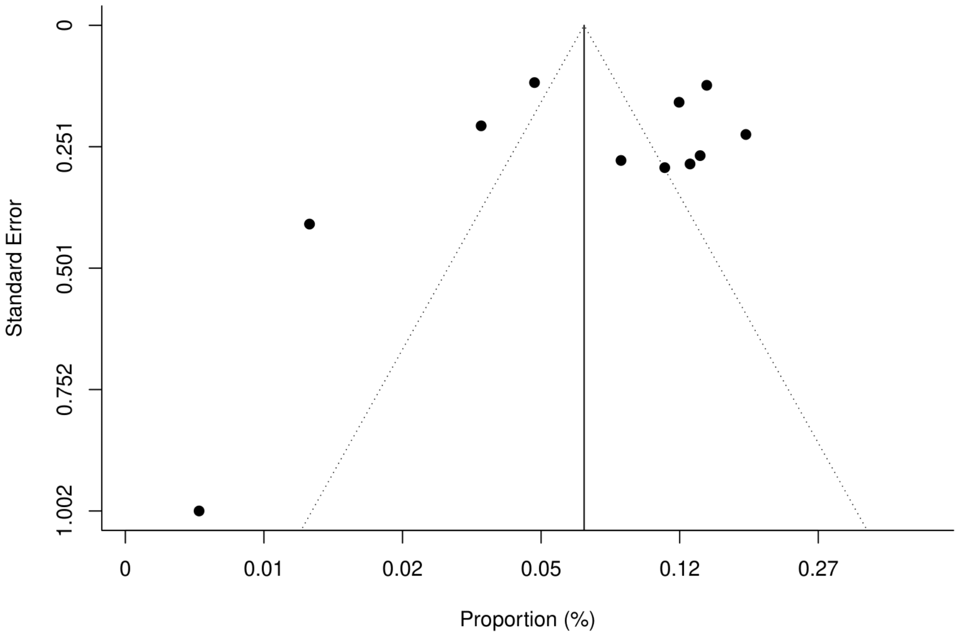

*Figure 20. Funnel plot. Cardiopulmonary (Chest pain) by Peter's method*

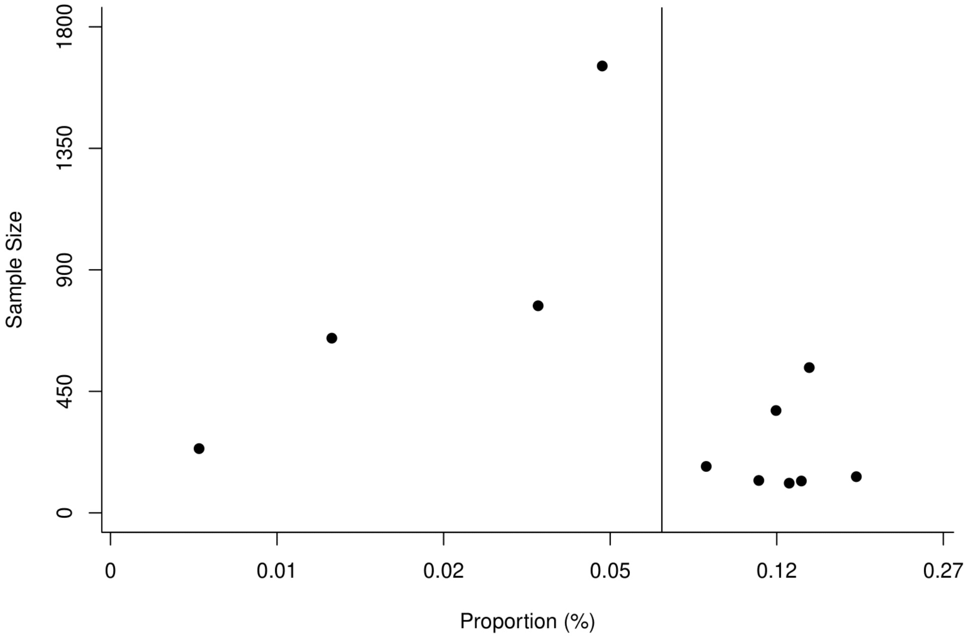

*Figure 21. Funnel plot. Cardiopulmonary (Cough) by Egger's method*

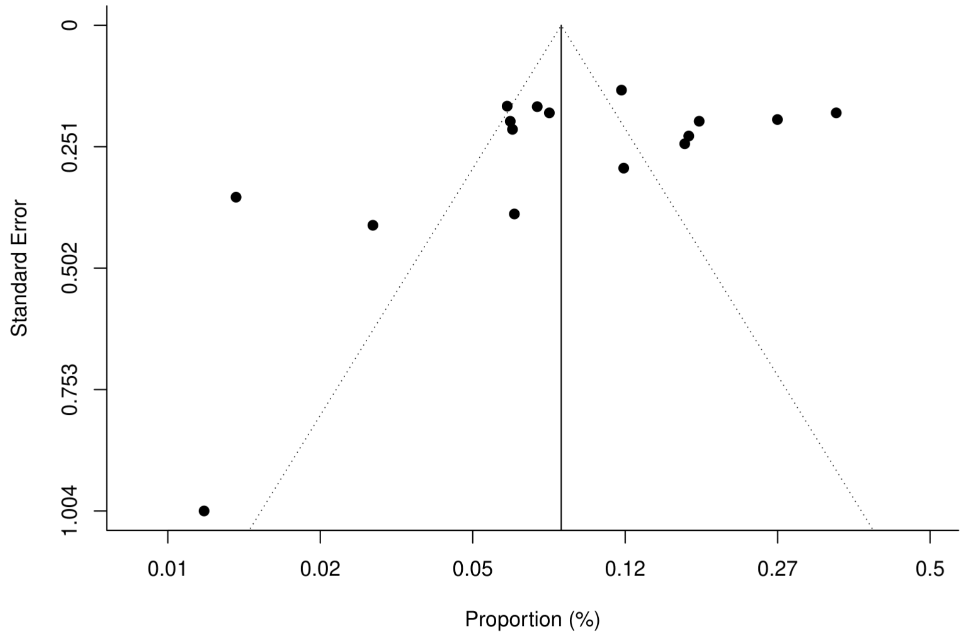

*Figure 22. Funnel plot. Cardiopulmonary (Cough) by Peter's method*

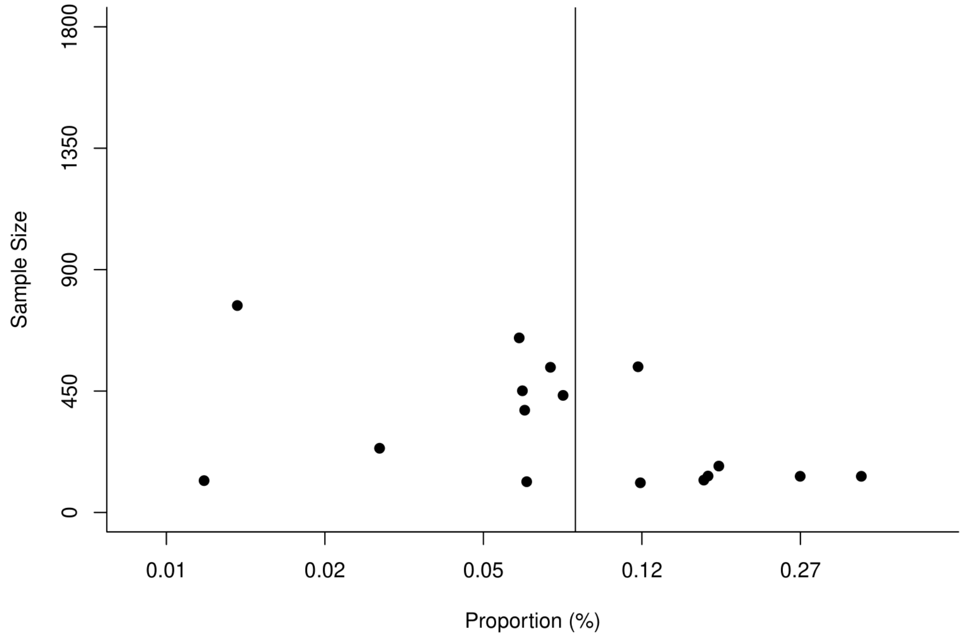

*Figure 23. Funnel plot. Gastrointestinal (Diarrhoea) by Egger's method*

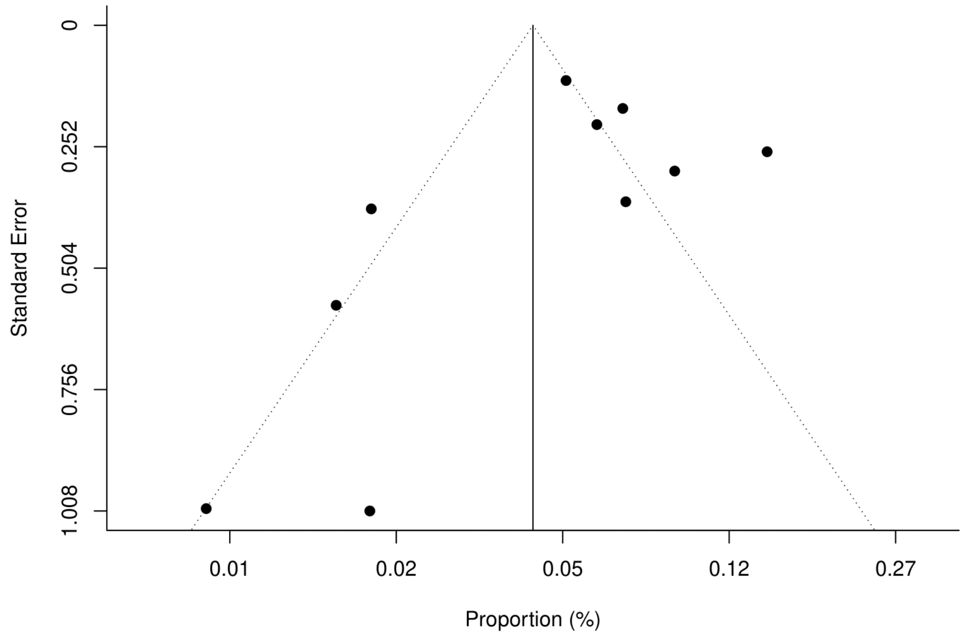

*Figure 24. Funnel plot. Gastrointestinal (Diarrhoea) by Peter's method*

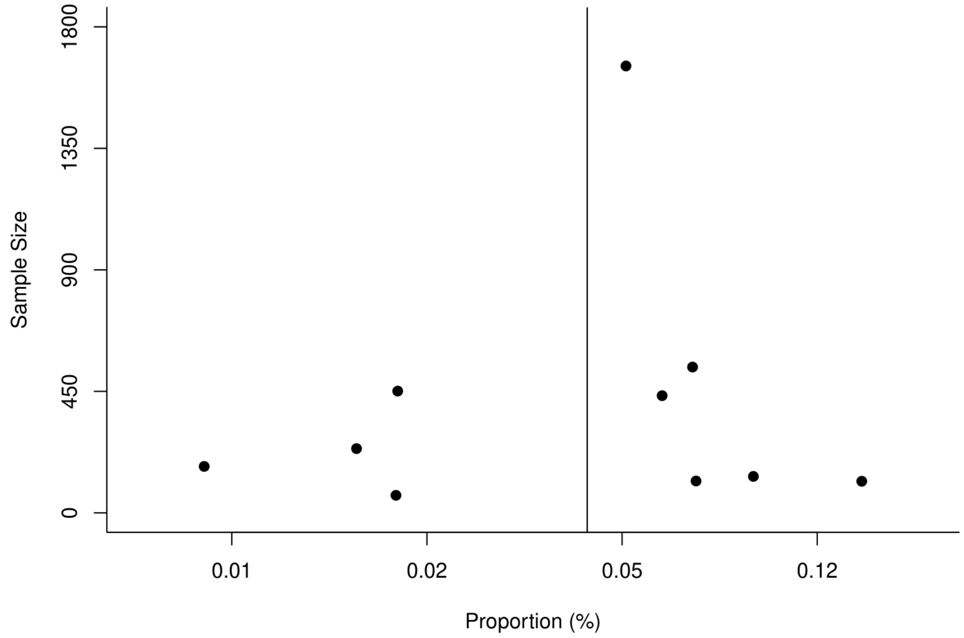

*Figure 25. Funnel plot. Musculoskeletal (Muscle pain or Myalgia) by Egger's method*

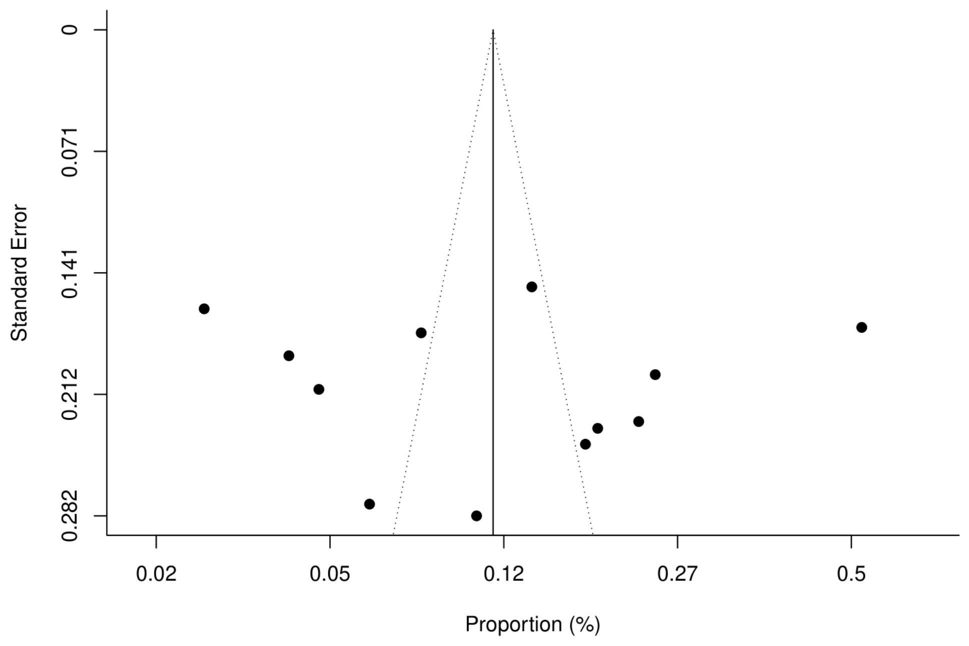

*Figure 26. Funnel plot. Musculoskeletal (Muscle pain or Myalgia) by Peter's method*

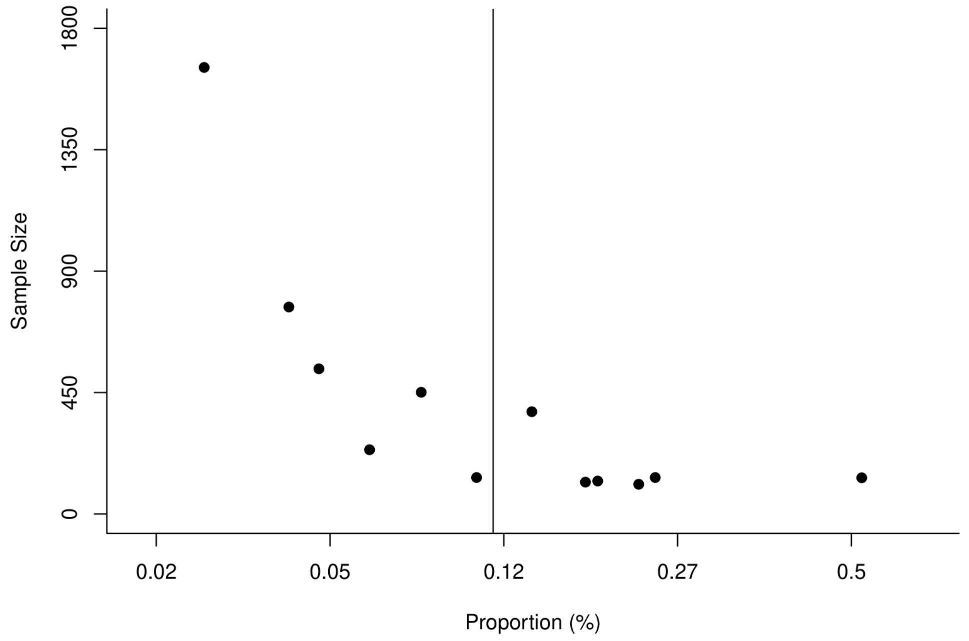

*Figure 27. Funnel plot. Neurological and neuromuscular (Headache) by Egger's method*

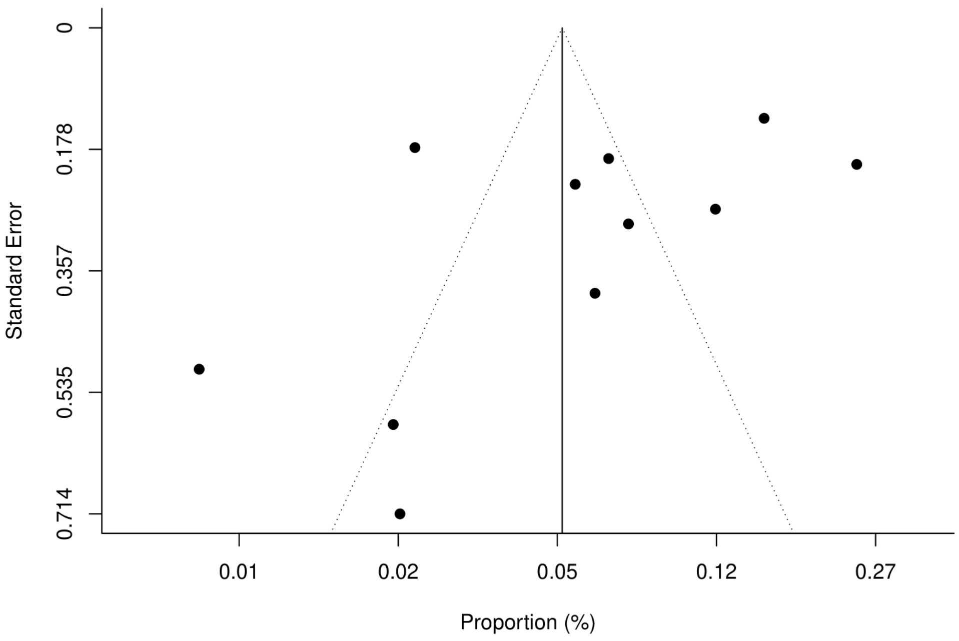

*Figure 28. Neurological and neuromuscular (Headache) by Peter's method*

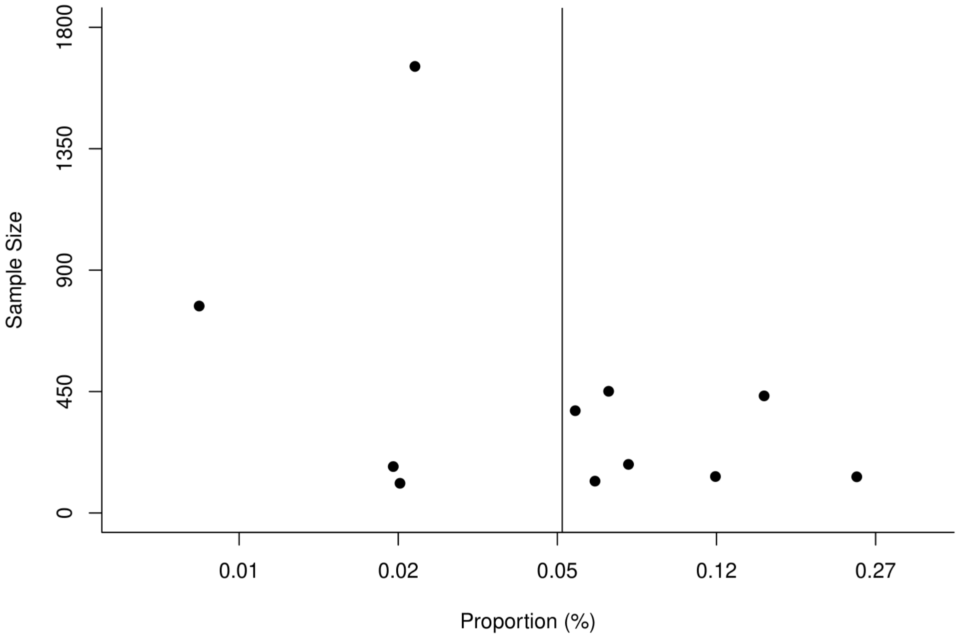

*Figure 29. Funnel plot. Neurological and neuromuscular (Smell disturbance) by Egger's method*

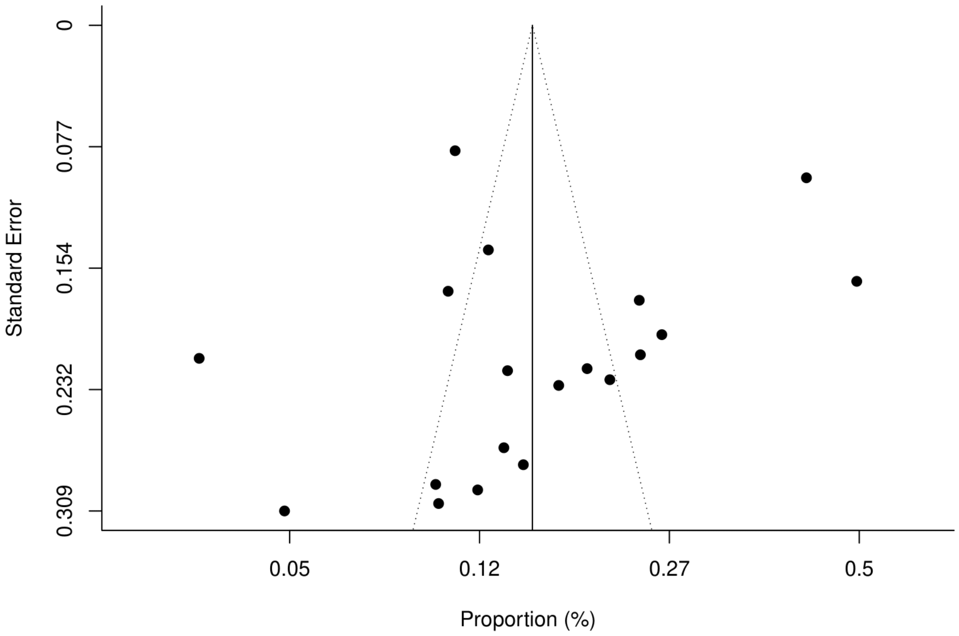

*Figure 30. Neurological and neuromuscular (Smell disturbance) by Peter's method*

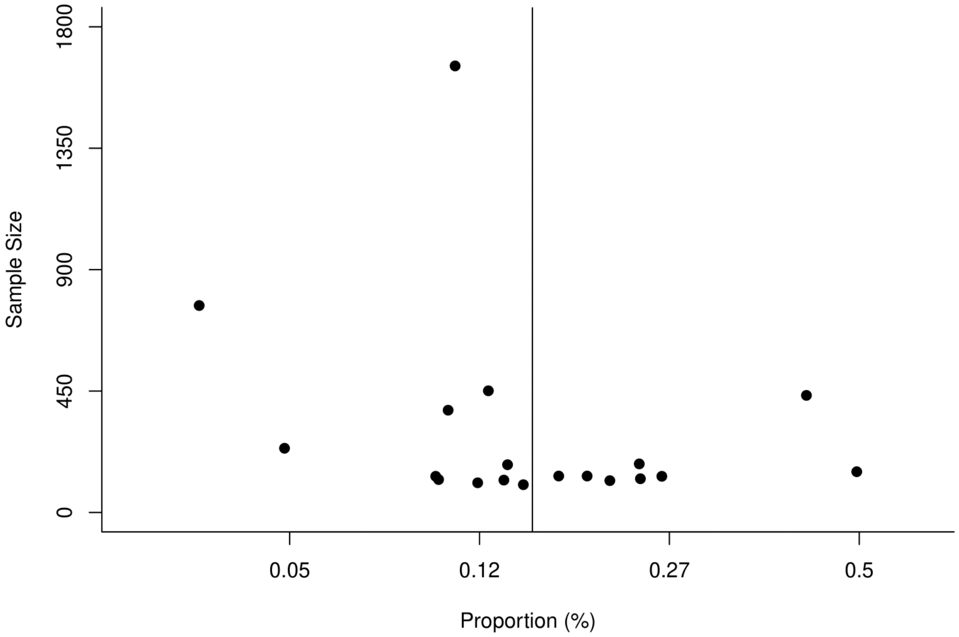

*Figure 31. Funnel plot. Neurological and neuromuscular (Taste disturbance) by Egger's method*

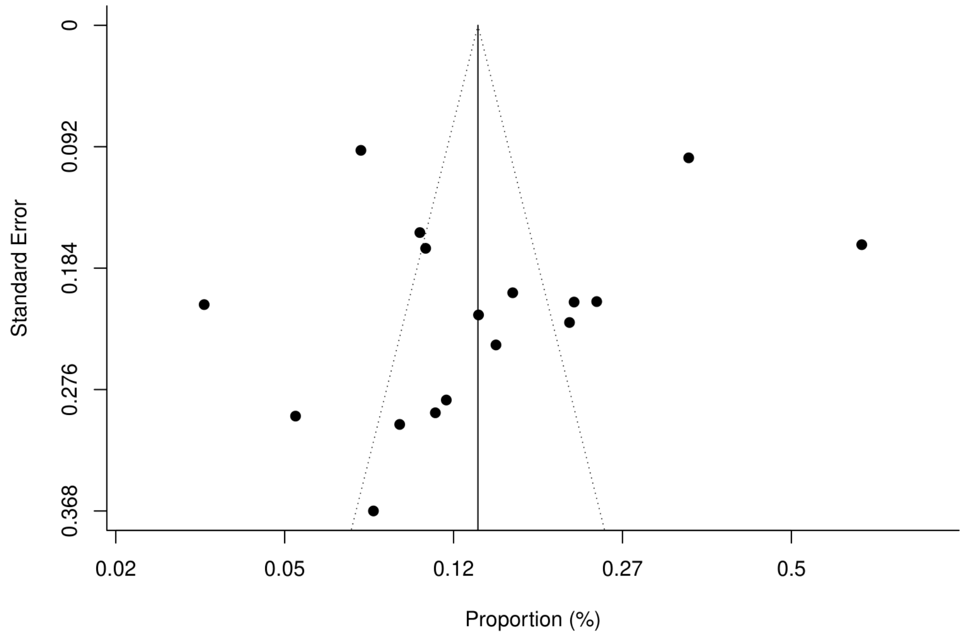

*Figure 32. Funnel plot. Neurological and neuromuscular (Taste disturbance) by Peter's method*

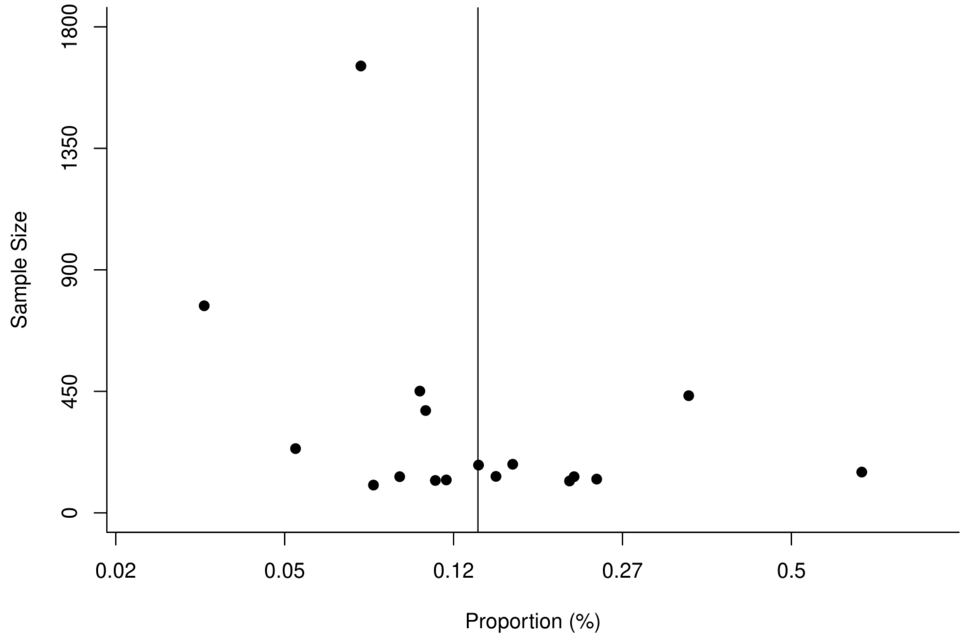

*Figure 33. Funnel plot. Systemic (Fatigue) by Egger's method*

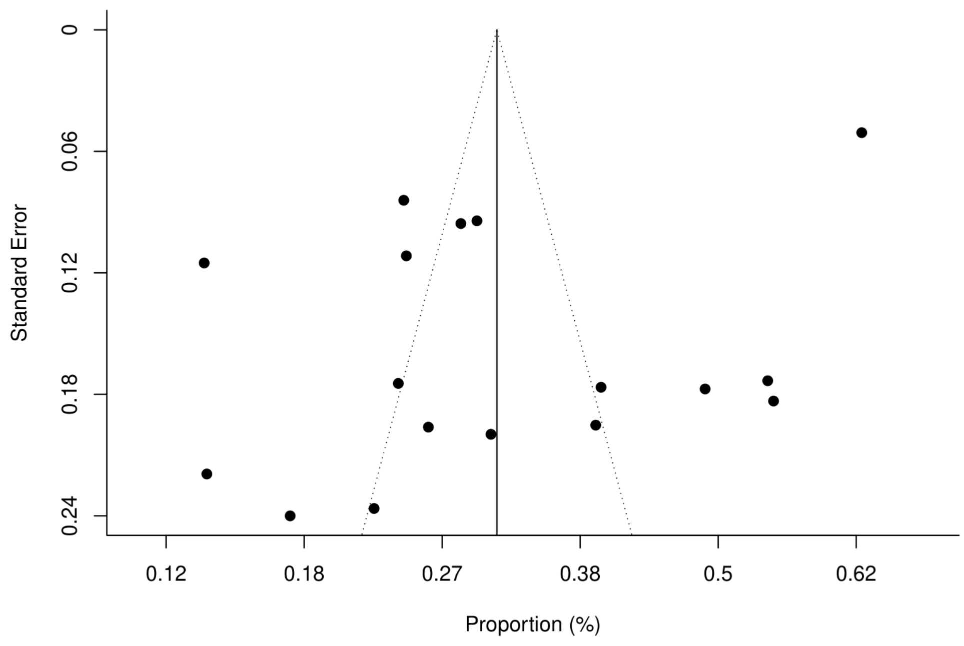

*Figure 34. Funnel plot.Systemic (Fatigue) by Peter's method*

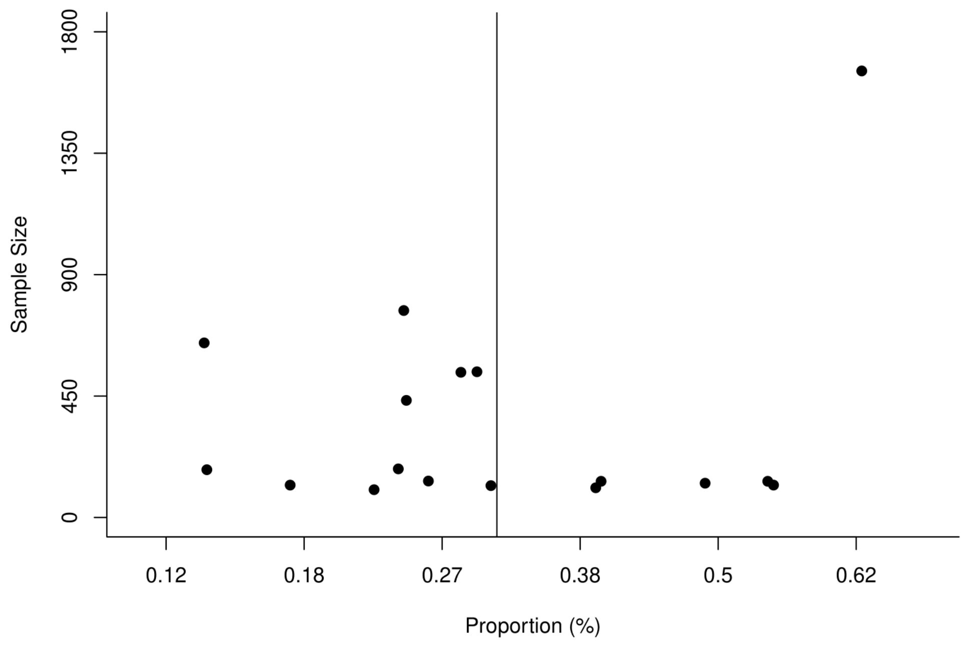

#### Supplement 14: Risk factors

| Study | Category | Risk factor | Associated with | Method | P Value/ CI |
|---|---|---|---|---|---|
| Nguyen et al. | Sex | Female sex | Persistent symptoms | Chi-squared or the Fisher exact test | p = 0.02 |
| Mazza et al. | Sex | Female sex | Persistence of depressive symptomatology | Multivariate GLM analysis | (Wilks' λ = 0.92; F = 5.76; p = 0.003) |
| | Comorbidities | Previous psychiatric diagnosis | | | (Wilks' λ = 0.93; F = 5.29; p = 0.006) |
| | Severity | Presence of psychopathology at one-month | | | (Wilks' λ = 0.82; F = 15.16; p < 0.001) |
| Parentes-Arias et al. | Age | <60 years | Olfactory dysfunction | Multivariable-adjusted ORs | p = 0.028 |
| | Sex | Female sex | | | p = 0.003 |
| | Comorbidities | 1 comorbidity | | | p = 0.031 |
| Xiong et al. | Sex | Female sex | Covid-19 sequelae | Multivariable logistic regression model | Physical decline/fatigue (p < 0.01)<br>Postactivity polypnoea (p= 0.04)<br>Alopecia (p < 0.01) |
| | Severity | Dyspnea during hospitalisation | Physical decline/fatigue, postactivity polypnoea and resting heart rate increases | Univariate analysis | Physical decline/fatigue (p=.02)<br>Postactivity polypnoea (p=.01)<br>Resting heart rate increases (p=.01) |
| Sykes et al. | Sex | Female sex | Persistent symptoms | Chi-Square and Mann–Whitney U testing | Anxiety (p=0.001),low mood (p=0.031), myalgia (p=0.022), fatigue (p=0.004), sleep disturbance (p=0.009), and memory impairment (p=0.001) |
| Taboada et al. | Age | Age | Limitations in the functional status (grade II-IV of PCSF) | Multivariate logistic regression model | (OR = 2.600, 95% CI: 1.192–5.671) |
| | Severity | Length of hospital stay | | | (OR = 1.049, 95% CI: 1.009–1.090) |
| | Severity | Admission to ICU / mechanical ventilation | | | P < 0.001 |
| Qu et al. | Sex | Female sex | Poor QoL scores | Logistic regression | (OR: 1.79, 95% CI: 1.04–3.06) |
| | Age | Older age (≥60 years) | | | (OR: 2.44, 95% CI: 1.33–4.47) |
| | Severity | Physical symptom after discharge | | | (OR: 40.15, 95% CI: 9.68–166.49) |
| Einvik et al. | Sex | Female sex | Symptoms of post-traumatic stress | Multivariable linear regression | NR |
| | Ethnicity | Born outside Norway | | | |
| | Severity | Dyspnoea during COVID-19 | | | |

| | | | | | |
|---|---|---|---|---|---|
| Gherlone et al. | Comorbidities | COPD | Dry mouth | Multivariable analysis | (OR= 9.10, 95% CI: 1.8 -68.49) |
| Stavem et al. | Severity | Number of symptoms (10–23) | Symptoms at follow-up | Multivariable negative binomial regression analysis | (OR= 4.16, 95% CI:2.57 to 6.72, p<0.001) |
| | Comorbidities | ≥2 | | | (OR=2.52, 95%CI: 1.58 to 4.02, p<0.001) |
| Baricich et al. | Severity | ICU admission | Physical impairment | Multivariable logistic regression model | (OR: 3.1, 95%CI: 1.3-7.9, p=0.01) |
| | Age | Age | walking ability (SPPB) | | p <0.02 |
| | Comorbidities | Number or comorbidities | walking ability (SPPB)<br>2MWT | | p <0.01<br>p <0.04 |
| | Sex | Male gender | SPPB total score | | p <0.01 |
| Jacobson et al. | Ethnicity | Latin ethnicity | lower expected 6-MWT | Multivariate analysis | (-7.40 [-11.55-{-3.25}], p=0.001 |
| | Comorbidities | BMI | | | (-0.52 [-0.81-{-0.22}], p=0.001) |
| | Severity | Persistence of symptoms at follow up | Shortness of breath | | P=0.004 |
| Petersen et al. | Age | Individuals in age group 50-66 compared with the youngest groups:<br>0-17 years<br>18-34 years | Persistent symptoms | Age-stratified analysis | p=0.003<br>p=0.001 |
| Alharthy et al. | Severity | Increased incidence of dyspnoea and fever prior to hospital admission, decreased ICU admission PaO2/FiO2 ratio < 100, longer duration of mechanical ventilation, increased inflammatory biomarkers such as lactate dehydrogenase, ferritin, and D-dimers on ICU admission, and significant lung abnormalities detected by LUS | Persistent symptoms | Continuous variables using the Wilcoxon rank sum or the student's t-test. Categorical variables were examined using the Fisher's exact test or the Chi square test | p < 0.05 |

| | | | | | |
|---|---|---|---|---|---|
| Anastasio et al. | Severity | Pneumonia and ARDS | Shortness of breath | Pearson's correlation coefficient and Cox regression were used | Patients who developed ARDS showed higher SBP (p=0.05) and DBP (p=0.02) and lower SpO2 during 6 MWT (p=0.004), FVC (p=0.004) and TLC (p<0.001). Patients without ARDS showed higher SR (p<0.001), RV (p<0.001), TLC (p<0.001) and RV/TLC (p=0.05). |
| Han et al. | Severity | Higher baseline CT lung involvement score (>=18 out of a possible score of 25) | Fibrotic-like changes in the lung at 6 months | Multivariate analysis | (OR: 4.2, 95%CI: 1.2-14) |
| Blanco et al. | Severity | Severity of the disease | DLCO <80% and a lower serum lactate dehydrogenase level | Multivariate analysis | DLCO<80% (OR 5.92; 95%CI 2.28–15.37; p < 0.0001) Serum lactate dehydrogenase (OR 0.98; 95%CI 0.97–0.99) |
| Lerum et al. | Severity | ICU admission | Persistent CT abnormalities and problems in usual activities | Mann–Whitney U-tests or Chi-squared tests | p=.031 |
| Bellan et al. | Severity | Higher DLCO | Decreased risk of physical impairment | Univariate analysis and logistic regression models | (OR, 0.96 [95% CI, 0.94-0.98]; P < .001) |
| | Comorbidities | COPD | Increase risk of physical impairment | | (OR, 12.70 [95% CI, 1.41-114.85]; P = .02) |
| Sonnweber et al. | Severity | Age, gender, and pre-existing diseases such as cardiovascular diseases, pulmonary diseases, diabetes mellitus type 2, and malignancy | Persistence of symptoms, patient performance status, and CT findings at follow-up | Friedman's or Wilcoxon signed-rank test | p=0.042 to p<0.001 |
| Mendez et al. | Sex | Female sex | Impaired DLCO | Linear regression analysis | 0.002 |
| | Severity | ICU patients | Pulmonary embolism | | p<0.001 |
| | | D-dimer levels | Impaired DLCO | | p= 0.011 |
| Blanco et al. | Severity | Lower serum LDH levels | Impaired DLCO | Multivariate analysis | OR 0.98; 95% CI 0.97-0.99; p 0.002 |
| Qin et al. | Severity | Higher TSS of the chest and ARDS lymphocyte count, MPA diameter on admission and ARDS | Impaired DLCO | Univariable analysis | TSS>10.5 (OR: 10.5; 95%CI: 2.5-44.1; P=0.001) ARDS ( OR: 4.6; 95%CI: 1.4-15.5; P=0.014 ) |
| | | Long hospital stay | Lung sequelae | | |

| | | | | | |
|---|---|---|---|---|---|
| Rass et al. | Severity | ICU patients | New neurological diseases | Chi-square or Kruskal-Wallis test | P=0.001 |
| | Age | Elderly | Neurological signs | NR | NR |
| Weng et al. | Severity | Less severe (Lower frequency of supplemental oxygen therapy (79% vs 94%; p=0·016), and lower frequency of ICU admission | Gastrointestinal sequelae | Univariable and multivariable logistic regressions | p=0·016 |
| | | Treated more often with proton pump inhibitors (PPIs) and corticosteroids and were less frequently treated with enteral nutrition | | | PPI (p=0.000)<br>Corticosteroids (p=0.024)<br>Enteral nutrition (p=0.007) |
| Arnold et al. | Severity | Severe cases | Lower physical score | Mann Whitney-U and Kruskal Wallis tests for continuous data and Fisher's exact test or Chi-squared testing for categorical data. | NR |
| Sibila et al. | Sex | Male gender | Spirometric abnormalities 3 months after discharge,= | NR | Reduced FEV1: (76.9% vs 51.2%, p = 0.005)<br>Reduced FVC: (76.3% vs 51.6%, p = 0.008) |
| | Comorbidities | Cardiovascular disease and diabetes | | | Reduced FEV1:<br>Cardiovascular disease (34.2% vs 9.4%, p = 0.001)<br>Diabetes (28.9% vs 12%, p = 0.02)<br>Reduced FVC:<br>Cardiovascular disease (29.7% vs 11.0%, p = 0.009) |
| Huang et al. | Severity | Participants with severity scale 5–6 | Higher risk of lung diffusion impairment, anxiety or depression, and fatigue or muscle weakness | Multivariable analysis | OR 4·60 (95% CI 1·85–11·48) for diffusion impairment, OR 1·77 (1·05–2·97) for anxiety or depression, and OR 2·69 (1·46–4·96) for fatigue or muscle weakness |
| | Sex | Female sex | | | |

ARDS: Acute respiratory distress syndrome; BMI: Body mass index; CT: Computerised Topography; DCLO: diffusing capacity for carbon monoxide; ICU: Intensive care unit; LDH: Lactate dehydrogenase; LUS: lung ultrasound; MWT: minute walking test; NR: Not reported; OR: Odds Ratio; PCSF: post covid functional status; QoL: Quality of life; SPPB: Short Physical Performance Battery test; TSS: Toxic shock syndrome
